# Convalescent plasma in patients admitted to hospital with COVID-19 (RECOVERY): a randomised, controlled, open-label, platform trial

**DOI:** 10.1101/2021.03.09.21252736

**Authors:** The RECOVERY Collaborative Group, Peter W Horby, Lise Estcourt, Leon Peto, Jonathan R Emberson, Natalie Staplin, Enti Spata, Guilherme Pessoa-Amorim, Mark Campbell, Alistair Roddick, Nigel E Brunskill, Tina George, Daniel Zehnder, Simon Tiberi, Ni Ni Aung, Alison Uriel, John Widdrington, George Koshy, Thomas Brown, Steven Scott, J Kenneth Baillie, Maya H Buch, Lucy C Chappell, Jeremy N Day, Saul N Faust, Thomas Jaki, Katie Jeffery, Edmund Juszczak, Wei Shen Lim, Alan Montgomery, Andrew Mumford, Kathryn Rowan, Guy Thwaites, Marion Mafham, David Roberts, Richard Haynes, Martin J Landray

## Abstract

**Background:** Treatment of COVID-19 patients with plasma containing anti-SARS-CoV-2 antibodies may have a beneficial effect on clinical outcomes. We aimed to evaluate the safety and efficacy of convalescent plasma in patients admitted to hospital with COVID-19.

**Methods:** In this randomised, controlled, open-label, platform trial (Randomised Evaluation of COVID-19 Therapy [RECOVERY]) several possible treatments are being compared with usual care in patients hospitalised with COVID-19 in the UK. Eligible and consenting patients were randomly allocated to receive either usual care plus high titre convalescent plasma or usual care alone. The primary outcome was 28-day mortality.

**Findings:** Between 28 May 2020 and 15 January 2021, 5795 patients were randomly allocated to receive convalescent plasma and 5763 to usual care alone. There was no significant difference in 28-day mortality between the two groups: 1398 (24%) of 5795 patients allocated convalescent plasma and 1408 (24%) of 5763 patients allocated usual care died within 28 days (rate ratio [RR] 1·00; 95% confidence interval [CI] 0·93 to 1·07; p=0·93). The 28-day mortality rate ratio was similar in all prespecified subgroups of patients, including in those patients without detectable SARS-CoV-2 antibodies at randomisation. Allocation to convalescent plasma had no significant effect on the proportion of patients discharged from hospital within 28 days (66% *vs*. 67%; rate ratio 0·98; 95% CI 0·94-1·03, p=0·50). Among those not on invasive mechanical ventilation at baseline, there was no significant difference in the proportion meeting the composite endpoint of progression to invasive mechanical ventilation or death (28% *vs*. 29%; rate ratio 0·99; 95% CI 0·93-1·05, p=0·79).

**Interpretation:** Among patients hospitalised with COVID-19, high-titre convalescent plasma did not improve survival or other prespecified clinical outcomes.

**Funding:** UK Research and Innovation (Medical Research Council) and National Institute of Health Research (Grant refs: MC_PC_19056; COV19-RECPLA).

## INTRODUCTION

A substantial proportion of individuals infected with severe acute respiratory syndrome coronavirus 2 (SARS-CoV-2) require hospital care, which can progress to critical illness with hypoxic respiratory failure. In those with severe Coronavirus Disease-19 (COVID-19), immunomodulation with corticosteroids and interleukin-6 receptor antagonists has been shown to improve survival.^1,2^ Treatments that effectively inhibit viral replication may reduce tissue damage and allow time for the host to develop an adaptive immune response that will clear the infection. To date, however, no treatment directed against the virus has been shown to reduce mortality (although remdesivir may shorten the duration of hospital stay).^3^

Humoral immunity is a key component of the immune response to SARS-CoV-2 and matures over several weeks following infection. Anti-SARS-CoV-2 antibodies are detectable at a mean of 13 days after symptom onset, but neutralising titres do not peak until day 23 and there is wide variation in both the timing of seroconversion and peak antibody levels between infected individuals.^4^ While patients with severe COVID-19 generally have higher final antibody concentrations than those with mild disease, their antibody responses are delayed.^5^ Antibodies may modulate acute viral disease either by a direct antiviral effect, binding and neutralizing free virus, or indirectly by activating antiviral pathways such as the complement cascade, phagocytosis and cellular cytotoxicity. Conversely, there is also a possibility that antibodies may enhance disease, either by promoting viral entry or by pro-inflammatory mechanisms such as Fcγ receptor stimulation.^6^

Convalescent plasma has been used for over a hundred years as passive immunotherapy for influenza pneumonia, and more recently for SARS-CoV-1. While observational studies have suggested it may reduce mortality in severe viral respiratory infections, randomised evidence remains limited and inconclusive.^7^ Convalescent plasma has been used widely outside of clinical trials, including by tens of thousands of patients in the United States Food and Drugs Administration (FDA) Expanded Access Program. An observational (non-randomised) analysis of 3082 patients who received convalescent plasma as part of that programme, reported that 30-day mortality was lower in those who had not received mechanical ventilation before transfusion with higher-titre plasma (containing higher concentrations of anti-SARS-CoV-2 spike IgG) compared to those transfused with lower-titre plasma.^8^ A number of randomised trials of convalescent plasma in patients hospitalised with COVID-19 have been reported but these trials have all been small and inconclusive.^9-17^ Moreover, hospitalised patients with COVID-19 are heterogeneous, and any benefit of convalescent plasma could depend on the stage of disease, i.e. possibly being limited to those with milder disease, early in the course of their illness or those who have not mounted an effective antibody response.^12^ The efficacy of convalescent plasma as a treatment for patients hospitalised with COVID-19 is, therefore, currently uncertain. Here, we report the results of a large randomised trial to evaluate the efficacy and safety of convalescent plasma in patients hospitalised with COVID-19.

## METHODS

### Study design and participants

The Randomised Evaluation of COVID-19 therapy (RECOVERY) trial is an investigator-initiated, individually randomised, controlled, open-label, adaptive platform trial to evaluate the effects of potential treatments in patients hospitalised with COVID-19. Details of the trial design and results for other evaluated treatments (dexamethasone, hydroxychloroquine, lopinavir-ritonavir, azithromycin and tocilizumab) have been published previously.^1,2^ The trial is conducted at 177 National Health Service (NHS) hospital organizations in the United Kingdom (appendix pp 5-28), supported by the National Institute for Health Research Clinical Research Network. The trial is coordinated by the the trial sponsor, the Nuffield Department of Population Health at the University of Oxford (Oxford, UK). The trial is conducted in accordance with the principles of the International Conference on Harmonisation–Good Clinical Practice guidelines and approved by the UK Medicines and Healthcare products Regulatory Agency (MHRA) and the Cambridge East Research Ethics Committee (ref: 20/EE/0101). The protocol, statistical analysis plan, and additional information are available on the trial website www.recoverytrial.net.

Hospitalised patients of any age were eligible for the trial if they had clinically suspected or laboratory-confirmed SARS-CoV-2 infection and no medical history that might, in the opinion of the attending clinician, put them at significant risk if they were to participate in the trial. Written informed consent was obtained from all patients or from their legal representative if they were too unwell or unable to provide consent.

### Randomisation and masking

Baseline data collected using a web-based case report form that included demographics, level of respiratory support, major comorbidities, suitability of the trial treatment for a particular patient and treatment availability at the trial site site (appendix pp 35-37). Patients had a serum sample taken prior to randomisation for the purpose of assessing the presence of antibodies against SARS-CoV-2. Eligible and consenting patients were allocated in a ratio of 1:1:1 to either usual care, usual care plus convalescent plasma or (from 18 September 2020) usual care plus REGN-COV2 (a combination of two monoclonal antibodies directed against SARS-CoV-2 spike protein). The REGN-COV2 evaluation is ongoing and not reported here. Randomisation was web-based simple (unstratified) randomisation with allocation concealment (appendix pp 33-34). For some patients, convalescent plasma was either declined, unavailable at the trial site at the time of enrolment, or considered in the opinion of the attending doctor to be definitely contraindicated (e.g. known moderate or severe allergy to blood components or unwilling to receive a blood product). These patients were ineligible for randomisation to the comparison of convalescent plasma versus usual care.

As a platform trial and in a factorial design, patients could be simultaneously randomised to other treatment groups: i) hydroxychloroquine or dexamethasone or azithromycin or lopinavir-ritonavir versus usual care, ii) aspirin versus usual care, and iii) colchicine versus usual care (appendix pp 33-34). The trial also allowed a subsequent randomisation for patients with progressive COVID-19 (evidence of hypoxia and a hyper-inflammatory state) to tocilizumab versus usual care. Participants and local study staff were not masked to the allocated treatment. Several of these treatment arms were added to or removed from the protocol over the period that convalescent plasma was evaluated (appendix pp 29-34). The trial steering committee, investigators, and all other individuals involved in the trial were masked to outcome data during the trial.

### Procedures

Convalescent plasma donors were recruited and screened by the four UK blood services: NHS Blood and Transplant; Northern Ireland Blood Transfusion Service; Scottish National Blood Transfusion Service; and the Welsh Blood Service (appendix pp 2-4 and p 29). Only plasma donations with sample to cut-off (S/CO) ratio of 6.0 or above on the EUROIMMUN IgG enzyme-linked immunosorbent assay (ELISA) test targeting the spike (S) glycoprotein (PerkinElmer, London, UK) were supplied for the RECOVERY trial use (appendix p 29). This assay cut-off was previously demonstrated to be associated with the presence of neutralising antibody titres of ≥1:100 in convalescent plasma.^18^ The United States Food and Drug Administration (US FDA) have determined that convalescent plasma with a EUROIMMUN S/CO of ≥3.5 qualifies as high-titre and can be used for the treatment of hospitalised patients under an Emergency use Authorization (EUA).^19^ For those allocated convalescent plasma, two units (275mls ± 75mls) were given intravenously, the first as soon as possible after randomisation and the second (from a different donor) the following day and at least 12 hours after the first. Early safety outcomes were recorded using an online form 72 hours following randomisation (appendix pp 38-42). An online follow-up form was completed when patients were discharged, had died, or at 28 days after randomisation, whichever occurred earlier (appendix pp 43-49). Information was recorded on adherence to allocated trial treatment, receipt of other COVID-19 treatments, duration of admission, receipt of respiratory or renal support, and vital status (including cause of death). In addition, routine health care and registry data were obtained including information on vital status at day 28 (with date and cause of death); discharge from hospital; and receipt of respiratory support or renal replacement therapy.

### Measurement of participant baseline SARS-CoV-2 serostatus

Baseline SARS-CoV-2 serostatus for each participant was determined using serum samples taken at the time of randomisation. Analysis was performed at a central laboratory using a validated 384-well plate indirect ELISA (appendix p 29).^20^ Participants were categorised as seropositive or seronegative using a predefined assay threshold that has ≥99% sensitivity and specificity in detecting individuals with SARs-CoV-2 infection at least 20 days previously.^20^

### Outcomes

Outcomes were assessed at 28 days after randomisation, with further analyses specified at six months. The primary outcome was all-cause mortality. Secondary outcomes were time to discharge from hospital and, among patients not receiving invasive mechanical ventilation at randomisation, subsequent receipt of invasive mechanical ventilation (including extra-corporeal membrane oxygenation) or death. Prespecified, subsidiary clinical outcomes included receipt of ventilation, time to successful cessation of invasive mechanical ventilation (defined as removal of invasive mechanical ventilation within, and survival to, 28 days), and use of renal dialysis or haemofiltration.

Prespecified safety outcomes were transfusion related adverse events at 72 hours following randomisation (worsening respiratory status, suspected transfusion reaction, fever, hypotension, haemolysis, and thrombotic events), cause-specific mortality, and major cardiac arrhythmia. Information on serious adverse reactions to convalescent plasma was collected in an expedited fashion via the existing NHS Serious Hazards Of Tranfusion (SHOT) haemovigilence scheme.

### Statistical Analysis

In accordance with the statistical analysis plan, an intention-to-treat comparison was conducted between patients randomised to convalescent plasma and patients randomised to usual care in those for whom convalescent plasma was both available and suitable as a treatment. For the primary outcome of 28-day mortality, the log-rank observed minus expected statistic and its variance were used both to test the null hypothesis of equal survival curves (i.e. the log-rank test) and to calculate the one-step estimate of the average mortality rate ratio. We constructed Kaplan-Meier survival curves to display cumulative mortality over the 28-day period. We used similar methods to analyse time to hospital discharge and successful cessation of invasive mechanical ventilation, with those patients who died in hospital right-censored on day 29. Median time to discharge was derived from Kaplan-Meier estimates. For the prespecified, composite, secondary outcome of progression to invasive mechanical ventilation or death within 28 days (among those not receiving invasive mechanical ventilation at randomisation) and the subsidiary clinical outcomes of receipt of ventilation and use of haemodialysis or haemofiltration, the precise dates were not available and so the risk ratio was estimated instead. (Through the play of chance, a slightly lower proportion of males were allocated convalescent plasma than usual care; analyses adjusted for sex are provided in the appendix [webtable 7] and are virtually identical to the main results shown.) Sensitivity analyses of the primary and secondary outcomes were conducted among those patients with a positive PCR test for SARS-COV-2.

Prespecified analyses of the primary outcome were performed in seven subgroups defined by characteristics at randomisation: age, sex, ethnicity, level of respiratory support received, days since symptom onset, use of systemic corticosteroids, and presence of anti-SARS-CoV-2 antibody. Observed effects within these subgroup categories were compared using a chi-squared test for heterogeneity or trend. *Post-hoc* exploratory analyses included further examination by days since symptom according to four rather than two levels and by level of respiratory support by sub-dividing the ‘oxygen only’ group into three sub-categories. In late 2020, a new SARS-CoV-2 variant, named B.1.1.7, with multiple substitutions in the receptor binding domain of the spike glycoprotein emerged in southeast England and rapidly grew to become the dominant virus variant throughout the UK.^21^ Convalescent plasma from individuals infected prior to the emergence of B.1.1.7 show a modest reduction in ability to neutralize B.1.1.7 compared with earlier SARS-CoV-2 virus variants.^22^ The clinical significance of this reduced in vitro neutralisation is not known. To assess if there was evidence of a difference in the effectiveness of convalescent plasma before and after the emergence of B.1.1.7, a further *post-hoc* exploratory analysis was done of the primary outcome comparing effects in those randomised before 1 December 2020 with those randomised from 1 December 2020 onwards.^21^

Estimates of rate and risk ratios are shown with 95% confidence intervals. All p-values are 2-sided and are shown without adjustment for multiple testing. The full database is held by the trial team who pooled the data from trial sites and performed the analyses at the Nuffield Department of Population Health, University of Oxford.

Analyses were performed using SAS version 9.4 and R version 3.4. The trial is registered with ISRCTN (50189673) and clinicaltrials.gov (<u>NCT04381936</u>).

### Sample size and decision to stop enrolment

As stated in the protocol, appropriate sample sizes could not be estimated when the trial was being planned at the start of the COVID-19 pandemic. During the trial, external data suggested that any benefits of antibody-based therapies may be greater among those patients who had not raised an adequate antibody response of their own.^12^ Consequently, while still blind to the results of the trial, the RECOVERY steering committee determined that the trial should enrol sufficient patients to provide at least 90% power at a two-sided p-value of 0.01 to detect a proportional reduction in 28-day mortality of one-fifth among those patients with and, separately, without detectable SARS-CoV-2 antibodies at randomisation (appendix p 34).

On 7^th^ January 2021, the independent data monitoring committee (DMC) conducted a routine review of the data and recommended that the chief investigators pause the recruitment to the convalescent plasma comparison in those patients receiving invasive mechanical ventilation (including extracorporeal membrane oxygenation) at the time of randomisation. At the same time, the DMC recommended that recruitment to the convalescent plasma comparison continue for all other eligible patients.

On 14^th^ January 2021, the DMC conducted another routine review of the data and notified the chief investigators that there was no convincing evidence that further recruitment would provide conclusive proof of worthwhile mortality benefit either overall or in any pre-specified subgroup. The DMC therefore recommended that recruitment to the convalescent plasma portion of the study should cease and follow-up be completed. Enrolment of patients to the convalescent plasma group was closed on 15^th^ January 2021 and the preliminary result for the primary outcome was made public.

## RESULTS

### Patients

Between 28 May 2020 and 15 January 2021, 13127 (81%) of 16287 patients enroled into the RECOVERY trial, were eligible to be randomised to convalescent plasma (that is, convalescent plasma was available in the hospital at the time and the patient had no known contraindication to convalescent plasma (figure 1). Of these, 5795 were randomised to convalescent plasma plus usual care and 5763 were randomised to usual care alone (figure 1), with the remainder being randomised to receive REGN-COV2. The mean age of trial patients in this comparison was 63.5 (SD 14.7) years and the median time from symptom onset to randomization was 9 days (IQR 6 – 12) (table 1, webtable 1). At randomisation, 617 (5%) were receiving invasive mechanical ventilation, 10044 (87%) were receiving oxygen only (with or without non-invasive respiratory support), and 897 (8%) were receiving no oxygen therapy (webtable 1). 92% of patients were receiving corticosteroids at time of randomisation.

**Table 1:** Baseline characteristics.

|  | **Convalescent**<br>**Plasma**<br>**(n=5795)** | **Usual Care**<br>**(n=5763)** |
| --- | --- | --- |
| Age, years | 63.6 (14.7) | 63.4 (14.6) |
| <70* | 3705 (64) | 3748 (65) |
| 70 to 79 | 1310 (23) | 1280 (22) |
| ≥80 | 780 (13) | 735 (13) |
| Sex |  |  |
| Men | 3643 (63) | 3787 (66) |
| Women† | 2152 (37) | 1976 (34) |
| Ethnicity |  |  |
| White | 4362 (75) | 4293 (74) |
| Black, Asian, and minority ethnic | 853 (15) | 889 (15) |
| Unknown | 580 (10) | 581 (10) |
| Number of days since symptom onset | 9 (6-12) | 9 (6-12) |
| Number of days since admission to hospital | 2 (1-3) | 2 (1-4) |
| Respiratory support received |  |  |
| No oxygen received | 442 (8) | 455 (8) |
| Oxygen only‡ | 5051 (87) | 4993 (87) |
| Invasive mechanical ventilation | 302 (5) | 315 (5) |
| Previous diseases |  |  |
| Diabetes | 1535 (26) | 1569 (27) |
| Heart disease | 1267 (22) | 1309 (23) |
| Chronic lung disease | 1385 (24) | 1328 (23) |
| Tuberculosis | 20 (<1) | 23 (<1) |
| HIV | 17 (<1) | 19 (<1) |
| Severe liver disease§ | 70 (1) | 72 (1) |
| Severe kidney impairment¶ | 323 (6) | 293 (5) |
| Any of the above | 3203 (55) | 3222 (56) |
| SARS-CoV-2 PCR test result |  |  |
| Positive | 5581 (96) | 5559 (96) |
| Negative | 125 (2) | 113 (2) |
| Unknown | 89 (2) | 91 (2) |
| Patient SARS-CoV-2 antibody test result |  |  |
| Positive | 3022 (52) | 2752 (48) |
| Negative | 1982 (34) | 1629 (28) |
| Missing | 791 (14) | 1382 (24) |
| Corticosteroids received |  |  |
| Yes | 5370 (93) | 5311 (92) |
| No | 391 (7) | 413 (7) |
| Not recorded | 34 (1) | 39 (1) |
| Other randomised treatments |  |  |
| Lopinavir-ritonavir | 5 (<1) | 14 (<1) |
| Dexamethasone | 3 (<1) | 3 (<1) |
| Hydroxychloroquine | 1 (<1) | 0 |
| Azithromycin | 587 (10) | 585 (10) |
| Colchicine | 792 (14) | 791 (14) |
| Aspirin | 1266 (22) | 1207 (21) |

Data are mean (SD), n (%), or median (IQR). \*Includes 26 children (<18 years). † Includes 28 pregnant women. ‡ Includes non-invasive ventilation. § Defined as requiring ongoing specialist care. ¶ Defined as estimated glomerular filtration rate <30 mL/min per 1.73 m<sup>2</sup>

**Figure 1:**
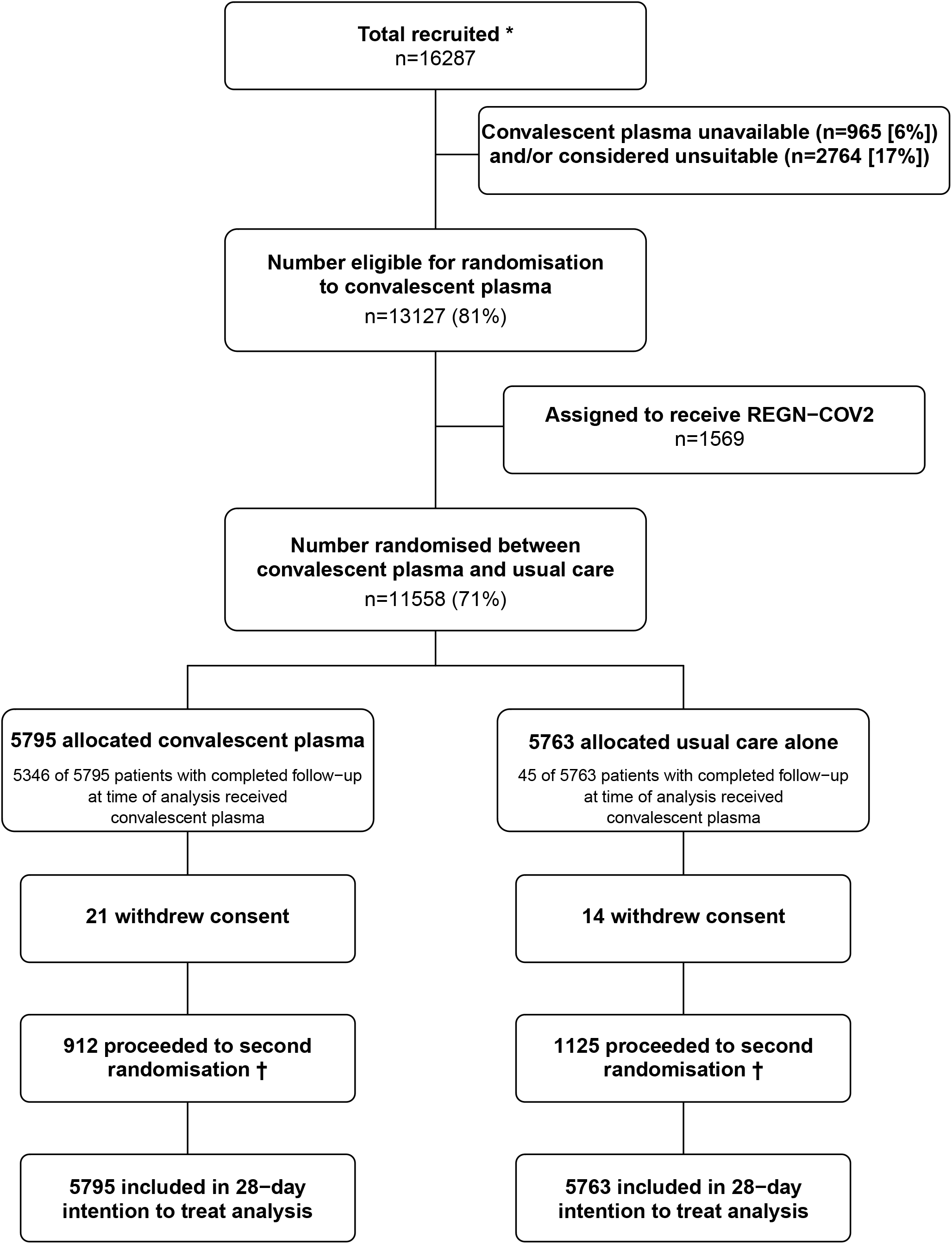
Trial profile - Flow of patients through the RECOVERY trial. *Number recruited overall during period that patients could be recruited into convalescent plasma comparison. † A second randomisation to tocilizumab versus usual care in patients with hypoxia and C-reactive protein ≥75 mg/L was introduced in protocol version 4.0. 426 patients in the convalescent plasma arm were randomised to tocilizumab vs. 486 randomised to usual care alone. 573 patients in the usual care arm were randomised to tocilizumab vs. 552 patients randomised to usual care alone.

Of the 9385 (81%) patients for whom a baseline serology result was available, 5774 (62%) were SARS-CoV-2 antibody seropositive (webtable 1). Patients were more likely to be seronegative if they were older, female, white, had shorter duration of symptoms, were receiving less intensive respiratory support, or were SARS-CoV-2 RNA negative by PCR (webtable 2). There was an imbalance in the availability of a baseline serology sample, with more missing samples in the usual care arm (table1). (This likely reflects a mistaken belief by some trial staff that a serology sample was only required in patients allocated to convalescent plasma.)

Among the 5795 patients allocated to convalescent plasma, 4675 (81%) received two units, 671 (12%) received one unit, and 449 (8%) received no units (webtable 3). Only two patients received both convalescent plasma units from the same donor. Forty-five (1%) patients allocated to usual care received convalescent plasma. Use of corticosteroids and remdesivir following randomisation was similar among patients allocated convalescent plasma and among those allocated usual care (webtable 3). Fewer patients received tocilizumab or sarilumab in the convalescent plasma group (8% vs. 10%, webtable 3).

There was no significant difference in 28-day mortality between the two randomised groups, with death reported in 1398 of 5795 patients (24%) allocated convalescent plasma versus 1408 of 5763 patients (24%) allocated usual care (rate ratio, 1·00; 95% confidence interval [CI], 0·93 to 1·07; P=0·93) (figure 2). We observed similar results across all subgroups with no evidence of heterogeneity of effect in either the pre-specified (figure 3) or the exploratory *post-hoc* (webfigure 1) subgroup analyses, and similar results in analyses restricted to those patients with a positive SARS-CoV-2 test (rate ratio 1·00; 95% CI, 0·93 to 1·08; P=0·98). Although 28-day mortality was higher among those patients who were seronegative at randomisation, the proportional effect of allocation to convalescent plasma on 28-day mortality was similar among seropositive patients (19% versus 18%; rate ratio, 1·05; 95% CI, 0·93 to 1·19) and seronegative patients (32% versus 34%; rate ratio, 0·94; 95% CI, 0·84 to 1·06) (figure 3; webfigure 2).

**Figure 2:**
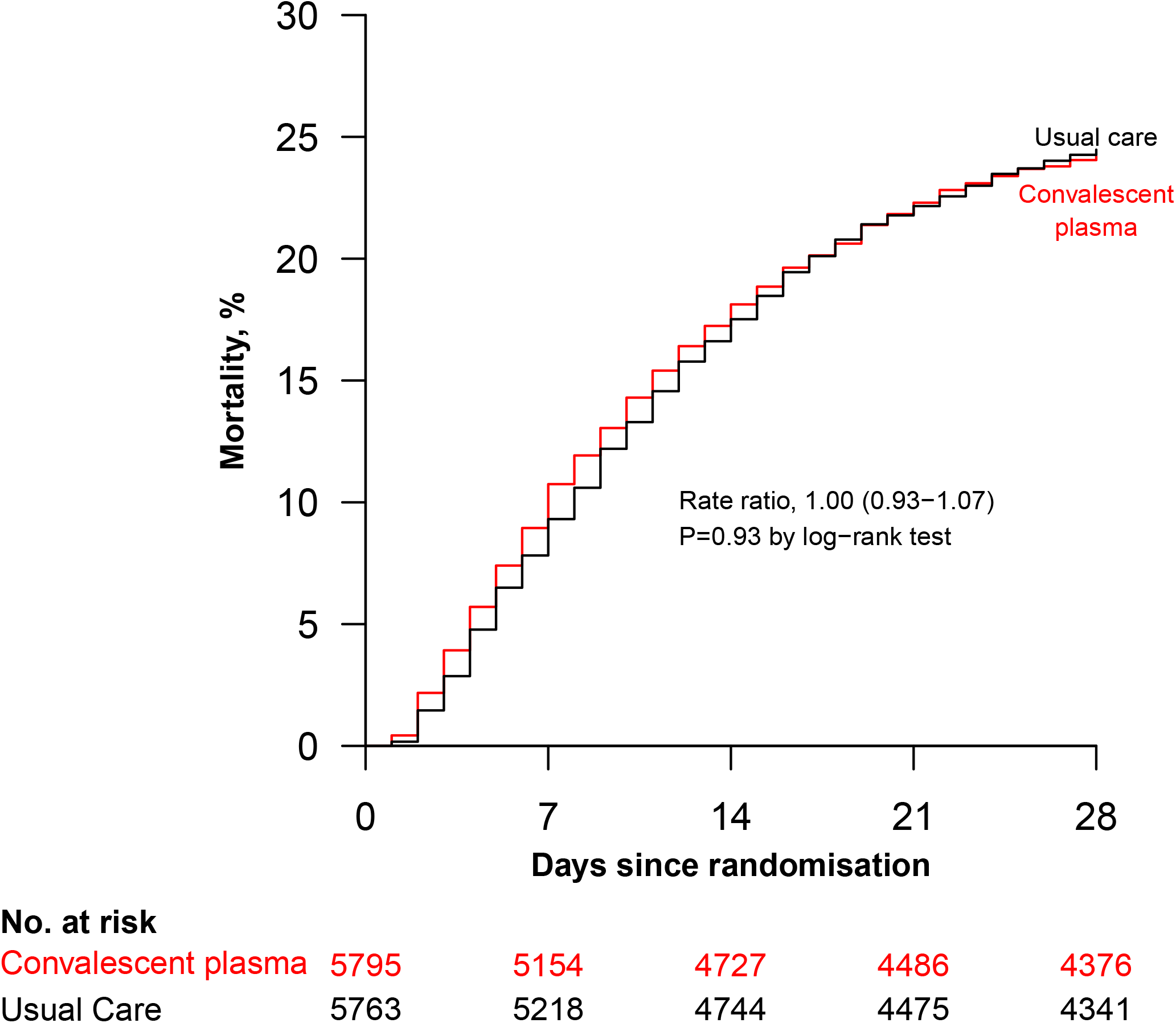
Effect of allocation to convalescent plasma on 28-day mortality.

**Figure 3:**
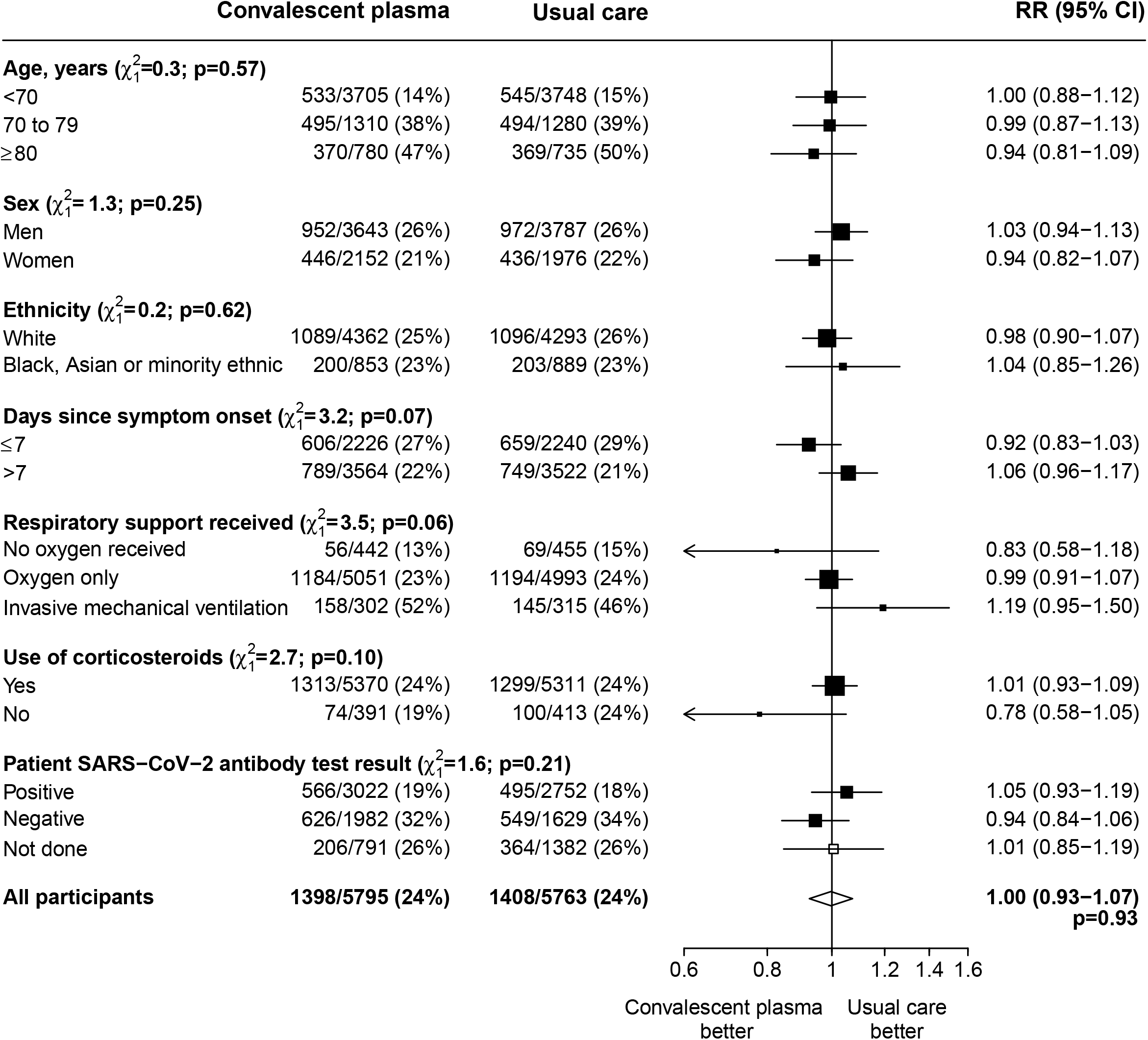
Effect of allocation to convalescent plasma on 28-day mortality by prespecified characteristics at randomisation. Subgroup− specific rate ratio estimates are represented by squares (with areas of the squares proportional to the amount of statistical information) and the lines through them correspond to the 95% CIs. The ethnicity, days since onset and use of corticosteroids subgroups exclude those with missing data, but these patients are included in the overall summary diamond. Information on use of corticosteroids was collected from 18 June 2020 onwards following announcement of the results of the dexamethasone comparison from the RECOVERY trial.

The median time to discharge was 11 days in both those allocated convalescent plasma and those allocted usual care, and allocation to convalescent plasma was associated with a similar probability of discharge alive within 28 days compared to usual care (66% vs. 67%, rate ratio 0·98 95% CI 0·94 to 1·03, p=0·50) (table 2). Among those not receiving invasive mechanical ventilation at baseline, the number of patients progressing to the prespecified composite secondary outcome of invasive mechanical ventilation or death was similar for those allocated to convalescent plasma or usual care (28% versus 29%, risk ratio 0·99 95% CI 0·93 to 1·05, p=0·79) (table 2). For both of these secondary outcomes, there was some evidence of heterogeneity by patient SARS-CoV-2 antibody test result, with slightly more favourable outcomes with convalescent plasma seen among seronegative than among seropositive patients (webfigures 3 and 4). Results were consistent across all other pre-specified subgroups of patients.

**Table 2:** Primary, Secondary and Subsidiary Outcomes.

|  | Convalescent<br>plasma<br>(n=5795) | Usual Care<br>(n=5763) | RR (95% CI) | p value |
| --- | --- | --- | --- | --- |
| **Primary outcome** |  |  |  |  |
| Mortality at 28 days | 1398 (24%) | 1408 (24%) | 1.00 (0.93-1.07) | 0.93 |
| **Secondary outcomes** |  |  |  |  |
| Median duration of hospitalization, days | 11 | 11 | - | - |
| Discharged from hospital within 28 days | 3850 (66%) | 3846 (67%) | 0.98 (0.94-1.03) | 0.50 |
| Invasive mechanical ventilation or death* | 1561/5493 (28%) | 1561/5448 (29%) | 0.99 (0.93-1.05) | 0.79 |
| Invasive mechanical ventilation | 670/5493 (12%) | 681/5448 (13%) | 0.98 (0.88-1.08) | 0.63 |
| Death | 1240/5493 (23%) | 1263/5448 (23%) | 0.97 (0.91-1.04) | 0.45 |
| **Subsidiary outcomes** |  |  |  |  |
| Use of ventilation † | 860/3564 (24%) | 863/3441 (25%) | 0.96 (0.89-1.04) | 0.36 |
| Non-invasive ventilation | 822/3564 (23%) | 821/3441 (24%) | 0.97 (0.89-1.05) | 0.43 |
| Invasive mechanical ventilation | 226/3564 (6%) | 237/3441 (7%) | 0.92 (0.77-1.10) | 0.36 |
| Successful cessation of invasive mechanical ventilation ‡ | 87/302 (29%) | 112/315 (36%) | 0.77 (0.59-1.03) | 0.07 |
| Renal replacement therapy § | 258/5729 (5%) | 249/5713 (4%) | 1.03 (0.87-1.22) | 0.71 |

Data are n (%) or n/N (%). RR=rate ratio for the outcomes of 28-day mortality, hospital discharge, and successful cessation of invasive mechanical ventilation, and risk ratio for other outcomes.
\* Analyses exclude those on invasive mechanical ventilation at randomisation.
† Analyses exclude those on invasive or non-invasive ventilation at randomisation.
‡ Analyses exclude those not receiving invasive mechanical ventilation at randomisation.
§ Analyses exclude those on renal replacement therapy at randomisation.

**Figure 4:**
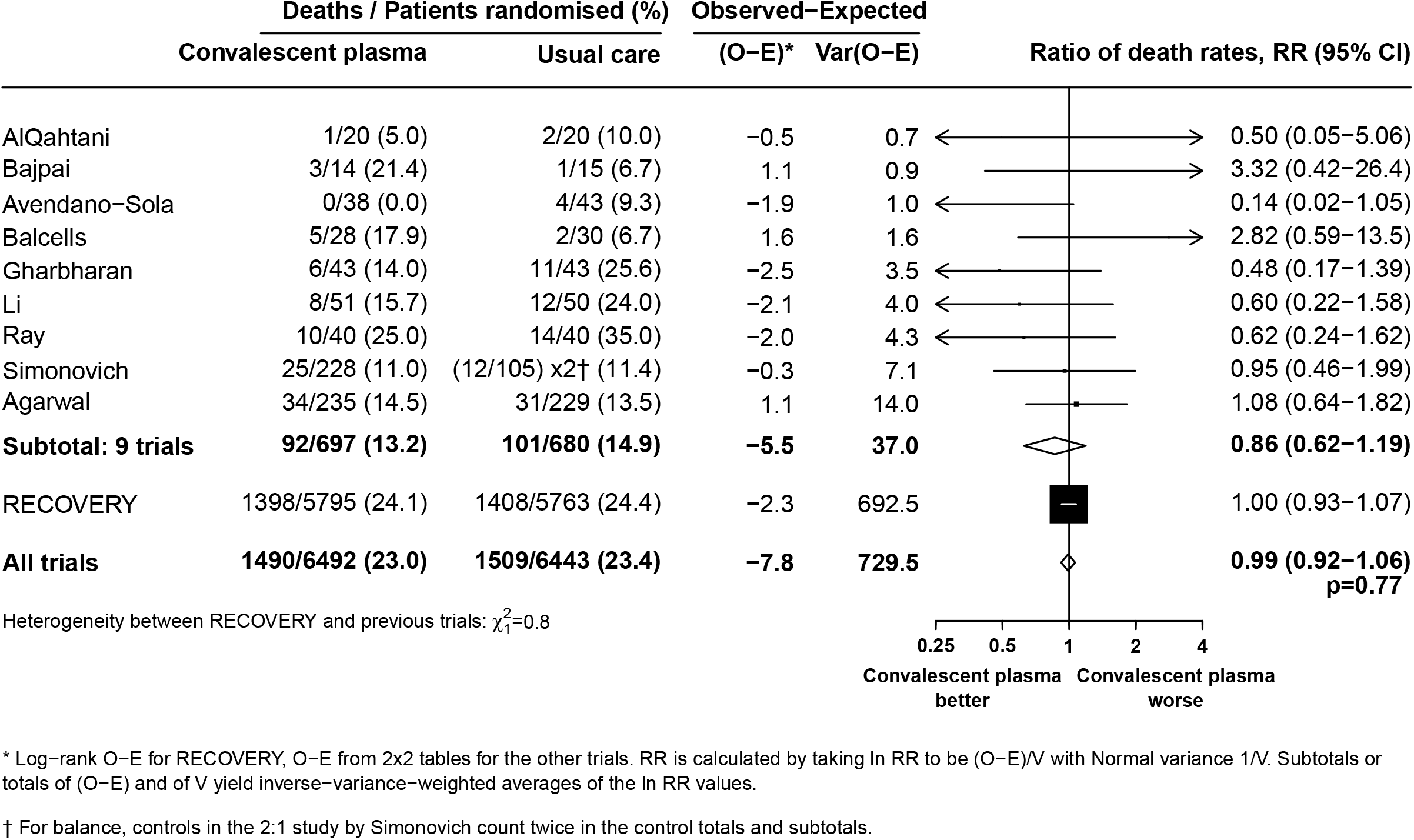
Convalescent plasma vs. usual care in patients hospitalised with COVID – meta-analysis of mortality in RECOVERY and other trials. *Log− rank O−E for RECOVERY, O− E from 2×2 tables for the other trials. RR is calculated by taking ln RR to be (O− E)/V with Normal variance 1/V. Subtotals or totals of (O− E) and of V yield inverse− variance− weighted averages of the ln RR values. † For balance, controls in the 2:1 study by Simonovich count twice in the control totals and subtotal

We observed no significant differences in the prespecified subsidiary clinical outcomes of use of ventilation, successful cessation of invasive mechanical ventilation, or progression to use of renal replacement therapy (table 2).

We observed no significant differences in cause-specific mortality (webtable 4). Within the first 72 hours after randomisation, severe allergic reactions were reported for 16 patients in the convalescent plasma group vs. 2 patients in the usual care group. The frequency of sudden worsening in respiratory status, temperature >39°C or ≥2°C rise above baseline, sudden hypotension, clinical haemolysis, and thrombotic events were broadly similar in the two groups (webtable 5). We also observed no significant differences in the frequency of major cardiac arrhythmia (webtable 6). There were 13 serious adverse reactions reported to SHOT: 9 patients with pulmonary reactions (including 3 deaths possibly related to transfusion), and 4 patients with serious febrile, allergic or hypotensive reactions (all of whom recovered).

## DISCUSSION

The results of this large, randomised trial show that convalescent plasma did not improve survival or other clinical outcomes in patients hospitalised with COVID-19. The results were consistent across subgroups of age, sex, ethnicity, duration of symptoms prior to randomisation, level of respiratory support received at randomisation, and use of corticosteroids. Nine other randomised trials of convalescent plasma for the treatment of hospitalised patients with COVID-19 have been reported, which, together, have included fewer than 200 deaths.^9-17^ None of these trials have demonstrated a beneficial effect of convalescent plasma on mortality. Taking the results of all trials together, including RECOVERY (which is more than ten times larger than all other trials combined) allocation to convalescent plasma does not improve mortality (mortality RR 0·99, 95% CI 0·92–1·06, p=0·77 (figure 4).

It has been suggested that the benefits of convalescent plasma may depend on the transfused neutralising titre, and that using plasma with lower titres could explain negative results from previous randomised trials. In RECOVERY, all convalescent plasma was supplied via the UK National Blood Services using standardised laboratory processing. Convalescent donors were chosen based on high anti-spike IgG levels, using an ELISA that has been shown to correlate well with neutralising antibody.^23-25^ We used a EUROIMMUN S/CO ratio of ≥6 for plasma to qualify for use in this trial, which is substantially above the level of ≥3.5 that the US FDA recognises as high titre.^19^Recipients received plasma from two different donors to increase the chance that at least one contained higher levels of neutralising antibodies.

The presence of anti-SARS-CoV-2 antibodies in recipients prior to transfusion with convalescent plasma has also been cited as a possible reason for a lack of effect of convalescent plasma.^12^ In this trial we found that around 38% of patients were seronegative at randomisation and, although they had a markedly higher 28-day mortality risk than seropositive patients, we did not observe a survival benefit from convalescent plasma in these seronegative patients. There was, however, a suggestion of small improvements (proportional risk reduction of about one tenth) in the probability of successful discharge from hospital by day 28 and of progressing to invasive mechanical ventilation or death in seronegative patients allocated to convalescent plasma. The apparent heterogeneity in these secondary outcomes according to serostatus should be interpreted with a great deal of caution however, not least because (perhaps by chance or perhaps as a result of conscious or unconscious decisions about who to collect a serological sample from) the seronegative convalescent plasma recipients were slightly younger than the seronegative usual care group, whereas seropositive convalescent plasma recipients were slightly older than the seropositive usual care group (webtable 2). Due to the known strong effects of age on mortality risk, even these minor age imbalances could have led to a spuriously lower relative relative risk of death in the seronegative convalescent plasma recipients and a spuriously higher relative risk of death in the seropositive convalescent plasma recipients.

It has also been suggested that antibody based therapies are likely to be most effective in the early stages of COVID-19, when viral replication dominates.^26,^ We did not identify a benefit when we stratified by time since onset of illness in the main analysis (or in an exploratory analysis further subdividing time since illness onset). However, RECOVERY only included patients admitted to hospital and does not, therefore, address whether convalescent plasma may be of benefit if given early after SARS-CoV-2 infection and before the onset of significant disease.

Following randomisation to convalescent plasma, patients with hypoxia and a raised C-reactive protein (CRP ≥75mg/L) were eligible for a second randomisation to usual care versus usual care plus tocilizumab. Although a slightly lower proportion of patients allocated convalescent plasma subsequently received tocilizumab than patients allocated usual care (8% vs 10%, webtable 3), and although tocilizumab itself reduces 28-day mortality by around 15%,^1^ this difference in the likelihood of progression to the second randomisation is far too small to have had any material impact on our estimate of the effect of convalescent plasma on mortality (or other outcomes).

SARS-CoV-2 is an RNA virus with antigenic variability. The efficacy of convalescent plasma is likely to depend on the ‘match’ between the strain-specific transfused anti-SARS-CoV-2 antibodies in donor plasma and the infecting virus variant in the recipient. In December 2020 a new SARS-CoV-2 variant (B.1.1.7) was detected in the South East and East of England, with an earliest date of detection in September, which spread rapidly to become the dominant SARS-CoV-2 variant, in most regions of the UK, by January 2021.^27^ Whilst B.1.1.7 has changes in the spike glycoprotein that could theoretically modify antigenicity, only modest reductions in neutralisation by convalescent plasma have been reported.^28^ Consistent with this, we did not identify any evidence of a differential effect of convalescent plasma prior to and after the emergence of B.1.1.7 in the UK.^22^

During an epidemic caused by a novel virus, convalescent plasma is an appealing treatment as it may be available within weeks of the outbreak, long before other targeted therapies are available. Consequenty, convalescent plasma has been widely used for COVID-19 outside of clinical trials but, until now, there has been insufficient evidence from randomised trials to reliably assess its safety and efficacy.^8^ In RECOVERY, the largest clinical trial of convalescent plasma for any infectious indication, high-titre convalescent plasma did not improve survival or other prespecified clinical outcomes.

## Data Availability

The protocol, consent form, statistical analysis plan, definition & derivation of clinical characteristics & outcomes, training materials, regulatory documents, and other relevant study materials are available online at www.recoverytrial.net. As described in the protocol, the trial Steering Committee will facilitate the use of the study data and approval will not be unreasonably withheld. Deidentified participant data will be made available to bona fide researchers registered with an appropriate institution within 3 months of publication. However, the Steering Committee will need to be satisfied that any proposed publication is of high quality, honours the commitments made to the study participants in the consent documentation and ethical approvals, and is compliant with relevant legal and regulatory requirements (e.g. relating to data protection and privacy). The Steering Committee will have the right to review and comment on any draft manuscripts prior to publication.

https://www.ndph.ox.ac.uk/data-access

## Contributors

This manuscript was initially drafted by the PWH and MJL, further developed by the Writing Committee, and approved by all members of the trial steering committee. PWH and MJL vouch for the data and analyses, and for the fidelity of this report to the trial protocol and data anlysis plan. PWH, LE, LP, MM, JKB, LCC, SNF, TJ, KJ, WSL, AM, KR, EJ, DR, RH, and MJL designed the trial and trial protocol. MM, AR, G P-A, NB, TG, DZ, ST, NA, AU, JW, GK, TB, SS, RH, the Data Linkage team at the RECOVERY Coordinating Centre, Health Records, and Local Clinical Centre staff listed in the appendix collected the data. ES, NS, and JRE did the statistical analysis. LE, DR, and the blood and transfusion service staff listed in the appendix coordinated the collection and supply of convalescent plasma. SH ran the ELISA assays on patient samples. All authors contributed to data interpretation and critical review and revision of the manuscript. PWH and MJL had access to the trial data and had final responsibility for the decision to submit for publication.

## Writing Committee (on behalf of the RECOVERY Collaborative Group)

Professor Peter W Horby PhD FRCP,^a,*^ Lise Estcourt PhD FRCP FRCPath,^b,c,g,^* Leon Peto PhD,^a,d^* Professor Jonathan R Emberson PhD,^e,f^ Natalie Staplin PhD,^e,f,^ Enti Spata,^e,f^ Guilherme Pessoa-Amorim MD,^d,g^ Mark Cambell FRCPath,^a,d^ Alistair Roddick MBBS,^d,e^ Nigel J Brunskill,^h^ Tina George,^i^ Daniel Zehnder,^j^ Simon Tiberi,^k^ Ni Ni Aung,^l^ Alison Uriel,^m^ John Widdrington,^n^ George Koshy,^o^ Thomas Brown,^p^ Stephen Scott PhD,^q^ J Kenneth Baillie MD PhD,^r^ Professor Maya H Buch PhD FRCP,^s^ Professor Lucy C Chappell PhD,^t^ Professor Jeremy N Day PhD FRCP,^a,u^ Professor Saul N Faust PhD FRCPCH,^v^ Professor Thomas Jaki PhD,^w,x^ Katie Jeffery PhD FRCP FRCPath,^c^ Professor Edmund Juszczak MSc,^y^ Professor Wei Shen Lim DM,^y,z^ Marion Mafham MD,^c,†^ Professor Alan Montgomery PhD,^y^ Professor Andrew D Mumford PhD,^A^ Kathryn Rowan PhD,^B^ Professor Guy Thwaites PhD FRCP,^a,u^ Professor David J. Roberts PhD FRCPath^b,c,g†^ Professor Richard Haynes DM,^d,e,†^ Professor Martin J Landray PhD FRCP.^d,e,f,†^

^a^ Nuffield Department of Medicine, University of Oxford, Oxford, United Kingdom.

^b^ NHS Blood and Transplant Service, Oxford, United Kingdom

^c^ Radcliffe Department of Medicine, University of Oxford, Oxford, United Kingdom

^d^ Oxford University Hospitals NHS Foundation Trust, Oxford, United Kingdom

^e^ Nuffield Department of Population Health, University of Oxford, Oxford, United Kingdom

^f^ MRC Population Health Research Unit, University of Oxford, Oxford, United Kingdom

^g^ NIHR Oxford Biomedical Research Centre, Oxford University Hospitals NHS Foundation Trust, Oxford, United Kingdom

^h^ Department of Cardiovascular Science,, College of Life Sciences, University of Leicester, Leicester, United Kingdom

^i^ Basildon and Thurrock Hospitals NHS Foundation Trust, Basildon, United Kingdom

^j^ North Cumbria Integrated Care NHS Foundation Trust, Carlisle, United Kingdom

^k^ Barts Health NHS Foundation Trust, London, United Kingdom

^l^ North Tees & Hartlepool NHS Foundation Trust, Stockton-on-Tees, United Kingdom

^m^ North Manchester General Hospital, Pennine Acute Hospitals NHS Trust, Bury, United Kingdom

^n^ Centre for Clinical Infection, James Cook University Hospital, Middlesbrough, United Kingdom

^o^ North West Anglia NHS Foundation Trust, Peterborough, United Kingdom

^p^ Portsmouth Hospitals NHS Foundation Trust, Portsmouth, United Kingdom

^q^ The Countess of Chester Hospital NHS Foundation Trust, Chester, United Kingdom

^r^ Roslin Institute, University of Edinburgh, Edinburgh, United Kingdom

^s^ Centre for Musculoskeletal Research, University of Manchester, Manchester, and NIHR Manchester Biomedical Research Centre, United Kingdom.

^t^ School of Life Course Sciences, King’s College London, London, United Kingdom

^u^ Oxford University Clinical Research Unit, Ho Chi Minh City, Viet Nam

^v^ NIHR Southampton Clinical Research Facility and Biomedical Research Centre, University Hospital Southampton NHS Foundation Trust and University of Southampton, Southampton, United Kingdom

^w^ Department of Mathematics and Statistics, Lancaster University, Lancaster, United Kingdom

^x^ MRC Biostatistics Unit, University of Cambridge, Cambridge, United Kingdom

^y^ School of Medicine, University of Nottingham, Nottingham, United Kingdom

^z^ Respiratory Medicine Department, Nottingham University Hospitals NHS Trust, Nottingham, United Kingdom

^A^ School of Cellular and Molecular Medicine, University of Bristol, Bristol, United kingdom

^B^ Intensive Care National Audit & Research Centre, London, United Kingdom

*,^†^ equal contribution

## Data Monitoring Committee

Peter Sandercock, Janet Darbyshire, David DeMets, Robert Fowler, David Lalloo, Ian Roberts (until December 2020), Mohammed Munavvar (from January 2021), Janet Wittes.

## DATA SHARING

The protocol, consent form, statistical analysis plan, definition & derivation of clinical characteristics & outcomes, training materials, regulatory documents, and other relevant trial materials are available online at www.recoverytrial.net. As described in the protocol, the trial steering committee will facilitate the use of the trial data and approval will not be unreasonably withheld. Deidentified participant data will be made available to bona fide researchers registered with an appropriate institution within 3 months of publication. However, the steering committee will need to be satisfied that any proposed publication is of high quality, honours the commitments made to the trial patients in the consent documentation and ethical approvals, and is compliant with relevant legal and regulatory requirements (e.g. relating to data protection and privacy). The steering committee will have the right to review and comment on any draft manuscripts prior to publication. Data will be made available in line with the policy and procedures described at: https://www.ndph.ox.ac.uk/data-access. Those wishing to request access should complete the form at https://www.ndph.ox.ac.uk/files/about/data_access_enquiry_form_13_6_2019.docx and e-mailed to

## ACKNOWLEDGMENTS

Above all, we would like to thank the thousands of patients who participated in this trial. We would also like to thank the many doctors, nurses, pharmacists, other allied health professionals, and research administrators at 176 NHS hospital organisations across the whole of the UK, supported by staff at the National Institute of Health Research (NIHR) Clinical Research Network, NHS DigiTrials, NHS Blood and Transplant, the Scottish National Blood Transfusion Service, Welsh Blood Service, Northern Ireland Blood Transfusion Service, Public Health England, Department of Health & Social Care, the Intensive Care National Audit & Research Centre, Public Health Scotland, National Records Service of Scotland, the Secure Anonymised Information Linkage (SAIL) at University of Swansea, and the NHS in England, Scotland, Wales and Northern Ireland.

The RECOVERY trial is supported by a grant to the University of Oxford from UK Research and Innovation (UKRI)/NIHR (Grant reference: MC_PC_19056),by Department of Health and Social Care (DHSC)/UKRI/NIHR COVID-19 Rapid Response Grant (COV19-RECPLA). and by core funding provided by NIHR Oxford Biomedical Research Centre, Wellcome, the Bill and Melinda Gates Foundation, the Department for International Development, Health Data Research UK, the Medical Research Council Population Health Research Unit, the NIHR Health Protection Unit in Emerging and Zoonotic Infections, NHS Blood and Transplant Research and Development Funding, European Union’s Horizon 2020 research and innovation programme (SUPPORT-E - 101015756), and NIHR Clinical Trials Unit Support Funding. TJ is supported by a grant from UK Medical Research Council (MC_UU_0002/14) and an NIHR Senior Research Fellowship (NIHR-SRF-2015-08-001). WSL is supported by core funding provided by NIHR Nottingham Biomedical Research Centre. Abbvie contributed some supplies of lopinavir-ritonavir for use in this trial. Tocilizumab was provided free of charge for this trial by Roche Products Limited. REGN-COV2 was provided free of charge for this trial by Regeneron. The collection of plasma was funded by the DHSC through core funding and funding under COVID-19 and EU SoHo Grant.

The views expressed in this publication are those of the authors and not necessarily those of the NHS, the NIHR, NHS Blood and Transplant, the DHSC, or the EU.

