## Supplementary Appendix for "Convalescent plasma in patients admitted to hospital with COVID-19 (RECOVERY): a randomised, controlled, open-label, platform trial"

#### RECOVERY Collaborative Group

**Contents**

**Details of the RECOVERY Collaborative Group .................................................. 2**
**Supplementary Methods ........................................................................ 29**
Study organization 29
Collection and distribution of convalescent plasma 29
Measurement of participant baseline SARS-CoV-2 serostatus 29
Protocol changes 29
Main and second randomisation for adults 33
Supplementary statistical methods 34
Interim analyses: role of the Data Monitoring Committee 34
Ascertainment and classification of study outcomes 35
**Supplementary Tables ........................................................................ 50**
Webtable 1: Baseline characteristics of patients considered unsuitable for randomisation to convalescent plasma compared with those randomised to convalescent plasma versus usual care. 51
Webtable 2: Baseline characteristics by patient baseline antibody status and randomised allocation. 52
Webtable 3: Treatments given, by randomised allocation 53
Webtable 4: Cause-specific 28-day mortality 54
Webtable 5: Adverse events at 72 hours after randomisation 55
Webtable 6: Effect of allocation to convalescent plasma on major cardiac arrythmia 56
Webtable 7: Primary, secondary and subsidiary outcomes, after adjustment for the sex imbalance at randomisation 57
**Supplementary Figures ........................................................................ 58**
Webfigure 1: Mortality at 28 days, according to by retrospectively−defined categories of days since onset, respiratory support received and date of randomisation 59
Webfigure 2: Mortality at 28 days, according to patient antibody status 60
Webfigure 3: Discharged alive within 28 days, according to subgroup 61
Webfigure 4: Invasive mechanical ventilation or death, according to subgroup 62
**References ........................................................................ 63**
**Appendices ........................................................................ 64**
Appendix 1: RECOVERY Trial Protocol V12.1 65
Appendix 2: RECOVERY Trial Statistical Analysis Plan V2.1 105
Appendix 3: Definition and Derivation of Baseline Characteristics and Outcomes 131

### Details of the RECOVERY Collaborative Group

#### Writing Committee

PW Horby*, L Estcourt*, L Peto*, JR Emberson, N Staplin, E Spata, G Pessoa-Amorim MD, M Campbell, A Roddick, N Brunskill, T George, D Zehnder, S Tiberi, N Aung, A Uriel, J Widdrington, G Koshy, T Brown, S Scott, J Baillie, MH Buch, LC Chappell, J Day, SN Faust, T Jaki, K Jeffery, E Juszczak, WS Lim, M Mafham, A Montgomery, A Mumford, K Rowan, G Thwaites, DJ Roberts†, R Haynes†, MJ Landray†.

*,† equal contribution

#### Steering Committee

*Co-Chief Investigators* PW Horby, MJ Landray, *Members* JK Baillie, M Buch, L Chappell, J Day, SN Faust, R Haynes, T Jaki, K Jeffery, E Juszczak, WS Lim, M Mafham, A Montgomery, A Mumford, K Rowan, G Thwaites.

#### Data Monitoring Committee

P Sandercock (chair), J Darbyshire, D DeMets, R Fowler, D Lalloo, M Munavvar (from January 2020), I Roberts (until December 2020), J Wittes *Non-voting statisticians* J Emberson, N Staplin.

#### RECOVERY Trial Central Coordinating Office

*Co-Chief Investigators* P Horby, MJ Landray; *Clinical Trial Unit Lead* R Haynes; *Trial management* L Fletcher (coordinator), J Barton, A Basoglu, R Brown, W Brudlo, S Howard, G McChlery, K Taylor, E Denis; *Programming and validation* G Cui, B Goodenough, A King, M Lay, D Murray, W Stevens, K Wallendszus, R Welsh; *Data linkage* C Crichton, J Davies, R Goldacre, C Harper, F Knight, J Latham-Mollart, M Mafham, M Nunn, H Salih, J Welch; *Clinical support* M Campbell, G Pessoa-Amorim, L Peto, A Roddick; *Quality assurance* C Knott, J Wiles; *Statistics* JL Bell, J Emberson, E Juszczak, L Linsell, E Spata, N Staplin; *Communications* G Bagley, S Cameron, S Chamberlain, B Farrell, H Freeman, A Kennedy, A Whitehouse, S Wilkinson, C Wood; *Evidence synthesis (Vascular Overviews Group)* C Reith (coordinator) K Davies, H Halls, L Holland, K Wilson; *Administrative support* L Howie, M Lunn, P Rodgers

#### Convalescent Plasma Clinical Trials Unit (on behalf of the UK Blood Services)

L Estcourt (Lead), E Arbon, A Deary, F Clemons, N Dallas, A Evans, C Fitzpatrick-Creamer, C Foley, R Hodge, C Hudson, K Keen, E Laing, M Lobo-Clarke, J Mullings, R Paul, G Powter, S Shanmugaranjan, S Sweity and all staff at NHSBT Clinical Trials Unit that have worked on the Convalescent Plasma Project

#### Serious Hazards of Transfusion (SHOT)

S Carter-Graham, S Narayan, D Poles, V Tuckley on behalf of the SHOT Office team and all members of the SHOT Steering Group and Working Expert Group

#### UK Convalescent Plasma Steering Group

M Davies, K Ellis, S Hopkins, D Olszowka, J Palmer, M Pappa, J Sheffield, C Taylor, C Townsend

**National Health Service Blood Transfusion Services**

*NHS Blood and Transplant Plasma Board* M Banerjee, B Bassis, I Bateman, R Bradburn, R Bradbury, W Clark, A Clarkson, L Fahey, P Grealish, L Hontoria Del Hoyo, G Methven, R Moruza, D Ralphs, A Rashid, K Robinson, D Rose, S Scahill, M Stredder, *Clinical* A Deary, L Estcourt, A Evans, C Foley, E Laing, S MacLennan, O May, S Narayan, DJRoberts, G Powter, S Sweity, H Wilkinson, *Digital* B Ashford, G Bali, A Brazier, D Broggio, N Cardenas, S Cherif, K Chinn, R Dehulia, J Dhaliwal, T Elliott, J Fryer, I Millar, R Newman, N Salmons, A Simpson, M Zelman, *Communication* S Bailey, H Crumley, S Gilchrist, R Mistry, P Naga, S Tryon, B Willian, *Project Leadership* G Miflin (Lead) G Apostolides, L Banks, S Davison, G Gogarty, J Gregson, B Hall, E Hampson, R Joshi, K Loregnard, S Nazarali, D Ormrod, N Rylance, T Staincliffe, R Stewart, L Streeter, DJ Roberts *Planning* H Gray, J Morgan, R Lanaway, *Procurement* J Allen, R Athay-Hunt, D Kirkbride, J Marshall*,* J Murphy, P Phillips, S Taibo, W Thorne, *Donor Experience* N Akhtar, R Anand, K Anderson, S Ashgar, D Batty, E Bays, D Brack, S Brailsford, A Cooper, R Creighton, H Duggan, T Eve, E Hillerby, M Hinde, H Jones, I Lenchuk, M Prasad, W Rabin, J Towler, C Washington, V Whiteside, J Williams, *Plasma Collection* D Abdo, L Adams, L Allen, G Ashby, P Baughan, L Bayliss, J Blaxland, D Blofield, S Bradshaw, T Brittin, W Brown, S Butt, A Carlton, R Chavez, L Clifford, L Clifford, A Cotton, I Cron, D Cullen, A Curtis, J Dhalliwal, R Derbyshire, A Dhesi, M Dineen, S Duff-Miller, N Drogman, C Eaton, H Escreet, J Feely-Henderson, D Fox, G Franklin, D Franks, I Freestone, R Garmey, A Gibb, H Hanwell, L Hennen, SB Hesson, M Hill, G Hollis, D Jackson, S Jaggard, V Kara, M Kerr, D Latimer, C Leaver, L Lordache, L Maynard, J Meier, C Michelson, M Milloy, R Moir, I Moody, J Norgrave, T Noyce, A O'Brien, D Odumeru, C Peters, N Philips, E Richardson, L Roberts, D Robertson, F San Diego, A Sivyer, K Smith, H Spicer, G Stevens, C Tranter, A Turco, L Turner, R Turner, A West, R Wildman, CA Wood, L Wright, A Yeatman, *Development and Testing* D Affron, C Borhani, M Duley, D Edmondson, H Harvala, P Mellor, M Ray, J Sell, K Tettmar, N Watkins, *People* S Ahmed, J Benstead, G Bowyer, A Broderick, A Canabarro, S Dixon-Mould, K Flynn, M Harper, N Harris, E Hilton, I Hunter, C Ingall, S Kearley, H Lund, J McMahon, A Morgan, D Paul, E Poppitt, A Quirk, G Rees, R Shortland, H Templar, L Unitt, E Wiltsher, L Zarb, *Operational Optimization* B Allen, A Barber, P Baughan, K Beard, A Bennett, S Bolton, L Butler, D Bowen, P Daggett, J Davies, S Durgacharan, C Eaton-Howell, F Harper, A Hunter, A Issa, D McNamara, G Moore, D Richards, P Wright, W Zuurbier, *Quality and Regulatory* B Pelle, P Senior, J Kellington, F Murphy, U Sabat *Data Modelling (NHSBT & Bain)* G Airoldi, A Arroyo, B Bansal, P Bassi, C Brown, B Curtis, M Garstka, A Griffiths, R Johnson, T King, M Lawley, J Lawson, J Mehew, D Phillips, M Robb, S Shanmugaranjan, D Sood, R Tao, K Whitley, YM Wong *Finance* I Kirker, J Spence, and all staff at NHSBT who have worked on the convalescent plasma project. *NHS Wales Transfusion Service, Cardiff (WBS) Medical Director* J Birchall, *Clinical & Nursing* S Blackmore, K Kent, *Programme Lead* H Lovett, P Richardson *Component Development* M Briggs, C George, *Quality Assurance* M Cheadle, M Evans, *Manufacturing & Testing* G Stevens, Distribution & Hospital Liaison Lee Wong, and all staff at WBS who have worked on the convalescent plasma project.*Scottish National BloodTransfusion Service (SNBTS) Medical Director* M Turner, *Project Lead* M Carter, *Clinical* M Rowley, *Information Technology* T MacDonald, A Stewart, *Communication* C McGuigan, *Donation and Collection* S Armstrong-Fisher, L Mason, L McLintock, D McNaughton, N Priddee, R Evans, *Manufacturing and Testing* L Jarvis, D Turner, *Quality and Regulatory* E Hargreaves, E Stevenson, and all staff at SNBTS who have worked on the convalescent plasma project. *NHS Northern Ireland Transfusion Service (NIBTS) Medical Director* J Simpson, *CEO* K Jackson, *Project Lead* B Mullin, *Clinical and Nursing* K Maguire, P Mackey C Corry, J Paul, F Waller *Donation and Collection A Geddis, M Gillespie*, *Manufacturing and Testing* M Clarke, P Gowdy, M McCalmont, S Rainey *Quality and Regulatory* Angela Macauley, *CP Team Support* L Hadzik, I Gray, B Bradley, E Jefferson, and all staff at NIBTS who have worked on the convalescent plasma project.

**Sample reception, processing and storage laboratories**

*Nuffield Department of Surgical Sciences, University of Oxford, Oxford* S Cross, H McGivern, M Oliveira, R Ploeg, S Zyengi

*Radcliffe Department of Medicine, University of Oxford, Oxford* D Roberts (Lead), M Barbosa, M Boshir, N Ciccone, A Lamikanra, U Leuschner, S Pratheepkumar, A Roman, W Slack, A Townsend, HP Tsang, H Xiao,

**Antibody testing laboratories**

*Nuffield Department of Medicine, University of Oxford, Oxford* R Brown, S Cameron, D Crook, D Ebner, S Hatch, S Hoosdally, A Howarth, L Koukouflis, W Lason, B Marsden, L Martins Ferreira, H Pickford, P Tamblin-Hopper, M Wolna

**National Institute for Health Research Clinical Research Network**

*Coordinating Centre* A Barnard, J Beety, C Birch, M Brend, E Chambers, L Chappell, S Crawshaw, C Drake, H Duckles-Leech, J Graham, T Harman, H Harper, S Lock, K Lomme, N McMillan, I Nickson, U Ohia, E OKell, V Poustie**,** S Sam, P Sharratt, J Sheffield, H Slade, W Van't Hoff, S Walker, J Williamson; *Urgent Public Health Clinical Links* A De Soyza, P Dimitri, SN Faust, N Lemoine, J Minton; *East Midlands* K Gilmour, K Pearson *Eastern* C Armah, D Campbell, H Cate, A Priest, E Thomas, R Usher; *North East & North Cumbria* G Johnson, M Logan, S Pratt, A Price, K Shirley, E Walton, P Williams, F Yelnoorkar; *Kent, Surrey & Sussex* J Hanson, H Membrey, L Gill, A Oliver; *North West London* S Das, S Murphy, M Sutu; *Greater Manchester* J Collins, H Monaghan, A Unsworth, S Beddows; *North West Coast* K Barker-Williams, S Dowling, K Gibbons, K Pine; *North Thames* A Asghar, P Aubrey, D Beaumont-Jewell, K Donaldson, T Skinner; *South London* J Luo, N Mguni, N Muzengi, R Pleass, E Wayman; *South West Peninsula* A Coe, J Hicks, M Hough, C Levett, A Potter, J Taylor; *Thames Valley and South Midlands* M Dolman, L Gerdes, C Hall, T Lockett, D Porter *Wessex* J Bartholomew, L Dowden, C Rook, J Walters; *West of England* E Denton, H Tinkler; *Yorkshire & Humber* A Alexander, H Campbell, K Chapman, A Hall, A Rodgers; *West Midlands* P Boyle, M Brookes, C Callens, H Duffy, C Green, K Hampshire, S Harrison, J Kirk, M Naz, L Porter, P Ryan, J Shenton, J Warmington; *Devolved nations* M Amezaga, P Dicks, J Goodwin, H Hodgson, S Jackson, M Odam, D Williamson.

**Paediatric working group**

SN Faust (coordinator), A Bamford, S Bandi, J Bernatoniene, K Cathie, P Dmitri, S Drysdale, M Emonts, J Evans, A Finn, P Fleming, J Furness, C Gale, R Haynes, CE Jones, E Juszczak, D Jyotish, D Kelly, C Murray, N Pathan, L Pollock, A Ramanan, A Riordan, C Roehr, M Wan, E Whittaker.

**Obstetric working group**

L Chappell (coordinator), K Hodson, M Knight, S Pavord, C Williamson.

**Clinical support**

*NHS Lothian Out of Hours support line team* M Odam (coordinator), P Black, B Gallagher, L MacInnes, R O'Brien, K Priestley, A Saunderson; *Clinical Trial Service Unit Out of Hours clinical support* L Bowman, F Chen, R Clarke, M Goonasekara, R Haynes, W Herrington, P Judge, M Mafham, S Ng, D Preiss, C Reith, E Sammons, D Zhu.

**Health records**

*NHS DigiTrials, Southport* H Pinches, P Bowker, V Byrne-Watts, G Chapman, G Coleman, J Gray, A Rees, MJ Landray, M Mafham, N Mather, T Denwood; *Intensive Care National Audit & Research Centre, London* D Harrison; *National Records of Scotland* G Turner; *Public Health Scotland* J Bruce; *SAIL Databank, University of Swansea* C Arkley, S Rees.

**Convalescent plasma supply**

NHS Blood and Transplant (NHSBT), Northern Ireland Blood Transfusion Service (NIBTS), Scottish National Blood Transfusion Service (SNBTS), Welsh Blood Service (WBS).

**Local Clinical Centre RECOVERY trial staff**

(listed in descending order of the number of patients randomised per site)

**University Hospitals Of Leicester NHS Trust** C Brightling (PI), N Brunskill (Co-PI), M Wiselka (Co-PI), S Adenwalla, P Andreou, P Bakoulas, S Bandi, S Batham, T Beaver, K Bhandal, M Bourne, L Boyles, M Browett, M Cannon, A Charalambou, C Cheung, R Cotter, S Debbie, T Dinh, S Diver, A Dunphy, O Elneima, J Fawke, J Finch, Y Fozdar, C Gardiner-Hill, G Genato, A Ghattaoraya, M Graham-Brown, C Haines, B Hargadon, H Holdsworth, W Ibrahim, L Ingram, J Jesus Silva, K Kaul, M Joshi, S Kapoor, J Kim, A Kuverji, K Kyriaki, A Lea, T Lee, A Lewszuk, L Lock, K Macconaill, R Major, H McAuley, P McCourt, D Mullasseril Kutten, A Palfreeman, E Parker, K Patel, M Patterson, R Phillips, L Plummer, H Qureshi, R Russell, H Selvaskandan, S Southin, K Tsilimpari, C Wiesender, A Yousuf, UHL Blood Transfusion Laboratory Staff.

**North Cumbria Integrated Care NHS Foundation Trust** C Graham (PI), A Abdelaziz, O Ali, J Atkinson, G Bell, C Brewer, M Clapham, J Gregory, S Hanif, R Harper, M Lane, A McSkeane, J Nicholson, U Poultney, K Poulton, S Pritchard, S Shah, C Smit, P Tzavaras, J Sutton, V Vasadi, A Wilson, T Wilson, D Zehnder, the Blood Transfusion laboratory team.

**Basildon and Thurrock University Hospitals NHS Foundation Trust** K Thomas (PI), T George (Co-PI), E Cannon, J Cartwright, M Forsey, T Green, A Ikomi, L Kittridge, G Maloney, D Mukherjee, M Mushabe, A Nicholson, M O'Connell, K Pannu, M Pittman, J Riches, J Samuel, N Setty, A Solesbury, D Southam, M Vertue, K Wadsworth, B Yung, the Blood Transfusion laboratory team.

**Barts Health NHS Trust** S Tiberi (PI), H Abbass, A Aboaba, A Alam, M Al-Bayati, F Ali, R Allen, V Amosun, C Ardley, R Astin-Chamberlain, G Bacon, H Baillie, V Baker, S Begum, F Bibi, B Bloom, C Booth, M Browne, R Buchanan, M Buckland, N Caponi, C Chan, S Chandler, B Cipriano, H Dawe, M Dawkins, P Dias, S Elia, K El-Shakankery, D Fedorova (Associate PI), M Fernandez, A Fikree, R Flynn (Associate PI), D Gabriel, R Goiriz, P Goldsmith, M Griffiths, S Grigoriadou, K Gunganah, B Gurung, B Hack (Associate PI), P Hartridge, C Harwood, K Hashem, U Hemmila, G Heywood-Beldon, D Hobden, L Howaniec, G Huntington, I Hussain, S Iliodromiti, M Jawad, A Jayakumar, V Kapil, S Kelly, M Khan, H Khatun, H Kunst, J Lai (Associate PI), I Lancona-Malcolm, J Lancut, I Lee, D Lieberman, S Liebeschuetz, C Lindsay, O Lucey (Associate PI), K Majid, H Malcolm, C Maniero, H Marshall (Associate PI), T Martin, J Martin-Lazaro, N Matin, E McAleese, H McAleese, R McDermott, B McDonald (Associate PI), A Miah, I Milligan (Associate PI), A Mohammed, N Moramorell, S Naeem, T Newman, C Nic Fhogartaigh, L Noba, M Omar, N Omer, G Osoata, A Pakozdi, K Parkin, R Pearse, P Pfeffer, N Plaatjies, J Pott, J Powell, R Rachman (Associate PI), M Ramirez, W Ricketts, A Riddell, P Rughani, N Sahdev, F Santos, K Sarkar, F Seidu, B Selvarajah, M Shaikh, T Simangan, I Skene, K Smallshaw, C Suarez, T Swaine (Associate PI), S Thomas, N Thorn, C

Tierney, G Tunesi, C Tweed, R Uddin, S Ullah, L Velauthar, K Wiles, P Winter, T Wodehouse, P Woodland, H Yong, S Youssouf, the Blood Transfusion laboratory team.

**North Tees and Hartlepool NHS Foundation Trust** B Prudon (PI), V Jagannathan (Co-PI), A Al Aaraj (Associate PI), C Adams, D Barker, B Campbell, V Collins, D A Cox, J Deane, I Fenner, S Gowans, W Hartrey, M Nafei, L Poole, S Purvis, J Quigley, A Ramshaw, L Shepherd, R Srinivasan, R Taylor, M Walker, M Weetman, B Wetherill, S Wild, the Blood Transfusion laboratory team.

**Pennine Acute Hospitals NHS Trust** A Ustianowski (PI), J Raw (Co-PI), R Tully (Co-PI), K Abdusamad, O Akinwumiju, A Allameddine, S Andrews, Z Antonina, E Ayaz, B Blackledge, P Bradley, F Bray, M Bruce, C Carty, B Charles, G Connolly, C Corbett, J Cornwell, D Curley, S Dermody, E Falconer, L Fielding, J Flaherty, S Flynn, C Fox, D Hadfield, J Harris, L Hoggett, A Horsley, C Houghton, S Hussain, R Irving, P Jacob, D Johnstone, R Joseph, J Melville, P Kamath, T Khatun, T Lamb, H Law, M Lazo, G Lindergard, S Lokanathan, L Macfarlane, S Mathen, S McCullough, P McMaster, D McSorland, B Mishra, H Morris, S Munt, K Mushtaq, J Naisbitt, A Neal, R Newport, G O'Connor, C Okyne-Turkson, D O'Riordan, I Page, V Parambil, J Philbin, M Pinjala, C Porada, C Rishton, M Riste, J Rothwell, M Sam, Z Sarwar, L Scarratt, A Sengupta, H Sharaf, J Shaw, J Shaw (Associate PI), K Shepherd, A Slack, D Symon, H T-Michael, J Uttley, O Walton, S Williams, the Blood Transfusion laboratory teams.

**North West Anglia NHS Foundation Trust** K Rege (PI), R Buttery (PI), M Davies (PI), J Faccenda (PI), G Koshy (PI), B Abdelqader, R Abdul-Raheem, A Achara, Z Aftab, C Agbo, O Akindolie, O Aktinade, S Al-Juboori, F Allan, A Al-Rabahi, A Al Swaifi, R Ambrogetti, S Amin, K Aung, A Auwal, G Aviss, A Azman Shah, A Babs-Osibodu, K Bahadori, J Baker, T Baker, N Bakhtiar, T Bancroft, K Bandaru, A Bhanot, K Bhatt, J Bhayani, K Blaylock, T Bodenham, T Bond, H Boughton, K Bowen, H Bowyer, D Braganza, S Brooks, D Butcher, M Butt, N Butterworth-Cowin, A Cabandugama, J Carpenter, P Carter, A Catana, L Cave, R Chaube, M Cheung, Z Chikwanha , C Chisenga, S Choi, S Choudhury, V Christenssen, M Chukwu , H Cooper, C Cordell, D Corogeanu, A Cowan, R Croysdill, S Dahiya, J de Souza, S Diaz, B Donohue, N Duff, L Dufour, O Ebigbola, C Eddings, M El-Din, I Eletu, N Enachi, A Feroz, L Fieldhouse, L Finch, C Freer, P Gerard, E Gillham, A Gkoritsa, P Goodyear, R Gooentilleke, R Gosling, G Gosney, V Goss, S Goyal, J Hafsa, W Halford, S Havlik, D Hornan, Z Horne, T Hoskins, C Huson, A Idowu, I Iordanov, M Ishak, M Iyer, D Jafferji, E Jarnell, H Javed, E Johnson, A Jones, D Jones, J Jones, T Jones, E Kallistrou, J Kamara, R Kamath, S Kar, R Kark, N Kasianczuk, R Kassam, D Kaur, A Kerr, S Khalid, A Khan, A Khan, M Kheia, A King-Venables, F Kleemann, E Kopyj, A Kozak Eskenazia, R Kurian, S Lahane, M Lami, D Lamrani, K Leng, C Lippold, F Magezi, A Malik, J Marshall, R Matewe, F Maxton, N McCammon, K McDevitt, S McGuire, M Meek, P Mitchell, L Molloy, P Morzaria , Q Moyo, I Mugal, O Muraina, N Muru, A Mustafa, Y Myint, S Nazir, O Oderinde, I Okpala, N Olokoto, T Old, G Oleszkiewicz, H Orme, S O'Sullivan, P Paczko, S Pal, A Pandey, N Pao, S Parker, A Patel, S Pathak, S Peacock, B Peirce, T Perumpral, S Ponnusamy, S Pooboni, S Poon, J Porter, A Prasad, A Rahimi, J Rajkanna, M Rather, M Reece, R Renu Vattekkat, Z Rimba, S Rizvi, P Roberts, S Sahedra, S Saliu, M Samyraju, J Sanyal, V Saulite, R Schofield, S Schofield, K Scholes, S Shah, W Shah, S Shahad, P Sharma, C Sidaway, L Singh, P Sivasothy, E Smith, S Stacpoole, O Syed, B Tan, K Tan, J Taylor, N Temple, K Thazhatheyil, M Uddin, S Vardy, N Veale, A Waheed Adigun, D Walter, B Whelan, A White, G Williams, C Willshire, L Wilmott, K Winham-Whyte, M Wong, H Wroe, the Blood Transfusion laboratory teams.

**South Tees Hospitals NHS Foundation Trust** D Chadwick (PI), S Armstrong, D Athorne, M Branch, J Brolly, S Brown, C Carveth-Marshall, J Cheaveau, H Chen, Y Chua, N Cunningham, J Dodds, S Dorgan, P Dunn, D Dunn, C Elliott, P Harper, H Harwood, K Hebbron, F Hunt, A Kala Bhushan, P Lambert, D Leaning, T Manders, B McCarron, N Miller-Biot, C Milne, W Mohammad, M Mollet, A Murad, M Owston, J Potts, C Proctor, S Rao, M Seelarbokus, P

Singh, V Srirathan, L Swithenbank, L Thompson, H Wardy, L Wiblin, J Widdrington, J Williams, P Winder, C Wroe, the Blood Transfusion laboratory team.

**Portsmouth Hospitals NHS Trust** T Brown (PI), J Andrews, R Baker, M Baker-Moffatt, A Bamgboye, D Barnes, S Baryschpolec, S Begum, L Bell, H Blackman, J Borbone, N Borman, L Brimfield, M Broadway, F Brogan, R Bungue-Tuble, K Burrows, A Chauhan, E Cowan, M Czekaj, Z Daly, A Darbyshire, M Davey, M David, A Davies, J Denham, C Edwards, S Elliott, H Evison, L Fox, Z Garner, I Gedge, B Giles, S Glaysher, S Gosling, A Gribbin, J Hale, Y Harrington-Davies, L Hawes, J Hayward, K Heron, A Hicks, A Holmes, S Howe, B Jones, C Lameirinhas, B Longhurst, M Mamman, J Marshall, G Matthias, S McCready, L Milne, C Minnis, M Moon, J Mouland, L Murray, C Pereira Dias Alves, L Richmond, D Rodgers, S Rose, M Rowley, M Rutgers, T Scorrer, L Shayler, S Smith, C Stemp, E Taylor, R Thornton, A Tiller, C Turner, L Vinall, M Wands, J Warren, L Watkins, M White, L Wiffen, C Wong, K Wren, Y Yang, L Wiffen (Associate PI), J Winter, the Blood Transfusion laboratory team.

**Countess Of Chester Hospital NHS Foundation Trust** S Scott (PI), A Abdulakeem, M Abouibrahim, S Ahmed, M Ahmed, S Ahmed, A Ajibode, L Alomari, A Asrar, E Austin, A Awan, P Bamford, A Banerjee, A Barclay, L Barker, K Barker-Williams, T Barnes, W Barnsley, H Batty, F Bawa, G Beech, J Bellamu, K Bennion, I Benton, S Billingham, K Blythe, S Brearey, S Brigham, V Brooker, C Burchett, M Burgess, R Cade, F Cameron, R Cannan, M Cardwell, K Cawley, N Chavasse, L Cheater, Z Cheng, K Clark, R Clarke, R Clifford, E Cole, C Cook, C Cotton, S Cremona, D Curran, H Dallow, A Davidson, A Davies, A Davies, K Davis, Y Doi, E Doke, C Dragos, L Eagles, M Edwards, L Ellerton, H Elmasry, E Entwistle, A Fallow, M Faulkner, E Ferrelly, K Flewitt, C Foden, D Font, R Gale, L Gamble, M Gardiner, K Gillespie, L Goodfellow, M Grant, S Griffiths, Y Griffiths, J Grounds, V Guratsky, L Hartley, P Hayle, C Haylett, N Henry, P Hettiarachchi, L Hill, H Hodgkins, L Hodgkinson, S Holt, S Hunt, M Ibraheim, M Irshad, M Iyer, Z Jamal, R James, A Jayachandran, R Jayaram, H Jeffrey, A Johari, O Johnson, C Jones, C Jones, R Jones, A Kazeem, N Kearsley, K Kouranloo, S Leason, N Lightfoot, B Lim, D Llanera, R Llewellyn, S Loh, E London, C Lonsdale, T Maheswaran, G Markham, E Martin, P Maskell, A Mather, A Mathew, I Mazen, M McBuigan, J McBurney, M McCarthy, R McEwen, E Meeks, G Metcalf-Cuenca, L Michalca-Mason, S Middleton, L Mihalca-Mason, A Moore, L Morris, K Murray, I Nagra, A Nallasivan, M Nayyar, F Naz, R Nelson, C Oakley, E Okpo (Associate PI), F Olonipile, C O'Neill, C Ord, j Osman, E Owens, S Pajak, S Pal, J Parker, H Parry, S Pearson, K Penney, E Perritt, D Phillips, C Pickering, A Ponnuswamy, V Prescott, J Prince, S Rahman, S Read, J Reed, A Reid (Associate PI), J Roberts, N Robin, R Robinson, M Saladi, D Scholes, A Scott, H Shabbir, M Shah, T Spencer, J Spriggs, C Steele, N Swarbrick, Z Syrimi, L Theocharidou, M Thevendra, C Thorne, A Timmis, S Tomlin, T Trussell, O Vass, B Vlies, A Walters, L Walton, K Webb, T Webster, S Wheaver, D Whitmore, C Wyatt, W Yap, E Young, L Zammit, E Zin, the Blood Transfusion laboratory team.

**Nottingham University Hospitals NHS Trust** W Lim (PI), M Ali, L Allen, A Andrews, L Anderson, S Ashraf, D Ashton, G Babington, G Bartlett, D Batra, L Bendall, N Benetti, H Bond, T Brear, A Buck, G Bugg, J Butler, J Butler, R Cammack, J Cantliff, C Chang, L Clark, E Connor, P Davies, M Dent, C Dobson, M Fatemi, A Fatemi, L Fleming, J Grundy, J Hallas, L Hodgen, S Hodgson, S Hodgkinson, N Horton-Turner, L Howard, C Hutchinson, B Jackson, J Kaur, E Keddie-Gray, C Khurana, M Langley, M Langley, L Looby, M Meredith, M Meredith, L Morris, H Navarra, R Nicol, J Oliver, C Peters, B Petrova, S Robinson, R Rose, Z Rose, L Ryan, M I Saez-Garcia-Holloway, J Sampson, M Skill, S Springworth, G Squires, J Squires, R Taylor, A Tervit, A Thomas, J Thornton, K Topham, S Warburton, S Wardle, H Waterfall, S Wei, T Wildsmith, C Wilkes, L Wilson, the Blood Transfusion laboratory team.

**Northampton General Hospital NHS Trust** E Elmahi (PI), M Zaman (PI), B Abdul, A Abdulmumeen, N Abdulshukkoor, A Adnan, M Ahammed Nazeer, A Bazli, N Benesh, R Bisa, W Chin, M Comery, N Cunningham, H Daggett, T Darling, E Davies, H Enyi, S Fawohunre, N

Geoghegan, J Glover, J Hague, K Hall, C Hallett, K Haresh, W Ul Hassan, J Hewertson, J Hosea, N Htoon, M Idrees, C Igwe, H Imtiaz, M Irshad, A Ismail, R Jeffrey, J Jith, P Joshi, R Kaliannan Periyasami, A Khalid, M Khalid, R Kodituwakku, P Lopez, A Mahmood, M Malanca, I Mapfunde, V Maruthamuthu, S Masood, M Matharu, A Merchant, F Merchant, S Naqvi, N Natarajan, R Natarajan, K Nawaz, O Ndefo, O Ogunkeye, S Paranamana, N Pugh, A Raj, K Rashid, M Rogers, M Saad, G Selvadurai, A Shah, M Shahzeb, N Shrestha, A Singh, K Smith, B Sohail, M Spinks, K Spreckley, L Stockham, A Takyi, Y He Teoh, S Ullah, H Vayalaman, S Wafa, T Ward, R Watson, D White, L Ylquimiche Melly, R Zulaikha, the Blood Transfusion laboratory team.

**Calderdale and Huddersfield NHS Foundation Trust** P Desai (PI), D Appleyard, D Bromley, N Chambers, S Dale, P Desai, L Gledhill, J Goddard, J Greig, K Hallas, K Hanson, K Holroyd, M Home, D Kelly, S Kershaw, M Lake, A Maharajh, L Matapure, S Mellor, E Merwaha (Associate PI), H Riley, M Robinson, K Sandhu, K Schwarz, H Senior, L Shaw, L Terrett, M Thompson (Associate PI), M Usher, A Wilson, T Wood, the Blood Transfusion laboratory team.

**Liverpool University Hospitals NHS Foundation Trust** P Hine (PI), P Albert (Co-PI), S Todd (Co-PI), I Welters (Co-PI), D Wootton (Co-PI), M Ahmed, R Ahmed, A Al Balushi, L Alomari, P Banks, D Barr, J Bassett, J Bell, A Bennett, H Bond, T Brankin-Frisby, G Bretland, M Brodsky, P Burgess, P Burgett, C Burston, J Byrne, S Casey, L Chambers, D Coey, T Cross, J Cruise, J Currie, L Dobbie, R Downey, C Dragomir, A Du Thinh, G Duncan, J Early, K Fenlon, H Frankland, J Gorry, S Gould, A Gureviciute, K Haigh, L Hampson, A Hanson, M Harrison, P Hazenberg, S Hicks, S Hope, I Houghton, K Hunter, A Islim, F Jaime, L Keogan, S King, K Knowles, K Krasauskas, J Lewis, M Lofthouse, P Lopez, C Lowe, Z Mahmood, F Malein (Associate PI), K Martin, A Mediana, Z Mellor, B Metcalfe, M Middleton, K Monsell, N Nicholas, R Osanlou, L Pauls, A Peacock, C Prince, S Pringle, L Rigby, M Riley, A Rowe (Associate PI), M Samuel, D Scanlon, J Sedano, D Shaw, C Smith (Associate PI), S Stevenson, A Stockdale, J Tempany, C Toohey, R Watson, V Waugh, R Westhead, K Williams, H Wissett, A Wood, R Wright, the lab BMS and MLA teams.

**Wirral University Teaching Hospital NHS Foundation Trust** A Wight (PI), L Bailey, S Bokhari, S Brownlee, A Bull, S Carter, J Corless, L Delaney, C Denmade, N Ellard, A Farrell, A Hufton, R Jacob, K Jones, H Kerss, J McEntee, N Morris, R Myagerimath, T Newcombe, M Parsonage, H Peake, D Pearson, R Penfold, S Rath, R Saunders, A Sharp, B Spencer, A Suliman, S Sutton, H Tan, D Tarpey, L Thompson, T Thornton, E Twohey, D Wagstaff, Z Wahbi, S Williams, D Woods, the Blood Transfusion laboratory team.

**Mid Cheshire Hospitals NHS Foundation Trust** D Maseda (PI), C Ball, K Best, G Bridgwood, R Broadhurst, C Brockelsby, T Brockley, J Brown, R Bujazia, S Clarke, J Cremona, C Dixon, S Dowson, H Drogan, F Duncan, M Emms, H Farooq, D Fullerton, N G, C Gabriel, S Hammersley, R Hum, T Jones, S Kay, E Kelly, M Kidd, D Lees, R Lowsby, E Matovu, K McIntyre, H Moulton, K Nourein, K Pagett, A Ritchings, S Smith, A Steel, J Taylor, K Thomas, K Turbitt, S Yasmin, the Blood Transfusion laboratory team.

**Epsom and St Helier University Hospitals NHS Trust** S Winn (PI), T Evans (Co-PI), R Wake (Co-PI), F Abbas, S Acheampong, S Ahamed Sadiq, A Aldana, B Al-Hakim, P Allen, G Azzopardi, E Balogh, Z Baxter, C Bond, S Burrage, C Cheeld, R Chicano, I Chukwulobelu, N Colbeck, R Dogra, E Doherty, A Elradi, J Emberton, L Evans, R Ganapathy, N Hamilton, J Handford, M Haque, J Hayes, R Hayre, D Howlett, L Ilves, C Jagadish, S Jain, K Jian, L Johnson, A Johnson, B Kim, J Kotecha, A Kundu, D Langer, E Lester, A Lunia, Y Mashhoudi, K Mathias, T Medveczky, F Mellor, G Mic, S Moore, M Morgan, O Mushtaq, H Nabakka, S Nafees, A Oluwole-Ojo, H Omuco, V Palagiri Sai, M Phanish, H Rahimi, S Ramanna, J Ratoff, M Ridha, S Rozewicz, T Samuel, G Saxena, S Selvendran, B Shah, S Shahnazari, R Shail, A Sharif, A Short, S Singh, S Somalanka, R Suckling, R Suri, P Swift, L Tarrant, V Tyagi, N Vilimiene, P Virdee, L Walsh, R Zuha, the Blood Transfusion laboratory staff.

**Medway NHS Foundation Trust** R Sarkar (PI), C Adams, I Ahmed, S Ahmed, F Aliyuda, S Ambler, S Arnott, A Arya, F Babatunde, A Bhandari, N Bhatia, L Brassington, F Brokke, D Bruce, B Cassimon, R Chauhan, V Chawla, R Chineka, A Davis, C Donnelly, N Edmond, M Elbeshy, C Froneman, C Gnanalingam, D Gotham, T Gower, S Haskins, W Hassan, M Hollands, S Jansz, B Josiah, M Kamara, S Kidney, T Kyere-Diabour, K Lewiston, A Liao, A Maheswaran, N Miah, S Millington, L Mires, A Mitchell, C Mizzi, K Naicker, R Nicholas, Z Nurgat, I Petrou, M Phulpoto, Z Rehman, A Ross-Parker, A Roy, A Ryan, Y Samuel, E Samuels, A Shaibu, A Sharma, P Soor, J Sporrer, R Squires, W Ul Hassan, P Vankayalapati, B Velan, E Vyras, J Walker, A Williams, J Wood, N Zuhra (Associate PI), the Blood Transfusion laboratory team.

**University Hospitals Bristol and Weston NHS Foundation Trust** J Willis (PI), N Blencowe (Co-PI), P Singhal (Co-PI), M Abraham, B Al-Ramadhani, A Archer, G Aziz, A Balcombe, K Bateman, M Baxter, A Archer (Associate PI), L Beacham, K Belfield (Associate PI), N Bell, M Beresford, J Bernatoniene, D Bhojwani, S Biggs, J Blazeby, K Bobruk, N Brown, L Buckley, P Butler, C Caws, E Chakkarapani, K Chatar, B Chivima, C Clemente de la Torre, K Cobain, S Cooke, D Cotterill, E Courtney, R Cowburn, S Cowman, K Coy, H Crosby, K Curtis, P Davis, O Drewett, H Dymond, K Edgerley, M Ekoi, M Elokl, B Evans, T Farmery, N Fineman, A Finn, L Gamble, F Garty, B Gibbison, S Goolam-Hossen, L Gourbault, D Grant, K Gregory, M Griffin, R Groome, M Hamdollah-Zadeh, A Hannington, J Heywood, A Hindmarsh, N Holling, R Houlihan, J Hrycaiczuk, H Hudson, K Hurley, R Jarvis, B Jeffs, A Jones, R E Jones, L Johnston, E King-Oakley, E Kirkham, R Kumar, M Kurdy, L Kyle (Associate PI), S Lang, L Leandro, H Legge, F Loro, A Low, H Martin, L McCullagh, G McMahon, L Millett, K Millington, J Mok, J Moon, L Morgan, S Mulligan, C O'Donovan, A Parejasanchez, E Payne, C Penman, J Pickard, C Plumptre, A Ramanan, J Ramirez, S Ratcliffe, J Robinson, M Roderick, S Scattergood, A Schadenberg, R Sheppeard, C Shioi, D Simpson, A Skorko, R Squires, M Stuttard, P Sugden, S Sundar (Associate PI), V Stefania, T Swart, E Swift, K Thompson, K Turner, S Turner, A Tyer, S Vergnano, R Vincent, R Ward, A Wardle, S White, S Wilkinson, J Williams, S Williams, H Winter, L Woollen, R Wright, A Younes Ibrahim, the Blood Transfusion laboratory teams.

**Barking, Havering and Redbridge University Hospitals NHS Trust** M Phull (PI), A Umaipalan (Co-PI), A Misbahuddin (Co-I), A Abdulle, A Asquith, C Baldwin, S Balkee, A Basumatary, S Bates, H Chandler, A Davison, P Dugh, K Dunne, K Fielder, R Gad, P Greaves, W Halder, J Hastings, A Holman, K Hunt, V Katsande, S Lockyer, A Loverdou, G Marange, A McGregor, A Minhas, A Minogue, A Mohamed, N Munu, H Neils, D Nicholls, N O'Brien, N Page, L Parker, P Patel, T Pogreban, L Rosaroso, S Sagrir, E Salciute, C Sanderson, M Sharma, G Siame, H Smith, X Tang, E Visentin.

**University Hospitals Birmingham NHS Foundation Trust** C Green (PI), T Whitehouse (Co-PI), D Dosanjh, I Ahmed, N Anderson, C Armstrong, A Bamford, H Bancroft, M Bates, M Bellamy, C Bergin, K Bhandal, N Cianci (Associate PI), K Clay (Associate PI), L Cooper, J Daglish, J Dasgin, A Desai, S Dhani, E Forster, E Grobovaite, B Hopkins, D Hull, Y Hussain, J Jones, M Lovell, D Lynch, C McGhee, C McNeill, F Moore, A Nilsson, J Nunnick, W Osborne (Associate PI), D Parekh, C Prest, V Price, M Sangombe, H Smith, I Storey, L Thrasyvoulou, K Tsakiridou, D Walsh, S Welch, H Willis, K Wood, L Wood, J Woodford, G Wooldridge, C Zullo, the Blood Transfusion laboratory teams.

**Manchester University NHS Foundation Trust** T Felton (PI), M Abbas, A Abdul Rasheed, T Abraham, S Aggarwal, A Ahmed, S Akili, P Alexander, A Allanson, E Anyanwu, B Al-Sheklly, D Arora, M Avery, C Avram, A Aya, J Banda, H Banks, M Baptist, M Barrera, R Bazaz, R Behrouzi, V Benson, A Bentley, A Bhadi, A Biju, A Bikov, K Birchall, S Blane, S Bokhari, P Bradley, J Bradley-Potts, J Bright, R Brown, S Burgess, G Calisti, S Carley, N Chaudhuri, S Chilcott, C Chmiel, E Church, R Clark, J Clayton-Smith, R Conway, E Cook, E Cooperwaite,

G Cummings-Fosong, S Currie, H Dalgleish, C Davies, K Dean, A Desai, R Dhillon, D Dolan, G Donohoe, A Duggan, B Duran, H Durrington, C Eades, R Eatough, S Elyoussfi, G Evans, M Evans, A Fairclough, D Faluyi, S Ferguson, J Fielding, S Fiouni, G Fogarty, S Fowler, S Giannopoulou, R Gillott, A Gipson, S Glasgow, T Gorsuch, G Grana, A Grayson, G Grey, B Griffin, P Hackney, B Hameed, I Hamid, S Handrean, J Henry, L Higgins, L Holt, A Horsley, L Howard, S Hughes, A Hulme, P Hulme, A Hussain, M Hyslop, J Ingham, O Ismail, A Jafar, S Jamal, L James, A John, E Johnstone, N Karunaratne, Z Kausar, J Kayappurathu, R Kelly, A Khan, S Khan, W Khan, S Knight, E Kolakaluri, C Kosmidis, E Kothandaraman, S Krizak, N Kyi, F Lalloo, C Logue, R Lord, L Macfarlane, A Madden, A Mahaveer, L Manderson, G Margaritopoulos, P Marsden, J Mathews, E McCarthy, B McGrath, C Mendonca, A Metryka, D Micallef, A Mishra, H Mistry, S Mitra, S Moss, A Muazzam, D Mudawi, C Murray, M Naguib, S Naveed, P Ninan, M Nirmalan, R Norton, N Odell, R Osborne, G Padden, A Palacios, A Panes, C Pantin, B Parker, C Parker, L Peacock, A Peasley, S Pendlebury, J Peters, M PI, F Pomery, J Potts, N Power, M Pursell, A Rasheed, S Ratcliffe, M Reilly, C Reynard, E Rice, M Rice, P Riley, P Rivera Ortega, J Rogers, T Rogers, R Santosh, A Scott, K Sellers, N Sen, T Shanahan, A Shawcross, S Shibly, A Simpson, S Sivanadarajah, N Skehan, J Smith, C Smith, L Smith, J Soren, W Spiller, J Stratton, A Stubbs, K Swist-Szulik, B Tallon, S Thomas, S Thorpe, M Tohfa, R Tousis, T Trimble, T Turgut, M Varghese, G Varnier, I Venables, S Vinay, R Wang, L Ward, C Warner, E Watson, D Watterson, L Wentworth, C Whitehead, D Wilcock, J Williams, E Willis, L Woodhead, S Worton, B Xavier, the Blood Transfusion laboratory teams.

**Luton and Dunstable University Hospital NHS Foundation Trust** D Shaw (PI), S Tariq (Co-PI), N Ahmed, C Alford, S Ali, S Allen, M Alzetani, C Ambrose, K Aneke, T Angel, Z Aung, R Banerjee, T Baqai, A Batla, M Bergstrom, S Bhakta, N Bibi, T Chapman, J Dijo, L Dirmantaite, M Edmondson, E Elfar, M Elgamal, H El-Sbahi, D Fisher, D Fishman, C Fornolles, T Forshall, A Francioni, N Gasmalseed, S Gent, A Gouveia, L Hunold, R Hussein, A Ibrahim, A Ingram, T Jacques- Brown, R James, D Joshi, K Kabiru Dawa, F Khan, M Khan, S Khan, S Lee, C Lingam, C Luximon, N Marcus, M Masood, R Mejri, G Mmadubuko, A Moharram, C Moss, T Mukwa, G Naik, A Ng, L Nicholls, M Nisar, J Nnadi, M Parmar, V Parmar, F Prasanth Raj, V Puisa, V Quick, B Ramabhadran, A Reddy, N Riaz, B Rudran, S Sabaretnam, H Sagoo, S Sarma, K Savlani, P Shah, S Soo, P Sothirajah, I Southern, E Strakosch, J Tan, M Tate, C Travill, V Uppal, W Wakeford, the Blood Bank team.

**Milton Keynes University Hospital NHS Foundation Trust** R Stewart (PI), J Bae (Co-PI), S Bowman (Co-PI), A Chakraborty (Co-PI), S Fox (Co-PI), L How (Co-PI), D Mital (Co-PI), L Anguvaa, G Bega, J Beharry, E Clare, N Jacob, M Khan, C Lowe, J Mead, L Mew, L Moran, E Mwaura, M Nathvani, T Perry, A Rose, D Scaletta, S Shah, L Siamia, O Spring, S Velankar, L Wren, F Wright, the Blood Transfusion laboratory team.

**East Suffolk and North Essex NHS Foundation Trust** V Kushakovsky (PI), M Ramali (Co-PI), K Ahmed, S Alam, B Atraskiewicz, J Bailey, I Balluz, D Beeby, S Bell, J Bloomfield, N Broughton, C Buckman, M Burton, E Byworth, C Calver, J Campbell, P Carroll, C Chabo, R Chalmers, M Chowdhury, K Cooke, N Deole, T Dinh (Associate PI), J Douse, C Driscoll, S Finbow, I Floodgate, R Francis, E Galloway, C Galloway, M Garfield, A Ghosh, G Gray, P Greenfield, A Gribble, M Gunawardena, M Hadjiandreou, H Hewer, M Hossain, R Howard-Griffin, K Howlett, C Huah, L James, A Islam, Z Jiao, K Johannessen, J Kathirgamachelvam, S Lee, R Lewis, R Lloyd, L Mabelin, P Mallett, M Mohan, D Morris, S Nallapareddy, S Nishat, R Osagie, H Ooi (Associate PI), H Patel, A Pereira, M Pretorius, B Purewal, F Ramali, H Rawlins, P Ridley, V Rivers, J Rosier, S Sharma, A Sharma, A Sheik, R Skelly, C Smith, R Smith, R Smith, R Sreenivasan, P Tovey, K Turner, A Turner, S Turner, K Vithian, I Weichert, W Win, the Blood Transfusion laboratory team.

**Royal Free London NHS Foundation Trust** B Caplin (PI), H Tahir (Co-PI), R Abdul-Kadir, I Alshaer, M Anderson, R Atugonza, G Badhan, S Bhagani, B Bobie, I Carboo, E Cheung, O Coker, V Conteh, E Damian, R Davies, N Davies, V Deelchand, L Ehiorobo, A M Ellis, C Ellis,

P Gardiner, H Hughes, C Jarvis, V Jennings, V Krishnamurthy, A Kurani (Associate PI), L Lamb, S Lee, A Li, J Li, H Mahdi, D Matila, S Mellin, R Moores, J Morales, A Nandani, S Nasa, A Natarajan, S O'Farrell, A Oomatia, A Osadcow, J Osei-Bobie, P Patel, C Patterson, R Rankhelawon, A Rodger, S Sithiravel, T Sobande, P Talbot, S Tekle, G Wallis, E Witele, the Blood Transfusion laboratory team.

**University Hospitals Of Derby and Burton NHS Foundation Trust** T Bewick (PI), P Daniel (Co-PI), U Nanda (Co-PI), H Ahmad, G Bell, J Buchan, H Clarke, C Downes, K English, A Fletcher, J Hampson, M Hayman, N Jackson, J Jeyachandran, K Kacinova, S Kaur, A Matthews, S Ohja, L Prince, J Radford, K Riches, G Robinson, A Sathyanarayanan, R Shiers, L Wilcox, L Wright, the Blood Transfusion laboratory teams.

**South Tyneside and Sunderland NHS Foundation Trust** L Fuller (PI), A MacNair (Co-PI), M Rangar (Co-PI), C Brown, A Burns, C Caroline, J Caulfield, R Davidson, N Elkaram, I Emmerson, L Fairlie, M Hashimm, J Henderson, K Hinshaw, R Hovvels, K Martin, S Matthews, M McKee, J Moore, N Mullen, P Murphy, L Palmer, G Parish, P Patterson, L Smith, A Smith, L Sudlow, L Terry, J Trattles, A Trotter, F Wakinshaw, E Walton, the Blood Transfusion laboratory teams.

**Wrightington, Wigan and Leigh NHS Foundation Trust** A Ashish (PI), J Cooper, J Cooper, D Heaton, S Hough, L McCreery, C Moore, V Parkinson, E Robinson, T Taylor, C Tierney, A Verma, N Waddington, J Wesson, C Williams, C Zipitis, the Blood Transfusion laboratory team.

**Surrey and Sussex Healthcare NHS Trust** E Potton (PI), N Jain (Sub-I), A Khadar (Sub-I), P Morgan (Sub-I), J Penny (Sub-I), E Tatam (Sub-I), S Abbasi, D Acharya, A Acosta, L Ahmed, T Akbar, S Ali, M Alkhusheh, V Amosun, A Arter, M Babi, J Bacon, K Bailey, N Balachandran, S Bandyopadhyam, L Banks, J Barla, T Batty, S Bax, A Belgaumkar, G Benison-Horner, A Boles, N Broomhead, E Cetti, C Chan, I Chaudhry, D Chudgar, J Clark, S Clueit, S Collins, E Combes, G Conway, O Curtis, M Das, M Daschel, S Davies, A Day, M Dhar, K Diaz-Pratt, C Dragan, H Dube, V Duraiswamy, J Elias, A Ellis, T-Y Ellis, J Emmanuel, A Engden, Y Fahmay, B Field, K Fishwick, U Ganesh, C Gilbert, T Giokanini-Royal, E Goudie, S Griffith, S Gurung, R Habibi, C Halevy, A Haqiqi, R Hartley, A Hayman, J Hives, M Horsford, S Hughes, C Hui, R Hussain, C Iles, L Jackson, N Jain, A James, D Jayaram, E Jessup-Dunton, T Joefield, N Khan, W Kieffer, E Knox, V Kumar, R Kumar, V Kurmars, H Lafferty, F Lamb, R Layug, N Leitch, J Lemos, W Lim, U Limbu, R Loveless, M Mackenzie, N Maghsoodi, S Maher, M Maljk, I Man, N McCarthy, B Mearns, C Mearns, K Morgan-Jones, G Mortem, G Morton, B Moya, G Murphy, S Mutton, A Myers, T Nasser, J Navaratnam, S Nazir, S Nepal, K Nimako, L Nimako, C O'Connor, A Patel, K Patel, J Penny, V Phongsathorn, PA Pillai, M Poole, N Qureshi, S Ranjan, A Rehman, R Rook, T Samuels, E Scott, G Sekadde, A Sharma, G Sharp, S Shotton, O Simmons, P Singh, S Smith, K Sri Paranthamen, S Suresh, A Tharmakulasingam, K Thevarajah, L Thomas, H Timms, N Tomasova, S Tucker, S Vara, C Vaz, S Weller, J White, M Wilde, I Wilkinson, C Williams, M Win, D Woosey, D Wright, the laboratory team.

**East Kent Hospitals University NHS Foundation Trust** N Richardson (PI), A Alegria (Co-PI), R Kapoor (Co-PI), K Adegoke, L Allen, E Beranova, H Blackgrove, G Boehmer, L Boorman, P Christian, T Cosier, N Crisp, T Curtis, J Davis, J Deery, A Elgohary, T Elsefi, A Gillian, C Hargreaves, T Hazelton, G Hector, R Hulbert, A Ionita, J Jeffrey, R Jennings, R Kapoor, A Knight, K Kolsteren, C Linares, D Loader, C Lorenzen, S Lymn, L March, E Matisa, K Mears, J McAndrew, S Mitchell, M Montasser, A Moon, H Moore, D Murdoch, C Oboh, S O’Brien, P Offord, S Parashar (Associate PI), C Pelham, F Perkins, C Price, A Rajasri, J Rand, B Rayner, S Rogers, N Schumacher, R Solly, R Spicer, D Stephensen, S Stirrup, L Tague, S Tilbey, F Turner, S Turney, V Vasu, M Venditti, R Vernall, H Weston, Z Woodward, the Blood Transfusion laboratory team.

**Aneurin Bevan University LHB** T Szakmany (PI), L Aitken, K Arora, E Baker, M Brouns, J Cann, S Cherian, S Cutler, C Davies, A Dell, C Dunn (Associate PI), N Duric (Associate PI), M Edwards, S El Behery, S Fairbairn, S Goyal, A Griffiths, J Harris, N Hawkins, E Heron, J Hickey, M Hoare, S Hodge, G Hodkinson, A Ionescu, C Ivenso, C James, T James, M Jones, S Jones, L Lewis-Prosser, A Lucey, G Mallison, R Manikonda, G Marshall, J Northfield, L Parfitt, L Peter, C Price, M Pynn, D Ritchie, A Roynon-Reed, M Scott, C Sing, J Singh, M Singh, C Somashekar, J Summers, R Venkataramakrishnan, G Warwick, A Waters, G Williams, K Zalewska, the Blood Transfusion laboratory team.

**Dartford and Gravesham NHS Trust** B Khan (PI), D Ail, R Aldouri, G Awadzi, B Bassoy, R Bhalla, J Billings, S Bokhari, G Boniface, J Cernova, T Chen, P Chimbo, N Chitalia, S Danso-Bamfo, D Depala, A Dhanoa, T Edmunds, E Fernandez, T Ferrari-Gersogamo, B Fuller, Z Galani, A Gherman, P Grist, R Heire, L Ilves, L Lacey, E Lawrence, M Lewis, A Maric, W Martin, Z Min, C Newman, R Nicholas, O Olufuwa, N Pieniazek, T Qadeer, S Rathore, S Sathianandan, C Scott, A Shonubi (Associate PI), S Siddique, G Sisson, M Soan, D Streit, C Stuart, M Szekely, W Umeojiako, S Urruela, B Warner, M Waterstone, S White, K Yip, A-M Zafar, S Zaman, the Blood Transfusion laboratory team.

**Chesterfield Royal Hospital NHS Foundation Trust** N Spittle (PI), P Hickey (Co-PI), S Beavis, S Beghini, L Blundell, J Bradder, J Cresswell, A Crowder, K Dale, A Foo, J Gardner, R Gascoyne, M Kelly-Baxter, C Lattimore, L Lawson, R Lewis (Associate PI), E Mackay, E Moakes, K Moxham, A Padmakumar, J Pobjoy (Associate PI), K Pritchard, J Salmon, C Sampson, A Smith, J Smith, L Stevenson, S Vittoria, N Weatherly, A Whileman, E Wolodimeroff, S Wright, the transfusion lab team.

**Ashford and St Peter's Hospitals NHS Foundation Trust** C Russo (PI), M Aquino, M Croft, J Finden, V Frost, M Gavrila, K Gibson, A Glennon, C Gray, N Holland, N Johnson, J Law, R Pereira, P Reynolds, Z Takats, H Tarft, J Thomas, L Walding, A Williams, the Blood Transfusion Laboratory team.

**Frimley Health NHS Foundation Trust** M Meda (PI), J Democratis (Co-PI), N Barnes, N Brooks, L Chapman, C Cole, J da Rocha, R Dolman, K East, A Edwards, J Finden, T Foster, F Fowe, E Gaywood, S Gee, S Jaiswal, N Johnson, A Kandaswamy, K Kislingbury, M Molloholli, J Newanji, A Raguro, F Regan, L Rowe-Leete, C Smith, M van de Venne, T Weerasinghe, the Blood Transfusion laboratory teams.

**London North West University Healthcare NHS Trust** A Whittington (PI), J Barnacle, J Barrett, K Buckmire, A Capps-Jenner, F Cawa, S Chhabra, S Chita, M Colin, A Dagens, E Davenport, E Dhillon, S Filson, J Goodall, R Gravell, A Gupta-Wright, S Gurram, H Houston, G Hulston (Associate PI), S Isralls, R Kahari, C Kukadiya, J Low, K Man, M Nassari, B Nayar, O Ojo, P Papineni, V Parris, M Patel, R Patel, S Quaid, A Sturdy (Associate PI), B Tyagi, L Vaccari, N van der Stelt, G Wallis, E Watson, D Wiles, the Blood Transfusion laboratory team.

**Southern HSC Trust** R Convery (PI), J Acheson, J Brannigan, D Cosgrove, C McCullough, D McFarland, R McNulty, S Sands, G Scott, O Thompson, T Thompson, the Blood Transfusion laboratory team.

**Royal Devon and Exeter NHS Foundation Trust** M Masoli (PI), H Bakere, A Bowring, T Burden, P Czylok, J Davies, L Dobson, A Forrest, E Goodwin, H Gower, A Hall, L Knowles, H Mabb, A Mackey, V Mariano, E McEvoy, L Mckie, P Mitchelmore, L Morgan, R Oram, N Osborne, S Patten, J Piper, I Seaton, R Sheridan, D Sykes, J Tipper, S Whiteley, S Wilkins, N Withers, K Zaki, the Blood Transfusion laboratory team.

**Hull University Teaching Hospitals NHS Trust** N Easom (PI), K Adams, G Barlow, R Barton, H Bridge, M Bryan, A Burns, D Clark, L Cullen, R Elshaw, P Gunasekera, A Harvey, M Hayes,

M Ivan, S Khan, M Kolodziej, P Lillie, V Lowthorpe, V Mathew, S Mongolu, I Muazzam, D Norton, K Pearson, T Perinpanathan, C Philbey, B Pickwell-Smith, C Poole, L Rollins, A Samson, T Sathyapalan, L Sherris, K Sivakumar, K Smith, K Smith, C Steel, P Swift, T Taynton (Associate PI), G Walker, A Wolstencroft, H Yates, the Blood Transfusion laboratory team.

**Bedford Hospital NHS Trust** E Thomas (PI), D Bagmane (Co-PI), B Jallow (Co-PI), I Nadeem (Co-PI), M Negmeldin (Co-PI), A Vaidya (Co-PI), J Addai, A Amjad, N Amjad, A Anthony-Pillai, I Armata, R Arora, A Ashour, V Bastion, R Bhanot, D Callum, K Chan, P Chrysostomou, C Currie, N Daoud, B Djeugam, F De Santana Miranda, C Donaldson, M Ellison, P Eskander, M Farinto, S Farnworth, N Fatimah, C Fernando, R Field, L Fitzgerald, K Gelly, L Grosu, A Haddad, M Hannun, T Hashimoto, K Hawes, E Hawley-Jones , M Hayathu, M Hikmat, S Hussain, B Jallow, C Jenkins, U Keke, U Khatana, I Koopmans, T Large, P Lemessy, H Lindsay- Clarke, E Lister, T Lodge, R Lorusso, G Lubimbi, F Miranda, J Morris, N Nasheed, N Nathaniel, K Ng, C Ong, K Pandya, M Penacerrada, J Prieto, Q Quratulain, S Rabbini, S Rahama, A Rasarathnam, M Revels, L Salih, J Sarella, Z Shaida, V Simbi, N Simon, B Small, M Smith, K Solanki, W Tan, S Trussell, U Vaghela, A Vaidya, J Valentine, I Walker, O Walker, F Wang, R Wulandari, L Ylquimiche, the Blood Transfusion laboratory team.

**Royal Berkshire NHS Foundation Trust** M Frise (PI), R Arimoto, A Aslam, A Barrett, A Burman, C Camm, H Coles, J D'Costa, A , E Duffield, F Emond, S Everden, E Gabbitas, E Garden, A Hayat, S Haysom, T Hawkins, N Jacques, L Keating, K Kemsley, C Knowles, K Lennon, B Mitchell, T Okeke, L Sathyanarayanan, F Selby, M Thakker, A Walden, the Blood Transfusion laboratory team.

**Sheffield Teaching Hospitals NHS Foundation Trust** P Collini (PI), S Ali, A Ang, J Belcher, L Chapman, K Chin, D Cohen, J Cole, H Colton, R Condliffe, M Cribb, S Curran, T Darton, D de Fonseka, T de Silva, A Dunn, E Ferriman, R Foster, J Greig, J Hall, M Ul Haq, S Hardman, E Headon, C Holden, L Hunt, E Hurditch, F Kibutu, T Kitching, L Lewis, T Locke, H Luke, L Mair, P May, J Meiring, J Middle, J Middleton, P Morris, T Newman, L Passby, R Payne, G Powell, G Rana, S Renshaw, A Rothman, D Sammut, S Sherwin, P Simpson, M Sterrenburg, B Stone, M Surtees, B Taylor, A Telfer, R Thompson, Z Johnson, N Vethanayagam, R West, H Wilkinson, T Williams, G Margabanthu, the Blood Transfusion laboratory teams.

**Western Sussex Hospitals NHS Foundation Trust** L Hodgson (PI), M Margarson (Co-PI), L Albon, K Amin, M Bailey, Y Baird, A Butler, V Cannons, C Chandler, A Colino-Acevedo, N Falcone, S Floyd, L Folkes, H Fox, S Funnell, J Gilbert, R Gomez-Marcos, C Gonzalez, D Green, K Hedges, G Hobden, A Hunter, M Iftikhar, P Jane, R Khan, S Kimber, K King, M Linney, L Lipskis, J Margalef, T Martindale, A Mathew, P McGlone, E Meadows, S Moore, S Murphy, L Norman, R O'Donnell, J Park, D Raynard, D Reynish, C Ridley, Z Sammut, T Shafi, T Shaw, J Smith, S Sweetman, Y Thirlwall, S Troedson, T Tsawayo, E Vamvakiti, R Venn, N White, J Wileman, E Yates, the Blood Transfusion laboratory team.

**Swansea Bay University Local Health Board** B Healy (PI), S Bareford, I Blyth, A Bone, E Brinkworth, R Chudleigh, P Diamond, Y Ellis, D Evans, S Georges, S Green, R Harford, A Holborow, S John, C Johnston, P Jones, M Krishnan, N Leopold, F Morris, A Mughal, C Murphy, L O'Connell, L Park, D Payne, K Phillips, T Rees, S Richards, M Ryan, G Saleeb, C Thomas, P Vallabhaneni, J Watts, M Williams, the Blood Transfusion laboratory team.

**Norfolk and Norwich University Hospitals NHS Foundation Trust** E Mishra (PI), C Atkins (Co-PI), J Nortje (Co-PI), D Archer, D Asher, S Bailey, M Cambell-Kelly, E Chiang, P Clarke, L Coke, B Collinson, J Cook, A Cooper, M Cornwell, A Dann, S Fletcher, K Ford, C Fraser, M Del Forno, S Rehman, S Gajebasia, H Gorick, N Gray, A Haestier, S Hand, M Harmer, L Harris, L Hudig, M Ilsley, K Jethwa, L Jones, A Kamath, D Kelly (Associate PI), J Kennedy, J Keshet-Price, J Knights, E Kolokouri, V Licence, E Lowe, E Malone, M Marks, G Maryan, K Metcalfe, A Miller-Fik, P Moondi, M Morris, K Myint, K Nagumantry, W Ng, J Nortje, M Oliver,

M Patel, T Potter, J Pring, R Rallan, J Roberts, A Rudd, A Sanda-Gomez, A Sanz-Cepero, J Staines, K Stammers, E Tropman, M Ur Rasool, S Walton, N Ward, D Watts, C Websdale, R Wiseman, C Wright, the Blood Transfusion laboratory team.

**Sherwood Forest Hospitals NHS Foundation Trust** M Roberts (PI), K Amsha(Co-I), G Cox (Co-I), N Downer (Co-I), D Hodgson (Co-I), L Hostler, J Hutchinson (Co-I), S Kalsoom (Co-I), A Molyneux (Co-I), Z Noor (Co-I), L Allsop, T Brear, P Buckley, L Dunn, M Gill, C Goodwin, C Heeley, M Holmes, R Holmes, E Langthorne, C Moulds, D Nash, J Rajeswary, S Shelton, K Slack, S Smith, B Valeria, J Walden, J Wren, I Wynter, M Yanney, I Ynter, Senior BMS, the Blood Transfusion laboratory team.

**Isle Of Wight NHS Trust** A Naqvi (PI), M Pugh (PI), A Abbas, A Attia, A Azkoul, A Bester, A Brown, G Debreceni, R Fasina, A Gardener (Associate PI), P Gartan, S Grevatt, S Hinch, O Hudson, E Jenkins, N Kadam, A Khan, S Knight, X Liu, C Magier, J Mattappillil, N Mburu, B Mohamud, S Mukhtar, R Naik, P Nathwani, E Nicol, E O'Bryan, B Osman, N Otey, B Rizvi, P Rogers, J Selley, I Smith, M Stevens, L Sylvester, O Tarin, C Taylor, E White, J Wilkins, R Young, the Blood Transfusion laboratory team.

**Worcestershire Acute Hospitals NHS Trust** C Hooper (PI), K Austin, T Dawson, A Durie, G Godding, C Hillman-Cooper, C Khan, M Ling, E Loxley, E Murphy, S Stringer, H Tranter, J Tyler, P Watson, H Wood, the Blood Transfusion laboratory team.

**Blackpool Teaching Hospitals NHS Foundation Trust** J Cupitt (PI), N Ahmed, O Assaf, A Barnett, L Benham, P Bradley, Z Bradshaw, M Brunton, T Capstick, M Caswell, V Cunliffe, R Downes, N Fuller, P Hudson, N John, N Latt, R McDonald, A Mulla, E Mutema, J Navin, C O'Reilly, A Parker, S Preston, N Slawson, H Stannard, E Stoddard, S Traynor, V Vasudevan, E Ward, S Warden, J Wilson, A Zmierczak, the Blood Transfusion laboratory team.

**Northumbria Healthcare NHS Foundation Trust** B Yates (PI), C Ashbrook-Raby, A Aujayeb, L Barton, C Bomken, S Bourke, H Campbell, D Charlton, D Cooper, K Connelly, S Dodds, S Fearby, V Ferguson, N Green, H Grover, E Hall, I Hamoodi, S Haney, P Heslop, J Luke, L Mackay, C McBrearty, I McEleavy, N McLarty, U McNelis, A Melville, J Miller, A Morgan, S Parker, L Patterson, S Pick, D Ripley, S Robinson, J Rushmer, D Snell, J Steer, E Sykes, A Syndercombe, C Tanney, L Taylor, K Ward, K Wardle, M Weatherhead, R Whittle, L Winder, the Blood Transfusion laboratory team.

**United Lincolnshire Hospitals NHS Trust** M Chablani (PI), R Barber (Co-PI), S Archer, S Beck, S Butler, A Chingale, C Flood, O Francis, C Hewitt, A Hilldrith, B Holmes, A Jackson, A Kirkby, S MacDonald, R Mishra, K Netherton, M Okubanjo, L Osborne, A Reddy, C Richardson, A Sloan, the Blood Transfusion laboratory team.

**St George's University Hospitals NHS Foundation Trust** T Bicanic (PI), T Harrison (Co-PI), Y Aceampong, A Adebiyi, M Ali, D Baramova, M Caudwell, J De Sousa, S Drysdale, K Feane, J Hayat, A Janmohamed, A Khalil, A Lisboa, M Mencias, V Michael, C Orchard, K Patel, A Rana, N Said, T Samakomva, A Seward, O Skelton, K Spears, A Sturdy, V Tavoukjian, J Texeira, S Tinashe, J Uprichard, the transfusion laboratory team.

**Bolton NHS Foundation Trust** M Balasubramaniam (PI), C Subudhi (Co-PI), C Acton, S Altaf, S Ahmad, R Ahmed, A Ajmi, A Al-Asadi, A Amin, A Bajandouh, R Carey, Z Carrington, J Chadwick, S Cocks, C Dawe, K Farrar, S Farzana, O Froud, A Gibson, P Hill, A Hindle, R Holmes, G Hughes, R Hull, M Ijaz, J Jeewa, R Kalayi, M Khan, A Koirata, S Latham, G Lipscomb, K Lipscomb, J McCullagh, M McNulty, O Meakin, N Meghani, N Natarajan, D Nethercott, P Nicholas, A Parkinson, V Priyash, T Pandya, L Pugh, J Rafique, J Robertson, M Saleh, D Shaw, W Schneblen, B Sharma, O Sharma, Z Shehata, J Shurmer (Associate PI), R

Sime, S Singh, R Smith, R Tallent, E Tanton, D Tewkesbury, P Thet, S Thornton, J Timerick, C Underwood, N Wang, M Watts, I Webster, B Wilson, transfusion laboratory staff.

**Doncaster and Bassetlaw Teaching Hospitals NHS Foundation Trust** C Wong (PI), S Allen, Y Aung, G Bell, R Chadwick, R Codling, M Cox (Associate PI), R Crawforth, F Dunning, M Fairweather, N Hennessy, G Herdman, M Highcock, G Kirkman, N Khoja, C Knapp, M Kyi, V Maxwell, E Mustafa, A Nasimudeen, A Natarajan, P Prentice, D Pryor, E Richards, L Thompson (Associate PI), D Walstow, L Warren, N Wilkinson, the Blood Transfusion laboratory team.

**Kingston Hospital NHS Foundation Trust** S Mahendran (PI), A Joseph (Co-PI), M Agarwal, S Bacciarelli, A Baggaley, G Bambridge, F Bazari, P Beak, S Bidgood, T Binas, L Boustred, D Caldow, T Carlin, S Cavinato, B Church, A Conroy, J Crooks, E Donnelly, A Edwards, L Elliot, S El-Sayeh, A Feben, J Fox, R Gisby, M Grout, S Hassasing, R Heath, R Herdman-Grant, E Jackson, D Jajbhay, O James, S Jones, J Khera, K Muralidhara, T Leahy, S Luck, R Maamari, M Madden, H Matthews, J May, M McCullagh, G McKnight, L Mumelj, G Natarajan, D Newman, A Nicholson, T O'Brien, J Odone, R Patterson, R Patterson, A Pavely, J Poxon, C Quamina, A Ratnakumar, R Rodriguez-Belmonte, T Sanderson, S Sharma, R Simms, D Sivakumran, A Sothinathan, A Swain, S Swinhoe, M Taylor, E Thuesday, M Trowsdale Stannard, B Ummat, J Vance-Daniel, H Warren-Miell, R Williams, T Woodhead, D Zheng, the Blood Transfusion laboratory team

**Mid Yorkshire Hospitals NHS Trust** A Rose (PI), J Ashcroft (Co-I), P Blaxill (Co-I), S Bond (Co-I), A Dwarakanath (Co-I), C Hettiarachchi (Co-I), B Sloan (Co-I), S Taylor (Co-I), M Thirumaran (Co-I), R Allan, R Beckitt, I Bennett, S Bhaskara, C Blears, S Buckley, G Castle, E Clayton, N de Vere, J Ellam, D Gomersall, S Gordon, J Greaves, A Harrison, J Hewitson, W Hussain, C Hutsby, R Kousar´┐¢, K Lindley, C McBride, S Oddy, M O'Shea, A Poole (Associate PI), L Slater, B Sloan, B Taylor, S Taylor, M Thornton, M Wilkinson, the Blood Transfusion laboratory team.

**York Teaching Hospital NHS Foundation Trust** J Azam (PI), K Chandler (PI), M Abdelfattah, A Abung, J Anderson, P Antill, S Appleby, P Armtrong, T Berriman, D Bull, O Clayton, A Corlett, D Crocombe, S Davies, K Elliott, L Fahel, J Fullthorpe, J Ghosh, N Gott, D Greenwood, T Holder, P Inns, C Ivel, W Lea, E Lindsay, K Mack, N Marshall, S McMeekin, T Momoniat, P Nikolaos, M O'Kane, G Patrick, H Pearson, P Ponnusamy, A Poole, N Price, R Proudfoot, C Purcell, G Purssord, S Roche, C Sefton, S Shahi, D Smith, B Sohail, R Thomas, A Thorpe, A Turnbull, L Turner, H Watchorn, E Wiafe, J Wilson, D Yates, the Blood Transfusion laboratory team.

**North Middlesex University Hospital NHS Trust** J Moreno-Cuesta (PI), S Rokadiya (Co-PI), T Amin, P Brooks, A Govind, A Haldeos, K Leigh-Ellis, K Madgwick, S Palihavadana, P Sandajam, C Vansomeren, R Vincent, L Walker, the Blood Transfusion laboratory team.

**Great Western Hospitals NHS Foundation Trust** A Kerry (PI), V Barlow, A Beale, S Bhandari, A Brooks, C Browne, J Butler (Associate PI), J Callaghan, B Chandrasekaran, S Charlton R Davies, L Davies, T Elias, D Finch, P Foley, E Fowler, G Gowda, J Gregory, A Ipe, A Jaffery, M Juniper, S Khan, I Laing-Faiers, C Lewis-Clarke, J Lodge, C Mackinlay, P Mappa, A Maxwell, W Mears, E Mousley, T Novak, L Pannell, S Peglar, A Pereira, I Ponte Bettencourt dos Reis, E Price, A Quayle, Q Qurratulain, S Small, H Smith, C Strait, E Stratton, M Tinkler, J Uppal, A van der Meer, A Waldron, R Waller, M Watters, S White, L Whittam, T Wiliams, Z Xia, K Yein, V Zinyemba, the Blood Transfusion team.

**Imperial College Healthcare NHS Trust** G Cooke (PI), E Adewunmi, Z Al-Saadi, R Ashworth, J Barnacle, L Chapple, V Chiroma, F Chowdhury, A Daunt (Associate PI), H Dawson, L Evison, S Fernandez Lopez, C Gale, M Gibani, A Jimenez Gil, D Johnson, B Jones, J Labao,

S Kassa, N Madeja, S Mashate, H McLachlan, U Munu, E Nweje, A Perry, A Rahman, F Regan, K Sabljak, R Thomas, E Whittaker, C Wignall, P Wilding, L Young, C Yu, the Blood Transfusion laboratory teams.

**James Paget University Hospitals NHS Foundation Trust** J Patrick (PI), B Burton (PI), A Ayers, R Brooks, J Chapman, V Choudhary, S Cotgrove, G Darylile, L Felton, D Griffiths, C Hacon, H Hall, W Harrison, P Hassell, A Hearn, F Iqbal Sait, J Jackson, K Mackintosh, J North, S Parslow-Williams, S Parsons, H Sutherland, M Whelband, C Whitehouse, E Wilhelmsen, J Wong, J Woods, the Blood Transfusion laboratory team.

**Sandwell and West Birmingham Hospitals NHS Trust** S Clare (PI), M Ahmed, Y Beuvink, K Blachford, L Blackwood, V Chauhan, S Clamp, J Colley, P De, M Fenton, B Gammon, A Hayes, L Henry, S Hussain, S Joseph, F Kinney, T Knight, R Kumar, W Leong, T Lim, B Mahay, Y Nupa, A Orme, W Osborne, Z Pilsworth, S Potter, S Prew, N Rajaiah, A Rajasekaran, H Senya, N Shah, N Shamim, S Sivakumar, L Smith, P Thozthumparambil, N Trudgill, A Turner, L Wagstaff, S Willetts, H Willis, M Yan, the Blood Transfusion laboratory teams.

**University Hospitals Plymouth NHS Trust** D Lewis (PI), D Affleck, O Anichtchik, K Bennett, M Binney, J Corcoran, M Cramp, H Davies, J Day, M Dobranszky Oroian, E Freeman, C Lowe, L Madziva, P Moodley (Associate PI), C Morton, M Mwadeyi, H Notman, C Orr, A Patrick, L Pritchard, G Selby, J Shawe, H Tan, N Thiri Phoo (Associate PI), the Blood Transfusion laboratory team.

**University College London Hospitals NHS Foundation Trust** H Esmail (PI), R Heyderman (Co-PI), V Johnston (Co-PI), D Moore (Co-PI), F Beynon, R Atugonza, P Bodalia, X Chan, Z Chaudhry, CY Chung, J Cohen, D Crilly, S Eisen, N Fard, J Gahir, L Germain, J Glanville, Z Jeewa, V Johnston, E Kilich, N Lack, M Merida Morillas, J Millard, N Platt, S Roy, I Skorupinska, M Skorupinska, J Spillane, the Blood Transfusion laboratory team.

**The Newcastle Upon Tyne Hospitals NHS Foundation Trust** A De Soyza (PI), R Agbeko, A Baird, K Baker, A Barr, E Cameron, Q Campbell Hewson, C Duncan, M Emonts, M Evans, A Fenn, S Francis, J Glover Bengtsson, A Greenhalgh, A Hanrath, K Houghton, D Jerry, G Jones, S Kelly, C Kennedy, N Lane, J Macfarlane, P McAlinden, I McCullagh, S McDonald, O Mohammed, A Muir, R Obukofe, J Parker, A Patience, B Payne, D Price, H Ranton, Z Razvi, S Robson, S Rowan-Ferry, A Sanchez Gonzalez, G Smithson, E Stephenson, R Welton, S West, M Wilson, E Wong, F Yelnoorkar, the Blood Transfusion laboratory team.

**The Royal Bournemouth and Christchurch Hospitals NHS Foundation Trust** M Schuster Bruce (PI), D Baldwin, C Baylem, V Chandler-Vizard, Z Clark, M Dale, B Grice, D Griffiths, E Gunter, T Haydon, S Horler, A Jose, T Joyce, M Keltos, S Kennard, N Lakeman, L Mallon, R Miln, L Mounsey, S Nix, S Orr, S Pitts, L Purandare, L Rogers, J Samways, E Stride, L Vamplew, M Trevett, L Wallis, the Blood Transfusion laboratory team.

**University Hospitals Of North Midlands NHS Trust** T Kemp (PI), J Alexander, W Al-Shamkhani, C Baker, N Bandla, D Bentley, N Bodasing, C Brackstone, E Brown, A Cadwgan, M Chaudhary, L Cheng, S Church, F Clark, M Davies, L Diwakar, A Farmer, F Farook, M Gellamucho, K Glover, J Graham, M Haris, J Humphries, I Hussain, P Irving, J Jeffrey, S Khan, L Korcierz, J Lee, J Marshall, H McCreedy, S Mitchell, G Muddegowda, D Murdoch, I Mustapha, K Nettleton, Z Noori, S O'Brien, W Osman, N Patel, F Perkins, M Ram, A Remegoso, L Rogers, R Rushworth, A Quinn, E Sadler, A Salmon, L Scott, T Scott, N Sheikh, R Sivers, R Swift, C Thompson, J Tomlinson, L Walker, J Weeratunga, J White, C White, P Wu, the Blood Transfusion laboratory teams.

**Warrington and Halton Teaching Hospitals NHS Foundation Trust** M Moonan (PI), M Murthy (PI), R Arya, R Chan, A Baluwala, B Chesterson, L Connell, M Davey, L Ditchfield, G Drummond, K Galilee, A Ibrahim, J Little, N Marriott, T Nagarajan, S Patel, H Prady, L Roughley, S Sharma, H Whittle, J Yates, the Blood Transfusion laboratory team.

**Northern Lincolnshire and Goole NHS Foundation Trust** A Mitra (PI), M N Akhtar, H Al-Moasseb, S Amamou, E Baker, T Behan, S Biuk, S Bollington, J Braithwaite, M Brazil, H Briggs, M Brocken, C Burnett, C Chatha, M Cheeseman, L-J Cottam, T Cruz Cervera, O Davies, K Dent, C Dyball, K Edwards, R Elmahdi, Q Farah, S Farooq, K Garnett, S Gooseman, J Hargreaves, MA Haroon, K Garnett, J Hatton, E Heeney, J Hill, E Horsley, R Hossain, D Hutchinson, J Hyde-Wyatt, M Iqbal, N James, S Khalil, H Jackson, M Madhusudhana, A Marriott, M T Masood, G McTaggart, K Mellows, R Miller, U Nasir, M Newton, GCE Ngui, S Pearson, C Pendlebury, R Pollard, N Pothina, D Potoczna, SD Raha, A Rehan, SAS Rizvi, A Saffy, K Shams, C Shaw, A Shirgaonkar, S Spencer, R Stead, R Sundhar, D Taylor, E Thein, L Warnock, A White, KY Wong, the Blood Transfusion laboratory teams.

**Whittington Health NHS Trust** C Parmar (PI), S Ahmed , W Ang, S Ashcroft , H Bateman, D Burrage, M Christy, P Dlouhy, R Fromson, G Fung , K Gilbert, F Green, M Horsford , L Howard, I Jenkin, C Jetha , M Kousteni, N Kulkarni, A Kyei-Mensah, I Lim, S Lock, C Macfadyen, S Marston, C Mason , P McCormack , J Merritt, S Mindel, H Montgomery , S Myers, L Parker, B Pattenden, A Pender, A Pratley, N Prevatt, A Reid, S Rudrakumar, J Sabale, S Sharma, P Sharratt, K Simpson , J Southwell , S Taylor, L Veys, N Wolff, A Zuriaga-Alvaro, D Worley, the Blood Transfusion laboratory team.

**Cwm Taf Morgannwg University LHB** C Lynch (PI), C Brookes, B Deacon, S Eccles, T Edwards, A Gaurav, B Gibson, K Hennessy, M Hibbert, T Home, K Jones, C Lai, R Lawton, L Margarit, D Nair, S Owen, T Payne, R Pritchard, J Pugh, L Roche, S Sathe, J Singh, D Tetla, R Ward, C Woodford, the Blood Transfusion laboratory team.

**Southend University Hospital NHS Foundation Trust** G Koduri (PI), S Gokaraju (Co-PI), F Hayes (Co-PI), V Vijayaraghavan Nalini (Co-PI), S Badhrinarayanan, N Chandran, J Galliford, T Green, V Gupta, P Harman, M Mercioniu, N Outten, D Qureshi, M Rabbani, the Blood Transfusion laboratory team.

**St Helens and Knowsley Teaching Hospitals NHS Trust** G Barton (PI), P Stockton (Co-PI), A Tridente (Co-PI), S Adair, N Collins, S Dealing, R Garr, S Greer, J Harrison, V Hayes, N Hornby, J Keating, T Kelly, S Mayor, A McCairn, S Rao, K Shuker, J Kirk, T Thornton, M Wood, the Blood Transfusion laboratory team.

**University Hospitals Coventry and Warwickshire NHS Trust** K Patel (PI), C Imray (Co-PI), N Aldridge, A Ali, C Bassford, A Busby, A Campbell, G Evans, E French, R Grenfell, S Hewins, D Hewitt, J Jones, R Kumar, E Mshengu, J Northcote, S Quenby, H Randheva, K Read, P Satodia, E Sear, T Taylor, M Truslove, the Blood Transfusion laboratory team.

**Southport and Ormskirk Hospital NHS Trust** S Pintus (PI), H Ahmed, L Bishop, S Burns, D Dickerson, P Gordon, J Harrison, Z Haslam, E Isherwood, M Jackson, A McMahon, A Morris, M Morrison, R Seabury, A Tageldin (Associate PI), M Wood, the Blood Transfusion laboratory team.

**Cardiff & Vale University LHB** C Fegan (PI), G Andrikopoulos, A Balan, B Basker, S Bird, R Borrell, Z Boult, V Britten, L Broad, S Burns, R Carnegie, H Cendl, M Chakraborty, J Cole, M Edgar, T Evans, J Evans, M Evans, J Forton, S Frayling, F Greaves, S Harrhy, M Haynes, H Hill, Z Hilton, L Jones, A Kelly, L Knibbs, D Lau, J May, E McGough, A McQueen, S McWilliam, J Milner, M Morgan, C M Neville, R Norman, K Nyland, C Oliver, K Paradowski, L Parkinson, M Patal, A Patterson, K Rahilly, C Robinson, S Scourfield, M Starr, S Struik, E Thomas, R

Thomas-Turner, J Underwood, G Williams, J Williams, M Williams, M Wise, S Zaher, the Blood Transfusion laboratory team.

**King's College Hospital NHS Foundation Trust** M McPhail (PI), J Adeyemi, J Aeron-Thomas, M Aissa, K Amenyah, A Buazon , T Buttle, S Candido, A Cavazza, M Cesay, K Clark, M Cockerell, S Cole, A Comerford, J Davies, M Depante, C Donaldson, K El-Bouzidi, C Finney , J Galloway, S Gogoi, P Gold, N Griffiths, A Gupta, J Hannah, C Hawkins, Y Hu, E Jerome (Associate PI), N Kametas, M Kumar, L Linkson, N Long, H Martin, D Nagra, H Noble, K Nwankiti, K O'Reilly, V Patel, T Pirani, D Rao, S Ratcliff, S Saha , B Sari, N Sikondari, J Smith, A Te, C Tey, R Uddin, D Veniard, J Vidler (Associate PI), J Ward, M West, C Williamson, M Yates, A Zamalloa, the Blood Transfusion laboratory team.

**Shrewsbury and Telford Hospital NHS Trust** J Moon (PI), R Baldwin-Jones, N Biswas, A Bowes, H Button, M Carnahan, E Crawford, S Crawford, S Deshpande, D Donaldson, C Fenton, S Hester, Y Hussain, M Ibrahim, J Jones, S Jose, H Millward, N Motherwell, M Rees, N Schunke, A Stephens, J Stickley, M Tadros, H Tivenan, E Wilson, the Blood Transfusion laboratory team.

**Northern HSC Trust** P Minnis (PI), W Anderson, S Baird, M BinRofaie, J Burns, C Cruickshank, L Davidson (Associate PI), L Denley, R Donnelly, G Doran, M Drain, A Fryatt, J Gallagher, B Graham, C Henry, M Hollyer, M Kawser, L Kingsmore, O Lepiarczyk, I Masih, S McKenna, C McGoldrick, M McMaster, S McNeill, E Murtagh, M Nugdallah, K Potter, T Scullion, M Wray, the Blood Transfusion laboratory team.

**North Bristol NHS Trust** N Maskell (PI), S Barratt (Sub-PI), J Dodd (Sub-PI), R Adhikary (Associate PI), H Adamali, M Alvarez, D Arnold, P Bailey, G Bardsley, S Bevins, R Bhatnagar, A Bibby, L Bradshaw, C Burden, H Cheshire, S Clarke, A Clive, R Cousins, S Dawson, A Dipper, F Easton, K Farmer, L Gethin, P Halford, F Hamilton, J Hardy, D Higbee, S Holloway, A Jayebalan, A Jenkins, L Jennings, S Jones, C Kilby, H Lee, H McDill, H McNally, K Mead, A Milne, E Morab, A Morley, G Nava, A Pereira, E Perry, N Rippon, V Sandrey, C Sellar, K Smith, L Solomon, L Stadon, J Townley, R Wach, D Warbrick, C Watkins, H Welch, T Wreford-Bush, the Blood Transfusion laboratory team.

**The Royal Wolverhampton NHS Trust** S Gopal (PI), R Barlow, M Blanton, M Boyd, C Cheong, D Churchill, K Davies, M Green, N Harris, M Herbert, A Kumar, S Metherell, S Milgate, L Radford, J Rogers, A Smallwood, L Wild, the Blood Transfusion Laboratory team.

**Tameside and Glossop Integrated Care NHS Foundation Trust** B Ryan (PI), A Abraheem, C Afnan, B Ahmed, O Ahmed, N Amar, M Anim-Somuah, A Armitage, P Arora, N Aziz, M Beecroft, T Bull, A Butt, S Chandra, J Fallon, A Fazal, J Foster, I Foulds, N Garlick, H Ghanayem, S Gulati, R Hafiz-Ur-Rehman, M Hamie, A Hewetson, B Ho, C Holt, V Horsham, W Hughes, W Hulse, A Humphries, M Hussain, N Johal, E Jude, A Kendall-Smith, M Kelly, M Khan, S Kilroy, R Law, G Lewis, J Majumdar, J McCormick, V Melnic, O Mercer, T Mirza, A Masud, B Obale, P Potla, S Pudi, K Qureshi, M Rafique, R Rana, R Roberts, J Roddy, C Rolls, M Sammut, H Savill, M Saxton, A Shanahan, M Summerton, V Turner, A Tyzack, J Vere, the Blood Transfusion laboratory team.

**South Eastern HSC Trust** D Alderdice (PI), J Courtney (Co-I), J Elder (Co-I), D Hart (Co-I), K Henry (Co-I), R Hewitt (Co-I), A Kerr (Co-I), J McKeever (Co-I), C O'Gorman (Co-I), S Rowan (Co-I), T Trinick (Co-I), B Valecka (Co-I), P Yew (Co-I), V Adell, J Baker, L Birch, A Campbell, S Carty, M El-Agnaf, J Foreman, P Gillen, Y Gail, S Graham, S Hagan, L Hammond, C Hewitt, J MacIntyre, C McKay, L Moore, L Palmer, S Regan, A Smith, G Young, the transfusion lab team.

**Harrogate and District NHS Foundation Trust** A Kant (PI), A Amin, C Bennett, O Cohen, S Foxton, R Gill, M Kuunal, E Lau, L Lowrey, C Morgan, N Singh, G Sugden, A Williamson, L Wills, the Blood Transfusion laboratory team.

**NHS Ayrshire and Arran: University Hospital Crosshouse** A Clark (PI), S Allen (Co-I), V Dey (Co-I), S Smith (Co-I), D Wilkin (Co-I), T Adams, C Burns, D Callaghan, N Connell, F Elliott, K Gibson, D Gilmour, M Henry, M McGovern, L McNeil, S Meehan, K Smith, C Turley, S Walton, M Wilson, S Wood, the Blood Transfusion laboratory team.

**Buckinghamshire Healthcare NHS Trust** R West (PI), J Abrams, B Alyas, A Baldwin, O Bannister, J Barker, H Beddall, H Blamey, O Bozdag, E Chan, J Chaplin, B Chisnall, C Cleaver, M Corredera, K Cotton, S Crotty, H Cui, L Day, P Dey, L Downs, B Hammans, M Kononen, S Kudsk-Iversen, A Kudzinskas, N Lambe, M Laurenson, R Mancinelli, J Mandeville, K Manso, S McLure, E Morgan-Smith, A Ngumo, H Noe (Associate PI), R Oxlade, A Parekh (Associate PI), V Pradhan, M Rahman, C Robertson, S Shah, K Shahzad, H Smith, J Tebbutt, N Vella, M Veres, A Watson, L Western, D Whitley, N Wong, M Zammit-Mangion, M Zia, the Blood Transfusion laboratory teams.

**The Hillingdon Hospitals NHS Foundation Trust** S Kon (PI), K Al-Asadi, T Bate, A Chan, W Chia, A Danga, O Dube, J Ganapathi, N Hadjisavvas , B Haselden, M Holden, T Hove, E Kam, J Korolewicz, M Kovac , A Lam, H Lamont, G Landers, P Law, N Mahabir, M Majumder, N Malhan, T Nishiyama, A Omojola, P Palanivelu, J Potter, D Quashie, S Ramraj, S Salawu, A Seckington, S Vandeyoon, W Varney, D Wahab, C Woollard, V Zindonda, the Blood Transfusion laboratory team.

**University Hospital Southampton NHS Foundation Trust** S Fletcher (PI), J Bigg, S Carrington, K Cathie, S Chabane, M Coleman, K Dowling, P Downey, S Faust, M Felongco, J Forbes, T Francis-Bacon, E Holliday, M Johnson, T Jones, C Jones, T Lofting, E Marouzet, S Michael, S Mumford, M Nelson, M Petrova, L Presland, A Procter, N Rayner, J Ricks, R Samuel, T Sass, M Shaji, C Silva Moniz, T Thomas, S Triggs, C Watkins, S Wellstead, H Wheeler, the Blood Transfusion laboratory team.

**Gloucestershire Hospitals NHS Foundation Trust** C Sharp (PI), F Ahmed, O Barker, O Bintcliffe, P Brown, R Bulbulia, T Clarke, J Collinson, T Cope, A Creamer, C Davies, W Doherty, M Fredlund, J Glass, S Harrington, A Hill, H Iftikhar (Associate PI), M James, S Jones, C Lim, S Message, H Munby, J Ord, J Patel, R Peek, T Pickett, A Simpson, M Slade, C Thompson, H Uru, D Ward, R Woolf, the Blood Transfusion laboratory team.

**Oxford University Hospitals NHS Foundation Trust** K Jeffery (PI), M Ainsworth, C Arnison-Newgass, A Bashyal, S Beer, A Bloss, D Buttress, W Byrne, A Capp, P Carter, P Cicconi, R Corrigan, C Coston, L Cowen, N Davidson, L Downs, J Edwards, R Evans, S Gardiner, D Georgiou, A Gillesen, A Harin, M Havinden-Williams, R Haynes, C Hird, A Hudak, P Hutton, R Irons, P Jastrzebska, S Johnston, M Kamfose, K Lewis, T Lockett, F Mendoza, F Maria del Rocio, J Martinez Garrido, S Masih, A Mentzer, S Morris, C O'Callaghan, Z Oliver, S Paulus, E Perez, L Periyasamy, L Peto, D Porter, S Prasath, C Purdue, M Ramasamy, C Roehr, A Rudenko, V Sanchez, A Sarfatti, M Segovia, T Sewdin, J Seymour, V Skinner, L Smith, A Sobrino Diaz, M Taylor-Siddons, J Staves, H Thraves, C Tsang, C Vander Riet, M Vatish, Y Warren, E Wilcock, the Blood Transfusion laboratory team.

**Guy's and St Thomas' NHS Foundation Trust** H Winslow (PI), P Dargan (Co-I), M Ostermann (Co-I), M Ahmad, K Bisnauthsing, L Brace, L Bremner, K Brooks, B Browne, J Butler, M Carter, B Castles , L Chappell, P Cinardo, D Cooke, J Cordle, B Crane, T Crowley, M Flanagan, O Fox, C Furtado, B Hamilton, D Hydes, B Jackson, T Jayatilleke, D Jeyapalan, L Jimenez, J Jones, J Kenny, H Kerslake, E Lee, A Lewin, C Lobato, T Maggs, N Martin, L Martinez, M Mathew, A Mazzella, L McCabe, B Merrick, S Mieres, F Morselli, L Nel, M Ng, R

O'Dea, M Opena, T Rajeswaran, A Raynsford, J Ross, C Singh, L Snell, H Tarft, J Tidman, S Turawa, E Wayman, C Williamson, K Wilson, J Wood, U Wood, L Woodford, A Xhikola, C Yearwood Martin, the Blood Transfusion laboratory teams.

**NHS Greater Glasgow and Clyde: Royal Alexandra Hospital** A Corfield (PI), S Brattan, A Brunton, C Clark, P Clark, D Grieve, M Hair, M Heydtmann, E Hughes, L Imam-Gutierrez, D McGlynn, G Ray, J Robertson, N Rodden, K Rooney, N Thomson, the Hospital Blood Transfusion Team.

**Lancashire Teaching Hospitals NHS Foundation Trust** S Laha (PI), A Ashfaq, S Baines, J Beishon (Associate PI), A Bellis, D Cameron, M Chiu, W Choon Kon Yune, M Deeley, E Fisher, W Flesher, K Gandhi, S Gudur, R Gupta, B Houghton, A Huckle, G Long, E Ly, A McCarrick, J Mills, P Mulgrew, S Nawaz, J Nixon, A Noyon, O Parikh, S Punnilath Abdulsamad, D Rengan, E Seager, S Sherridan, B So, R Sonia, T Southworth, K Spinks, A Timoroksa, K Treuberg, B Vernon, D Wallbank, A Williams, K Williams, H Wu, the Blood Transfusion laboratory team.

**East Sussex Healthcare NHS Trust** A Marshall (PI), S Ahmed (Associate PI), S Bafekr Mishamandani, S Blankley, H Brooke-Ball, T Christopherson, M Clark, T de Freitas, E de Sausmarez, A Ekunola, D Hemsley, J Highgate, I Humphreys, O Kankam, A Lowe, S Merritt, Y N S Mohammed, T Morley, A Newby, S Panthakalam, S Qutab (Associate PI), R Reddy, N Roberts, M Simon, K Subba, S Tieger, A Trimmings, R Venn, F Willson, T Win, M Yakubi, A Zubir, the Blood Transfusion laboratory team.

**Poole Hospital NHS Foundation Trust** H Reschreiter (PI), S Bokhandi, J Camsooksai, V Chandler-Vizard, C Colvin, J Dube, B Grice, S Grigsby, C Humphrey, S Jenkins, S Patch, M Tighe, M Trevett, L Vinayakarao, B Wadams, M Woolcock, the Blood Transfusion laboratory team.

**NHS Lothian: Royal Infirmary of Edinburgh** A Gray, M Adam, R Al-Shahi Salman, A Anand, R Anderson, K Baillie, D Baird, T Balaskas, J Balfour, C Barclay, P Black, C Blackstock, S Brady, R Buchan, R Campbell, J Carter, P Chapman, M Cherrie, C Cheyne, C Chiswick, A Christides, D Christmas, A Clarke, M Coakley, A Coull, A Crawford, L Crisp, C Cruickshank, D Cryans, J Dear, K Dhaliwal, M Docherty, R Dodds, L Donald, S Dummer, M Eddleston, J Falconer, S Ferguson, N Fethers, E Foster, R Frake, N Freeman, B Gallagher, E Gaughan, D Gilliland, E Godden, E Godson, J Grahamslaw, A Grant, A Grant, N Grubb, S Hainey, Z Harding, M Harris, M Harvey, D Henshall, S Hobson, N Hunter, Y Jaly, J Jameson, D Japp, H Khin, L Kitto, S Krupej, C Langoya, R Lawrie, M Lindsay, A Lloyd, B Lyell, D Lynch, J Macfarlane, L MacInnes, I MacIntyre, A MacRaild, M Marecka, A Marshall, M Martin, E McBride, C McCann, F McCurrach, M McLeish, R Medine, H Milligan, E Moatt, W Morley, M Morrissey, K Murray, D Newby, K Nizam Ud Din, R O'Brien, M Odam, J Oldham, E O'Sullivan, R Penman, A Peterson, P Phelan, G Pickering, T Quinn, N Robertson, L Rooney, N Rowan, M Rowley, R Salman, C Scott, J Simpson, E Small, P Stefanowska, A Stevenson, S Stock, J Teasdale, I Walker, K Walker, A Williams, the Blood Transfusion laboratory team.

**NHS Greater Glasgow and Clyde: Glasgow Royal Infirmary** K Puxty (PI), J Alexander, L Bailey, H Bayes, A Begg, A Brown, Caldwell, S Carmichael, S Cathcart, F Christie (Associate PI), R Colbert, I Crawford, M A David, C Dunne, T Grandison, N Hickey, J Ireland, A Jamison, D Jenkins, J Johnstone, R Kearns, L Martin, M McIntyre, A Munro, S Nelson, C Ogilvie, H Peddie, G Piper, L Pollock, D Rimmer, K Scott (Associate PI), G Semple, S Thornton, the Transfusion Laboratory team.

**NHS Greater Glasgow and Clyde: Queen Elizabeth University Hospital** M Sim (PI), K Blyth (Co-PI), L Pollock (Co-PI), N Baxter, J Ferguson, K Ferguson, R Hague, A Hanlon, S Henderson, L Jawaheer, L Kelly, S Kennedy-Hay, A Kidd, M A Ledingham, F Lowe, M Lowe,

G McCreath, R McDougall, M McFadden, M McGarvey, M McGettrick, N McGlinchey, L McKay, C McKie, B McLaren, C McParland, J McTaggart, J Millar, A Molloy, A Phillips, J Rollo, H Stubbs, M Wilson, S Wishart, the Blood Transfusion laboratory team.

**Torbay and South Devon NHS Foundation Trust** T Clark (PI), I Akinpelu, S Atkins, J Blackler, J Clouston, G Curnow, M Fisher Crisp, A Foulds, J Graham, C Grondin, S Howlett, C Huggins, L Kyle (Associate PI), S Martin, P Mercer, S Mills, W O'Rourke, A Redome, A Penny, J Pinder, H Randall, J Redome, R Tozer, J Turvey, the Blood Transfusion laboratory team.

**Stockport NHS Foundation Trust** R Stanciu (PI), M Afridi, M Al Dakhola, S Bennett, L Brown, C Cooper, A Davison, B Devine, D Eleanor, J Farthing, A Ferrera, S Gandhi, L Gomez, P Haywood, C Heal, H Jackson, J Johnston, A Lloyd, S McCaughey, R Owen, A Pemberton, F Rahim, H Robinson, N Sadiq, R Samlal, V Subramanian, D Suresh, H Wieringa, I Wright, R Zaman, the Blood Transfusion laboratory team.

**NHS Lothian: St John's Hospital** S Lynch (PI), R Anderson, S Begg, J Bentley, S Brady, M Colmar, J Falconer, R Frake, A Gatenby, C Geddie, F Guarino, S Guettari, C Hurley, C Kuronen-Stewart, A MacRaild, M Mancuso-Marcello, M Marecka, G McAlpine, N McCullough, A Megan, T North, J Oldham, O Otite, L Primrose, L Rooney, A Saunders, A Saunderson, A Stevenson, S Stock, A Wakefield, E Walsh, J Wraight, T Wright, S Yusef, the Blood Transfusion laboratory team.

**NHS Lothian: Western General Hospital** O Koch (PI), A Abu-Arafeh, E Allen, L Bagshaw, C Balmforth, A Barnett-Vanes, R Baruah, S Begg, S Blackley, G Clark, S Clifford, D Dockrell, M Evans, J Falconer, V Fancois, C Ferguson, S Ferguson, N Freeman, E Gaughan, E Godden, S Hainey, R Harrison, B Hastings, S Htwe, A Ju Wen Kwek, O Lloyd, C Mackintosh, A MacRaild, W Mahmood, E Mahony, J McCrae, S Morris, C Mutch, K Nunn, J Oldham, D O Shea, I Page, M Perry, J Rhodes, N Rodgers, A Shepherd, G Soothill, S Stock, R Sutherland, A Tasiou, A Tufail, D Waters, R Weerakoon, T Wilkinson, R Woodfield (Associate PI), J Wubetu, the Blood Transfusion laboratory team.

**Kettering General Hospital NHS Foundation Trust** N Siddique (PI), M Abedalla, A Abeer Ul Amna, K Adcock, S Adenwalla, O Adesemoye, R Ahmed Ali, A Ali, S Aransiola, M Ashraf, M Ashraq, S Ashton, H Asogan, H Aung, A Aung, A Baral, F Bawani, S Beyatli, S Chowdhury, D Ramdin, J Dales, R Deylami, A Elliott, J Frankcam, S Gali, A Gkioni, L Hollos, A Houston, M Hussam El-Din, A Ibrahim, Y Jameel, G Keyte, J Khatri, S Kiran, R Kusangaya, C Little, S Little, E Lyka, A Madu, G Margabanthu, M Mustafa, H Naeem, D Ncomanzi, G Nikonovich, A Nisha James, Y Owoseni, N Pandian, D Patel, D Patel, I Petras, M Raceala, G Ramnarain, E Rich, S Saunders, A Shaikh, M Shakeel, M Silverstone, S Stapley, S Sudershan, P Swift, N Veli, A Virk, T Ward, A Wazir, S White, J Wood, N Zakir, the Blood Transfusion laboratory team.

**Hampshire Hospitals NHS Foundation Trust** R Partridge (PI), Y Abed El Khaleq, A Arias, E Bevan, J Conyngham, E Cox, M Cundall, E Defever, A Goldsmith, D Griffin, A Heath, L Holloway, B King, E Levell, X Liu, J Martin, O Martins, N Muchenje, M Mupudzi, A Nejad, S Senra, F Stourton, R Thomas, B White (Associate PI), G Whitlingum, C Wilson, L Winckworth, C Wrey Brown, S Zagalo, the Blood Transfusion laboratory team.

**NHS Dumfries and Galloway: Dumfries & Galloway Royal Infirmary** D Williams (PI), M McMahon (Co-PI), M McAdam, J Candlish, P Cannon (Associate PI), J Duignan, J Gardiner, C Jardine (Associate PI), S Jeffrey, A Mitra, P Neill (Associate PI), F Parker, S Svirpliene, S Wisdom, the Blood Transfusion laboratory team.

**Bradford Teaching Hospitals NHS Foundation Trust** D Saralaya (PI), N Akhtar, V Beckett, L Brear, V Drew, J Eedle, N Hawes, S Kmachia, N Mistry, S Moss, S Oddie, J Paget, J Parkinson, K Regan, D Ryan-Wakeling, A Shenoy, H Slater, K Storton, R Swingler, J Syson, J Todd, R Wane, A Whittingham-Hirst, A Wilson, the Blood Transfusion laboratory team.

**Cambridge University Hospitals NHS Foundation Trust** M Knolle (PI), E Gkrania-Klotsas (Co-PI), O Abani, R Bousfield (Associate PI), H Dakers-Black, T Dymond, T Foukanelli, K Gajewska-Knapik, J Galloway, S Grist, M Ibrahim, K Leonard, M Lewin, H Madiyiko, T Mikolasch (Associate PI), T Moore, C Newsam, N Pathan, K Philpott, V Rose, J Sahota, K Sharrocks (Associate PI), R Smith, the laboratory team.

**Royal United Hospitals Bath NHS Foundation Trust** J Suntharalingam (PI), H Ali, R Anstey, J Avis, C Broughton, A Buchanan, S Burnard, R Coles, T Cooke, C Demetriou, G Dixon (Associate PI), A Dornan, J Fiquet, J Ford, J Goodlife, O Griffiths, R Hamlin, T Hartley, S Jones, I Kerslake, D Loveys, J Macaro, R MacKenzie Ross, C Marchand, H Maria-Osborn, V Masani, R Mason, C McKerr, S Mitchard, J Nolan, A Palmer, K Pass, P Perera, L Ramos, M Rich, C Risbridger, J Rossdale (Associate PI), Z Sibanda, S Sturney, L Swan, G Towersey, K Turek, J Tyler, W Vietri, J Walters, A Wardle, K White, S Zulfikar, the laboratory team.

**Croydon Health Services NHS Trust** T Castiello (PI), J Adabie-Ankrah, G Adkins, B Ajay, A Ameen, S Ashok, B Cheung, S Conran, A Dean, S Dillane, F Fedel, V Florence, M Free, D Griffiths, I Griffiths, J Hajnik, J Hetherington, C Jones, A Latheef, S Lee, M Mbwembwe, J McCammon, S Patel, A Raghunathan, P Shah, J Talbot-Ponsonby, G Tsinaslanidis, G Upson, the Blood Transfusion laboratory team.

**Royal Surrey County Hospital NHS Foundation Trust** K McCullough (PI), H Abu, C Beazley, D Burda, P Bradley, B Creagh-Brown, J de Vos, S Donton, C Everden, J Fisher, H Gale, D Greene, O Hanci, L Harden, E Harrod, N Jeffreys, E Jones, J Jones, R Jordache, J Lawrence, C Marriott, I Mayanagao, R Mehra, N Michalak, O Mohamed, O Mohamed, S Mtuwa, A Nubi, K Odedra, C Piercy, V Pristopan, A Salberg, M Sanju, S Stone, J Verula, S Vimalanathan, the Blood Transfusion laboratory team.

**West Suffolk NHS Foundation Trust** M Moody (PI), J Balaam, S Barkha, J Barry, G Bass, H Cockerill, M Connolly, M El-Karim, J Godden, J Hoyle, J Kellett, I Lentell, T Murray, A Saraswatula, A Williams, L Wood, the Blood Transfusion laboratory team.

**George Eliot Hospital NHS Trust** S George (PI), V Bains, M Bryant, S Bukhari, Y Chang, Chaven, S Chucha, S Dean, L Dyble, N Elndari, H Gachi, J Glasgow, K Ellis, V Gulia, J Gunn, J Heys, C Ison, : K Jawaid, T Kannan, R Khan, D McDonald , R Musanhu, A Naeem, R Nair, N Navaneetham, S Nawaz, B Nazari, N Patel, W Penlington, G Ripley, G Roper, M Scally, E Sharrod , K Steinert, E Sung, D Suter, M Tahir, T Taylor, P Vanmali, B Vilcinskaite, E Vlad, R Walker, R Wookey, I Yusuf, the Blood Transfusion laboratory team.

**Yeovil District Hospital NHS Foundation Trust** A Broadley (PI), S Board, M Barnett, A Daxter, I Doig, A Getachew, A Hill, L Howard, R Jonnalagadda, A Kubisz-Pudelko, A Lewis, K Mansi, R Mason, A Melinte, B Mulhearn, J Reid, A Shah, R Smith, D Wood, the Blood Transfusion laboratory team.

**Dorset County Hospital NHS Foundation Trust** J Chambers (PI), J Birch, L Bough, J Colton, M Deighton, N Dewland, J Graves, M Greenslade, S Horton, L Poole, J Rees, R Thomas, W Verling, P Williams, S Williams, B Winter-Goodwin, S Wiseman, D Wixted, the Blood Transfusion laboratory team.

**Wye Valley NHS Trust** I DuRand (PI), J Al-Fori, J Annett, N Bray, A Carrasco, M Cohn, E Collins, K Crowley, A Davies, H Gashau, K Hammerton, I Hancock, A Hassan, A Hedges, R

Homewood, M Kevern, S Maryosh, S Meyrick, B Mwale, L Myslivecek, M Qayum, P Ryan, A Salam, C Seagrave, H Slade, F Suliman, A Talbot -Smith, S Turner, H Walker, J Woolley, the Blood Transfusion laboratory team.

**County Durham and Darlington NHS Foundation Trust** J Limb (PI), V Atkinson, M Birt, E Brown, A Cowton, V Craig, D Egginton, D Fernandes, J Gilbert, A Iqbal, A Ivy, D Jayachandran, J Jennings, A Kay, M Kent, J Lawson, S McAuliffe, C Morrell, S Naylor, G Nyamugunduru, J O'Brien, R Pipes, K Postlethwaite, K Potts, P Ranka, G Rogers, S Sen, J Temple, S Wadd, H Walters, S Watson, J Yorke, the Blood Transfusion laboratory team.

**Belfast HSC Trust** D Downey (PI), A Blythe, S Carr, A Cassells, D Comer, B Craig, D Dawson, K Hayes, C A Henry, R Ingham, L Keith, J Kidney, E Killen, J Leggett, D Linden, N Magee, R Marshall, D McClintock, M McFarland, L McGarvey, C McGettigan, S McGinnity, S McMahon, A Nugent, B O'Neill, A Redfern-Walsh, S Reilly, J Richardson, M Spence, R Stone, D Tweed, A UsherRea, B Wells, P Windrum, the Blood Transfusion laboratory team.

**NHS Grampian: Aberdeen Royal Infirmary** J Cooper (PI), R Soiza (Co-PI), V Bateman, M Black, R Brittain-Long, K Colville, D Counter, S Devkota, P Dospinescu, J Irvine, L Jappy, C Kaye, L Kane (Associate PI), A Khan, R Laing, J Lussier, A Mackenzie, M MacLeod, G Malik, M Mathie, J McLay, D Miller, K Norris, Y Pang, V Taylor, I Tonna, the Blood Bank Laboratory.

**Betsi Cadwaladr LHB: Ysbyty Gwynedd** C Subbe (PI), N Boyle, C Butterworth, C Hemmings, E Hughes, M Joishy, E Knights, J Leung, G Rieck, W Scrase, A Thomas, C Thorpe, the Blood Transfusion laboratory team.

**NHS Lanarkshire: University Hospital Monklands** M Patel (PI), C McGoldrick (Co-PI), P Anstey, T Baird, C Beith, A Blunsum, D Cairney, M Caldwell, A Crothers, S Dundas, F Farquhar, L Fraser, A Fyfe, L Glass, P Goyal, P Grant, S Howard, T Jones, J Lees, C Macrae, A McAlpine, A McCreath, C McInnes, C McKeag, A McKie, M McLaughlin, N Quail, M Ralston, J Reid, J Robb, S Rundell, A Stark, C Sykes, M Taylor, B Welsh, the Blood Transfusion laboratory team.

**NHS Fife** D Dhasmana (PI), P Marks, F Adam, K Aniruddhan, M Berwick, J Boyd, N Bulteel, P Cochrane, K Gray, L Hogg, S Iwanikiw, M Macmahon, P Marks, A McGregor, A Morrow, J Penman, J Pickles, A Scullion, H Sheridan, D Sloan, C Stewart, M Topping, S Whyte, the Blood Transfusion laboratory team.

**Royal Papworth Hospital NHS Foundation Trust** R Rintoul (PI), H Barker, F Bottrill, K Dorey, R Druyeh, M Earwaker, C East, S Fielding, C Flatters, A Fofana, C Freeman, C Galloway, L Garner, A Gladwell, U Hill, A Hudson, V Hughes, R Hussey, N Jones, J Joseph, P Madhivathanan, S Mepham, A Michael, M Muir, M Nizami, J Pack, K Paques, H Parfrey, G Polwarth, R Staples, J Quijano-Campos, S Victor, A Vuylsteke, S Webb, S Woods, K Wooodall, J Zamikula, the Biomedical Scientists and Associate Practitioners of the Transfusion laboratory.

**Salford Royal NHS Foundation Trust** P Dark (PI), N Bakerly, : L Baxter, B Blackledge, L Catlow, B Charles, M Collis, L Cooper, J Harris, A Harvey, K Knowles, S Lee, E Loutraris, T Marsden, L McMorrow, A Michael, J Pendlebury, J Perez, D Seddon, R Sukla, M Taylor, V Thomas, S Warran, J Wesson, Haematology and Blood Transfusion staff.

**The Rotherham NHS Foundation Trust** A Hormis (PI), C Bilton, C Brown, D Collier, J Field, J Ingham, S Lord, R Perkins, S Poku, S Sampath, R Stirk, R Walker, L Zeidan, the Blood Transfusion laboratory team.

**NHS Highland** B Sage (PI), F Barrett, W Beadles, C J Bradley, A Cochrane, R Cooper, A Goh, J Law, S Makin, J Matheson, D McDonald, C Millar, K Monaghan, L Murray, D Patience, V Rodrigues, G Simpson, SNBTS Inverness blood transfusion team.

**Taunton and Somerset NHS Foundation Trust** J Pepperell (PI), J Ashcroft, M Barnett, R Burgess, S Crouch, I Cruickshank, M Davey, T Dean, J Foot, L Graham, N Grieg, S Harlow, N Heydon, K James, K Johnson, C Lanaghan, D Lewis, A Locke, C Lorimer, J Lucas, H Mills, G Modgil, A Moss, M Nixon, S Northover, K O'Brien, I Parberry, K Roberts, J Rogers, C Susan, C Thompson, N Thorne, R Twemlow, C Vickers, R Wallbutton, A Whitcher, J Youens, E Zebracki, the Blood Transfusion laboratory team.

**The Dudley Group NHS Foundation Trust** H Ashby (PI), P Amy, S Ashman-Flavell, S de Silva, J Dean, N Fisher, E Forsey, J Frost, S Jenkins, A John, D Kaur, A Lubina Solomon, S Mahadevan-Bava, T Mahendiran, M O'Toole, S Pinches, D Rattehalli, U Sinha, A Smith, M Subramanian, C Tuckwell, J Vamvakopoulos, S Waidyanatha, the Blood Transfusion laboratory team.

**University Hospitals Of Morecambe Bay NHS Foundation Trust** S Bari (PI), A Higham (Co-PI), M Al-Jibury, K Allison, V Anu, C Bartlett, S Bhuiyan, L Bishop, K Burns, A Davies, A Fielding, M Gorst, C Hay, J Keating, T Khan, J Livingstone, F Mahmood, P Mallinder, S Peters, D Power, J Ritchie, K Simpson, H Spickett, C Stokes, E Stanton, H Thatcher, A Varghese, T Wan, R Williams, F Wood, the Blood Transfusion laboratory team.

**Betsi Cadwaladr LHB: Glan Clwyd Hospital** D Menzies (PI), A Abou-Haggar (Co-I), R Pugh (Co-I), N Abdallah, S Ambalavanan, K Darlington, F Davies, G Davis, I Davis, H Duvnjak, T Grenier, E Hall, L Hughes, M Joishy, R Lean, J Lewis, C Mackay, K Parvin, R Poyner, X Qui, S Rees, J Scanlon, N Sengupta, S Ullah, H Williams, the Blood Transfusion laboratory team.

**Airedale NHS Foundation Trust** T Gregory (PI), M Babirecki, H Bates, J Burnett, D Carr, E Dooks, F Farquhar, B Hairsine, A Hirst, A Imtiaz, T Knights, K Leighton, S Marsden (Associate PI), S Nallapeta, S Packham, N Yaqub, the Blood Transfusion laboratory team.

**The Queen Elizabeth Hospital, King's Lynn, NHS Foundation Trust** M Blunt (PI), S Abubacker, J Ali, C Atterbury, K Beaumont, K Bishop, H Bloxham, P Chan, Z Coton, H Curgenven, A Ebbs, M Elsaadany, T Fuller, M Iqbal, M Israa, S Jeddi, S Kamerkar, E Lim, E Nadar, K Naguleswaran, O Poluyi, H Rangarajan, G Rewitzky, S Ruff, S Shedwell, A Velusamy, H Webb, the Blood Transfusion laboratory team.

**Salisbury NHS Foundation Trust** M Sinha (PI), A Anthony, L Bell, S Diment, S Gray, S Haviland, A Hawkins, M Johns, V King, I Leadbitter, C Mathews, W Matimba-Mupaya, A Rand, S Salisbury, C Stacey, S Strong-Sheldrake, F Trim, the Blood Transfusion laboratory team.

**Brighton and Sussex University Hospitals NHS Trust** M Llewelyn (PI), H Brown, E Barbon, G Bassett, L Bennett, A Bexley, P Bhat, H Brown, Z Cipinova, J Cole, A Elkins, L Evans, J Gaylard, Z He, L Humber, S Jujjavarapu, C Laycock, D Mullan, J Newman, C Richardson, V Sellick, D Skinner, M Smith, D Yusef, R West, the Blood Transfusion laboratory team.

**NHS Borders: Borders General Hospital** A Scott (PI), S Alcorn, G Alcorn, J Aldridge, J Bain, C Barnetson, P Bell, A Campbell, B Chow, S Corbet, F Craighead, J Dawson, E Dearden, S Dickson, G Donaldson, T Downes, A Duncan, H El-Taweel, C Evans, T Fairbairn, L Finlayson, C Flanders, J Fletcher, J Foo, R Gallagher, N Hafiz, F Hall, M Herkes, M Hume, J Inglis, L Knox, J Lonnen, E MacVean, J Manning, H Matthews, A McLaren, C Murton, R Murton, B Muthukrishnan, E Nicol, F North, C Osbourne, R Porter, K Ralston, R Richmond, F Rodger, H Sakuri, B Soleimani, R Stewart, R Sutherland, A Tan, M Tolson, J Wilkie, R Williamson, the hospital Blood Transfusion team.

**Leeds Teaching Hospitals NHS Trust** J Minton (PI), S Ahmed, A Ashworth, A Ballinger, M Baum, L Bonney, J Calderwood, C Coupland, M Crow, M Davy, S Ferguson, S Hemphill, D Howarth, K Johnson, A Jones, M Kacar, M Karakantza, K Khokhar, P Lewthwaite, A Liversidge, F McGill, D Mistry, J Murira, Z Mustufvi, S O'Riordan, K Robinson, J Rock, S Straw, A Westwood, the Blood Transfusion laboratory team.

**Hywel Dda LHB: Prince Philip Hospital** S Ghosh (PI), K Lewis (Co-PI), S Coetzee, K Davies, S Griffiths, R Hicks, L Jones, L O'Brien, Z Omar, H Peregrine, Z Tariq, C Williams, C Woollard, the Blood Transfusion laboratory team.

**Maidstone and Tunbridge Wells NHS Trust** M Szeto (PI), K Cox (Co-PI), A Abbott, F Ajeneye, S Anandappa, B Babiker, M Barbosa, W Bonnert, C Boyd, G Chamberlain, D Datta, M Davey, R Gowda, S Goyal, A Gupta, E Harlock, C Hart, E Hughes, S Husaini, E Hutchinson, A Keough, A Khan, S Kumar, C Lawler, C Moore, R Nemane, L Orekoya, I Pamphlett, T Patel (Associate PI), C Pegg, P Rajagopalan, R Reilly, A Richards, S Sathianandan, R Seaman, S Siddavaram, E Small, O Solademi, P Tsang, A Waller, the Blood Transfusion laboratory team.

**NHS Greater Glasgow and Clyde: Inverclyde Royal Hospital** M Azharuddin (PI), H Papaconstantinou (Co-PI), D Cartwright, W Gallagher, T McClay, E Murray, O Olukoya, the Hospital Blood Transfusion team.

**Walsall Healthcare NHS Trust** A Garg (PI), K Shalan (PI), P Aiden Joseph, S Al-Hity, A Alina, B Allman, J Bearpark, S Bibi, A Bland, A Bowes, L Boyd, L Brown, W Campbell, Z Chandler, J Collins, M Dhanda, L Dwarakanath, A Farg, A Foot, A Gondal, T Gupta, T Jemima, B Jones, O Khan, J Khatri, R Krishnamurthy, H Mahmoud, R Marsh, R Mason, M Matonhodze, S Misra, J Muhammad, A Naqvi, C Newson, S Nortcliffe, S Odelberg, M Phipps, K Punia, H Qureshi, G Rajmohan, P Ranga, A Raymond-White, N Richardson, L Rogers, A Sheikh, A Srirajamadhuveeti, N Sunni, R Turel, E Virgilio, H Willis, F Wyn-Griffiths, the laboratory team.

**NHS Lanarkshire: University Hospital Hairmyres** M Patel (PI), F Burton (Co-PI), T McLennan (Co-PI), R Tejwani (Co-PI), D Bell, R Boyle, M Caldwell, L Clark, K Douglas, L Fraser, A Fyfe, L Glass, E Jarvie, E Lee, L Lennon, A McCreath, S Naidoo, the Blood Transfusion laboratory team.

**Mid Essex Hospital Services NHS Trust** A Hughes (PI), J Radhakrishnan (Co-PI), R Arnold, T Camburn, C Catley, E Dawson, C Fox, N Fox, H Gerrish, S Gibson, T Green, H Guth, F McNeela, T Nicholas, T Parker, A Rao, S Reid, B Singizi, S Smolen, S Williams, L Willsher, J Wootton, the Blood Transfusion laboratory team.

**Barnsley Hospital NHS Foundation Trust** K Inweregbu (PI), R Bowmer, R Colwell, A Daniels, J Emberey (Associate PI), C Green, R Gupta, L Harrison, A Hassan, S Hope, M Hussain, A Khalil, M Longshaw, S Meghjee, A Nicholson, A Sanderson, Y Sorour, C Swales, M Taplin, D Webster, the Blood Transfusion laboratory team.

**Royal Brompton & Harefield NHS Foundation Trust** A Shah (PI), A Reed (Co-PI), P Agent, A Angela, B Araba, L Banton, C Biring, H Day, P De Sousa, M Gaspar, V Jardim, V Jeyakumar, K Mahay, T Maria Pfyl, H Middleton, T Poonian, C Prendergast, P Rogers, G Sloane, N Soussi, V Teli, V Thwaiotes, J Visuvanathan, J Wallen, A Watson, the Blood Transfusion Laboratory teams.

**NHS Lanarkshire: University Hospital Wishaw** M Patel (PI), A Fyfe (Co-PI), A Smith (Co-PI), N Ali, K Black, R Boyle, M Caldwell, S Clements, J Fleming, L Fraser, A Fyfe, L Glass, L Hamilton, L Lennon, A Lynas, C Macdonald, M Maycock, A McCreath, E McGarry (Associate

PI), N Moody (Associate PI), Z Puyrigaud, C Stewart, P Wu, the Blood Transfusion laboratory team.

**Betsi Cadwaladr LHB: Wrexham Maelor Hospital** D Southern (PI), S Ahmer, R Bachar, C Bennett, G Bennett, L Brohan, T Coates, D Counsell, R Cowell, S Davies, J Harris, E Heselden, M Howells, R Hughes, S Kelly, J Kilbane, A Lloyd, H Maraj, A Mashta, G Mayers, R Petersen, H Reddy, L Richards, S Robertson, S Sandow, O Smith, S Smuts, G Spencer, G Szabo, S Thomas, S Thomas-Wright, S Tomlins, the Blood Transfusion laboratory team.

**The Princess Alexandra Hospital NHS Trust** U Ekeowa (PI), S Sakthi (Co-PI), Q Shah (Co-PI), M Anwar, B Badal, K Bumunarachchi, G Cook, A Daniel, K Dyer, A Easthope, J Finn, C Freer, A Gani, S Harris, E Haworth, L Hughes, K Ixer, N Konar, G Lucas, C McGuinness, C Muir, S Naik, T Nevin, O Newman, F Ozdes, R Ragatha, P Russell, R Saha, L Sandhu, E Shpuza, N Staines, S Waring, L Wee, T White, A Zahoor, the Blood Transfusion laboratory team.

**East Lancashire Hospitals NHS Trust** S Chukkambotla (PI), A Batista, H Collier, S Duberley, W Goddard, B Hammond, T Johnson, A Konstantinidis, L Mannion, K Marsden, A Mulla, A Newby, J Nugent, S Rigby, Rusk, M Sokolowski, C Spalding, A Sur, D Sutton, J Umeadi, the Blood Transfusion laboratory team.

**East Cheshire NHS Trust** X Lee (PI), L Alghazawi, M Babores, L Cunningham, C Gorman, A Gould, A Henderson, M Holland, R Hughes, L Huhn, M Husain, N Keenan, T Nagarajan, M O'Brien, L Paisley, H Wassall, L Wilkinson, K Wolffsohn, the Blood Transfusion laboratory team.

**Great Ormond Street Hospital For Children NHS Foundation Trust** M Peters (PI), A Bamford (Co-PI), L Grandjean, O Akinkugbe, H Belfield, P Eyton-Jones, J Hassell, G Jones, T McHugh, R Moss, L O'Neill, J Penner, A Tomas, the Blood Transfusion laboratory team.

**Northern Devon Healthcare NHS Trust** R Manhas (PI), K Allen, F Bellis, H Black, D Dalton, D Davies, B-L Halls, R Hartley, M Jeelani, M Lamparski, E Lekoudis, S Ley, B Rowlands, H Thomas, A Umeh, K Wedgeworth, E Willis, the Blood Transfusion laboratory team.

**Royal Cornwall Hospitals NHS Trust** D Browne (PI), Z Berry, H Chenoweth, A Collinson, F Hammonds, L Jones, E Laity, T Nisbett, A Parsons, R Sargent, I Sullivan, K Watkins, L Welch, the Blood Transfusion laboratory team.

**Homerton University Hospital NHS Foundation Trust** K Woods (PI), A Claxton (Co-PI), Y Akinfenwa, N Aladangady, A Begum, R Brady, L Canclini, S Capocci, A Chiapparino, R Corser, M Elmi, E Fonyonga, R Frowd, C Gorman, C Holbrook, M Jagpal, S Jain, J Kaur, A Martinez, J Miah, R Mullett, C Nimmo (Associate PI), C Quah, P Reynolds, T Tanqueray, L Terry, E Timlick, the Blood Transfusion laboratory team.

**Hywel Dda LHB: Bronglais General Hospital** M Hobrok (PI), A Snell (Co-PI), D Asandei, B Atkins, S Jenkins, K Jones, K Khan, R Loosley, J Markham, H McGuinness, D McKeogh, A Mohamed, L Raisova, H Tench, W Wolf, R Wolf-Roberts, the Blood Transfusion laboratory team.

**Hywel Dda LHB: Withybush Hospital** J Green (PI), R Hughes, G Lewis, C Macphee, A Meess, H Thomas, H Tollick, the Blood Transfusion laboratory team

**Alder Hey Children's NHS Foundation Trust** D Hawcutt (PI), K Allison, J Fu, L O'Malley, S Owens, L Rad, G Seddon, T Shackleton.

**Birmingham Women's and Children's NHS Foundation Trust** K Morris (PI), D Jyothish (PI), E Al-Abadi (Co-PI), S Hartshorn (Co-PI), H Krishnan (Co-PI), R Cole, J Groves, K Hong, J Jackson, C O'Hara , H Sohal , S Sultan, M Wakelin, H Williamson , H Winmill.

**Hywel Dda LHB: Glangwili General Hospital** K Lewis (PI), S Griffiths, R Hicks, L Jones; H Peregrine, Z Tariq, the Blood Transfusion laboratory team.

**Lewisham and Greenwich NHS Trust** S Kegg (PI), A Aghababaie, H Azzoug, E Bates, M Chakravorty, K Chan, P Coakley, E Gardiner, A Hastings, D Jegede, A-M Johnson, J Juhl, T Kelani, M Magriplis, C Milliken, J Muglu, D Mukimbiri, M Nadheem, T Nair, M Nyirenda, T OConnor, T Ogbara, R Olaiya, C Onyeagor, V Palaniappan, A Pieris, S Pilgrim, P Richards, T Simpson, A Taylor, K Wesseldine, M Woodman, E Woolley, the laboratory team.

**Liverpool Women's NHS Foundation Trust** R McFarland (PI), M Dower, C Dragomir, S Holt, K Knowles, H Mcnamara, C Morgan, E Neary, A Smith, M Turner.

**NHS Forth Valley: Forth Valley Royal Hospital** M Spears (PI), A Baggott, K Bohmova, G Clark, J Donnachie, S Huda, G Jayasekera, I Macpherson, M Maycock, N McInnes, J McMinn, D Morrison, A Pearson, L Prentice, C Rafique, D Salutous, M Stewart, R Thompson, A Todd, P Turner.

**The Christie NHS Foundation Trust** V Kasipandian (PI), A Binns, S Jackson, J King, P Mahjoob-Afag, R Mary-Genetu, P Nicola, A Patel, D Seals, R Shotton, D Sutinyte.

**The Walton Centre NHS Foundation Trust** R Davies (PI), H Arndt, A Clyne, E Hetherington, G Hull, J McEnerney, R Wright.

**Western HSC Trust** M Kelly (PI), D Concannon, A Crawford, K Ferguson, L Gelmon, J Kara, C Kelly, C Kerlin, D McClintock, T McManus, M P McNicholl, J Monaghan, B O'Neill, J Pastrana, N Smyth, J Wieboldt, the Haematology/Blood Transfusion/Central Specimen Reception laboratory teams.

**Chelsea and Westminster Hospital NHS Foundation Trust** P Shah (PI), B Mann (Co-PI), K Alatzoglou, K Alizaedeh, M Al-Obaidi, K Ballard, A Barker, C Bautista, M Boffito, M Bourke, R Bull, C Caneja, J Carungcong, F Conway, P Costa, E Dwyer, C Fernandez, J Garner, J Girling, E Hamlyn, A Holyome, L Horsford, N Hynes, M Johnson, N Kazi, U Kirwan, S Maheswaran, M Martineau, T Ngan, K Nundlall, C Orton, T Peters, A Sayan, A Schoolmeesters, M Svensson, A Tana, A Thayanandan, J Tonkin, B Vijayakumar, C Winpenny, O Zibdeh.

**Dragon's Heart Hospital** J Coulson (PI).

**East and North Hertfordshire NHS Trust** P Ferranti (PI), M Boampoaa, M Chaudhury, C Cruz, M Ebon, J Edmonds, M Erotocritou, A Fajardo, A Frosh, S Gelves-Zapata, D Gorog, T Ingle, J Kefas, E Malundas, C Matei, J Mathers, L Miller (Associate PI), I Nadeem (Associate PI), Y Odedina, D Palit, L Peacock, S Tewari (Associate PI), L Ventilacion, R Zill-E-Huma, E Zinkin (Associate PI).

**Gateshead Health NHS Foundation Trust** R Allcock (PI), D Mansour (PI), M Armstrong, J Barbour, M Bokhar, J Caulfield, J Curtis, A Dale, V Deshpande, I Hashmi, E Johns, L Jones, R King, E Ladlow (Associate PI), R Mackie, D Mansour, B McClelland, W McCormick, C McDonald, C Moller-Christensen, R Petch, S Razvi, R Sharma, L Southern, G Stiller, E Watkins, H Wilkins.

**NHS Ayrshire and Arran: University Hospital Ayr** K Walker (PI), C Burns, D Callaghan, R Cuthbertson, K Gibson, D Gilmour, M Henry, J Locke, L McNeil, S Meehan, A Murphy, K Naismith, K Prasad, M Rodger, C Turley, S Walton, M Wilson, S Wood.

**NHS Golden Jubilee National Hospital** B Shelley (PI), V Irvine, F Thompson.

**NHS Tayside: Ninewells Hospital** J Chalmers (PI), H Abo-Leyah, C Deas, H Loftus, A Nicoll, A Strachan, J Taylor, C Tee.

**NHS Western Isles** G Stanczuk (PI), M Murdoch (Co-PI), A Apostolopoulos, A Carstairs, S Klaczek.

**North West Boroughs Healthcare NHS Foundation Trust** A Baldwin (PI).

**Sheffield Children's NHS Foundation Trust** P Avram (PI), A Bellini, A Bellini, F Blakemore, H Chisem, J Clements, H Cook, S Gormely, D Hawley, C Kerrison, N Lawrence, G Margabanthu, A McMahon, N Roe, M Scott, F Shackley, J Sowter, T Williams.

**South Warwickshire NHS Foundation Trust** S Tso (PI), P Parsons (Co-PI), S Bird, B Campbell, G Kakoullis, F Mackie, C O'Brien, T Taylor, K Webb, A Smith.

**The Royal Marsden NHS Foundation Trust** K Tatham (PI), P Angelini, E Bancroft, E Black, A Dela Rosa, E Durie, M Hogben, S Jhanji, I Leslie, A Okines, I Sana, S Shepherd, N Taylor, S Wong.

**Velindre NHS Trust** J Powell (PI), R Adams (Co-PI), A Jackson.

**West Hertfordshire Hospitals NHS Trust** V Page (PI), R Vancheeswaran, L Norris, X Zhao, A Yousafzar.

### Supplementary Methods

#### Study organization

The RECOVERY trial is an investigator-initiated, individually randomised, open-label, controlled trial to evaluate the efficacy and safety of a range of putative treatments in patients hospitalized with COVID-19. The protocol is available at www.recoverytrial.net. The trial was conducted at 176 National Health Service (NHS) hospital organizations in the United Kingdom. The trial was coordinated by a team drawn from the Clinical Trial Service Unit and the National Perinatal Epidemiology Clinical Trials Unit within the Nuffield Department of Population Health at University of Oxford, the trial sponsor. Support for local site activities was provided by the National Institute for Health Research Clinical Research Network.

Access to relevant routine health care and registry data was supported by NHS DigiTrials, the Intensive Care National Audit and Research Centre, Public Health Scotland, National Records Service of Scotland, and the Secure Anonymised Information Linkage (SAIL) at University of Swansea.

#### Collection and distribution of convalescent plasma

Supply of convalescent plasma to local sites was coordinated and managed by the four blood services (NHS Blood and Transplant, Northern Ireland Blood Transfusion Service, Scottish National Blood Transfusion Service, and Welsh Blood Service) according to the standard donor selection guidelines in the UK [https://www.transfusionguidelines.org/dsg]. Convalescent plasma was collected via apheresis from individuals with self-reported symptoms consistent with COVID-19 or a laboratory confirmed SARS-CoV-2 infection, in both cases at least 28 days after the symptom resolution. The median time between symptom onset or test-positivity and donation was 54 days (IQR 41-78).

All convalescent plasma donations collected in England, Northern Ireland, Scotland and Wales were routinely tested for SARS-CoV-2 IgG antibodies using the EUROimmun IgG enzyme-linked immunosorbent assay (ELISA) test targeting the spike (S) glycoprotein (PerkinElmer, London, UK) at the four UK blood services. Only donations with sample to cut-off (S/CO) ratio of 6.0 or above were supplied for clinical trial use, as this cut-off was previously demonstrated to be associated with the presence of neutralising antibody titres of ≥1:100.[1,2] Among first-time donors with PCR-confirmed infection not leading to hospitalisation, 1,382/5,906 (23%) had Euroimmun S/CO values above this threshold.

#### Measurement of participant baseline SARS-CoV-2 serostatus

The presence of SARS-CoV-2 IgG antibodies in serum samples taken at randomization was determined using a validated high throughput ELISA, the Oxford immunoassay. This is an indirect ELISA, measuring serum IgG against trimeric spike protein using a horseradish peroxidase-linked anti-human IgG antibody as the secondary. Readouts are measured as a fluorescent signal, normalised to standard units by calibrating to a dilution series of NHSBT plasma controls and a monoclonal antibody (CR3022) run on each plate. A full description of the assay and its evaluation can be found in the report by the National Sars-CoV-Serology Assay Evaluation Group.[3]

#### Protocol changes

RECOVERY is a randomised trial among patients hospitalized for COVID-19. All eligible patients receive usual standard of care in the participating hospital and are randomly allocated between no additional treatment and one of several active treatment arms. Over time, additional treatment arms have been added (see Table).

The original and final protocol relevant to convalescent plasma are included in the supplementary material to this publication, together with summaries of the changes made.

**Table. Protocol changes to treatment comparisons**

| Protocol version | Date | Randomisation | Treatment arms |
|---|---|---|---|
| 1.0 | 13-Mar-2020 | Main (part A) | No additional treatment<br>Lopinavir-ritonavir[a]<br>Low-dose corticosteroid[b]<br>Nebulised Interferon-ß-1a (never activated) |
| 2.0 | 23-Mar-2020 | Main (part A) | No additional treatment<br>Lopinavir-ritonavir[a]<br>Low-dose corticosteroid[b]<br>Hydroxychloroquine |
| 3.0 | 07-Apr-2020 | Main (part A) | No additional treatment<br>Lopinavir-ritonavir[a]<br>Low-dose corticosteroid[b]<br>Hydroxychloroquine[c]<br>Azithromycin[d] |
| 4.0 | 14-Apr-2020 | Main (part A)<br><br><br><br><br><br>Second[e,f] | No additional treatment<br>Lopinavir-ritonavir[a]<br>Low-dose corticosteroid[b]<br>Hydroxychloroquine[c]<br>Azithromycin[d]<br><br>No additional treatment<br>Tocilizumab |
| 5.0 | 24-Apr-2020 | - | (no change – extension to children <18 years old) |
| 6.0 | 14-May-2020 | Main (part A)<br><br><br><br><br><br>Main (part B factorial)<br><br><br>Second[e,f] | No additional treatment<br>Lopinavir-ritonavir[a]<br>Low-dose corticosteroid[b]<br>Hydroxychloroquine[c]<br>Azithromycin[d]<br><br>No additional treatment<br>Convalescent plasma[h]<br><br>No additional treatment<br>Tocilizumab |
| 7.0 | 18-Jun-2020 | Main (part A)<br><br><br><br><br>Main (part B factorial)<br><br><br>Second[e,f] | No additional treatment<br>Lopinavir-ritonavir[a]<br>Low-dose corticosteroid[b]<br>Azithromycin[d]<br><br>No additional treatment<br>Convalescent plasma[h]<br><br>No additional treatment<br>Tocilizumab |

| Protocol version | Date | Randomisation | Treatment arms |
|---|---|---|---|
| 8.0 | 03-Jul-2020 | Main (part A)<br><br><br><br><br><br>Main (part B factorial)<br><br><br>Second[e,f] | No additional treatment<br>Low-dose corticosteroid[b]<br>Intravenous immunoglobulin[g]<br>High-dose corticosteroid[g]<br>Azithromycin[d]<br><br>No additional treatment<br>Convalescent plasma[h]<br><br>No additional treatment<br>Tocilizumab |
| 9.1 | 18-Sep-2020 | Main (part A)<br><br><br><br><br><br>Main (part B factorial)<br><br><br><br>Second[e,f] | No additional treatment<br>Low-dose corticosteroid[b]<br>Intravenous immunoglobulin[g]<br>High-dose corticosteroid[g]<br>Azithromycin[d]<br><br>No additional treatment<br>Convalescent plasma[h]<br>REGN-COV2<br><br>No additional treatment<br>Tocilizumab |
| 10.1 | 01-Nov-2020 | Main (part A)<br><br><br><br><br><br>Main (part B factorial)<br><br><br><br>Main (part C factorial)<br><br><br>Second[e,f] | No additional treatment<br>Low-dose corticosteroid[b]<br>Intravenous immunoglobulin[g]<br>High-dose corticosteroid[g]<br>Azithromycin[d]<br><br>No additional treatment<br>Convalescent plasma[h]<br>REGN-COV2<br><br>No additional treatment<br>Aspirin<br><br>No additional treatment<br>Tocilizumab |

| Protocol version | Date | Randomisation | Treatment arms |
|---|---|---|---|
| 11.1 | 27-Nov-2020 | Main (part A) | No additional treatment<br>Low-dose corticosteroid[b]<br>Intravenous immunoglobulin[g]<br>High-dose corticosteroid[g]<br>Colchicine |
| | | Main (part B factorial) | No additional treatment<br>Convalescent plasma[h]<br>REGN-COV2 |
| | | Main (part C factorial) | No additional treatment<br>Aspirin |
| | | Second[e,f] | No additional treatment<br>Tocilizumab |
| 12.1 | 16-Dec-2020 | Main (part A)[h] | No additional treatment<br>Low-dose corticosteroid[b]<br>Intravenous immunoglobulin[g]<br>High-dose corticosteroid[g]<br>Colchicine |
| | | Main (part B factorial)[i] | No additional treatment<br>Convalescent plasma[h]<br>REGN-COV2 |
| | | Main (part C factorial)[i] | No additional treatment<br>Aspirin |
| | | Second[e,f] | No additional treatment<br>Tocilizumab |

[a] enrolment ceased 29 June 2020 when the Data Monitoring Committee advised that the Chief Investigators should review the unblinded data.
[b] enrolment of adults ceased 8 June 2020 as more than 2,000 patients had been recruited to the active arm
[c] enrolment ceased 5 June 2020 when the Data Monitoring Committee advised that the Chief Investigators should review the unblinded data.
[d] enrolment of adults ceased 27 November 2020 as more than 2,500 patients had been recruited to the active arm
[e] for patients with (a) oxygen saturation <92% on air or requiring oxygen or children with significant systemic disease with persistent pyrexia; and (b) C-reactive protein ≥75 md/L)
[f] enrolment of adults ceased 24 January 2021 as more than 2,000 patients had been recruited to the active arm.
[g] for children only
[h] enrolment of adults ceased 15 January 2021 when the Data Monitoring Committee advised that the Chief Investigators should review the unblinded data.
[i] from protocol version 12.1, children could enter the second randomisation regardless of whether they were included in the main randomisation

**Main and second randomisation <u>for adults</u>**

All RECOVERY trial participants received usual standard of care. On study entry, adult participants initially underwent the Main Randomisation. Trial participants with clinical evidence of progressive COVID-19 (defined as oxygen saturation <92% on room air or requiring oxygen therapy, and C-reactive protein ≥75 mg/L) could be considered for the Second Randomisation at any time up to 21 days after the initial randomisation, and regardless of initial treatment allocation(s). A web-system was used to provide simple randomisation (without stratification or minimisation) with allocation concealment until randomisation had been completed.

Over time, treatment arms were added and removed from the protocol, factorial randomisations were introduced (see below), and not all treatments were available at every hospital. Similarly, not all treatments were deemed by the attending clinician to be suitable for some patients (e.g. due to comorbid conditions or concomitant medication). In any of these cases, randomisation involved fewer arms (and/or fewer factorial elements).

*Main randomisation for adults*

A single participant could be randomised at most to 1 arm from each of part A, B, and C of the three factorial randomisations, and thus receive 0, 1, 2, or 3 treatments on top of usual standard of care.

<u>Part A</u> (from 19 March 2020)

Eligible participants could be randomised to one of the following arms:

| Treatment arm | Arm opened | Arm closed |
|---|---|---|
| No additional treatment | 19 March 2020 | Ongoing |
| Dexamethasone | 19 March 2020 | 8 June 2020 |
| Lopinavir-ritonavir | 19 March 2020 | 29 June 2020 |
| Hydroxychloroquine | 23 March 2020 | 5 June 2020 |
| Azithromycin | 7 April 2020 | 27 November 2020 |
| Colchicine | 27 November 2020 | Ongoing |

<u>Part B</u> (from 14 May 2020)

Eligible participants could be randomised to one of the following arms:

| Treatment arm | Arm opened | Arm closed |
|---|---|---|
| No additional treatment | 14 May 2020 | Ongoing |
| Convalescent plasma | 14 May 2020 | 15 January 2021 |
| REGN-COV2* | 18 September 2020 | Ongoing |

* monoclonal neutralising antibody cocktail

<u>Part C</u> (from 1 November 2020)

Eligible participants could be randomised to one of the following arms:

| Treatment arm | Arm opened | Arm closed |
|---|---|---|
| No additional treatment | 1 November 2020 | Ongoing |
| Aspirin | 1 November 2020 | Ongoing |

*Second randomisation for adults (from 14 April 2020)*

From 14 April 2020, a participant could be randomised to one of the following arms and thus receive 0 or 1 treatment on top of those allocated in the initial randomisation and usual standard of care:

| Treatment arm | Arm opened | Arm closed |
|---|---|---|
| No additional treatment | 14 April 2020 | 24 Janaury 2021 |
| Tocilizumab | 14 April 2020 | 24 Janaury 2021 |

**Supplementary statistical methods**

*Sample size*

As stated in the protocol, appropriate sample sizes could not be estimated when the trial was being planned at the start of the COVID-19 pandemic. As the trial progressed, the Trial Steering Committee, blinded to the results of the study treatment comparisons, formed the view that sufficient patients should be enrolled to each comparison to provide at least 90% power at two-sided P=0.01 to detect a proportional reduction in 28-day mortality of one-fifth. For example, if 28-day mortality was 20% then a comparison of at least 2500 participants allocated to active drug and 2500 to usual care alone would suffice.

For the antibody therapies (convalescent plasma and REGN-COV2), data from external sources emerged during the trial. These suggested that clinical response to these treatments may be greater among those patients who did not have existing anti-SARS-CoV-2 antibodies prior to treatment.[4-6] Consequently while still blind to the results, the RECOVERY trial steering committee determined the trial should enrol sufficient patients to provide adequate power (90% at 2P=0.01) to detect an effect of convalescent plasma among those patients without known positive antibody levels at baseline.

The Trial Steering Committee determined that recruitment to this comparison should continue until at least 5,000 patients without known positive antibodies (and 11,000 patients in total) had been randomised. If 28-day mortality in those allocated to usual care was 20%, then a study of this size would have 90% power at two-sided P=0.01 to detect a proportional reduction of one-fifth in this subgroup.

**Interim analyses: role of the Data Monitoring Committee**

The independent Data Monitoring Committee reviews unblinded analyses of the study data and any other information considered relevant at intervals of around 2 weeks. The committee is charged with determining if, in their view, the randomised comparisons in the study provide evidence on mortality that is strong enough (with a range of uncertainty around the results that was narrow enough) to affect national and global treatment strategies. In such a circumstance, the Committee would inform the Steering Committee who would make the results available to the public and amend the trial arms accordingly. Unless that happened, the Steering Committee, investigators, and all others involved in the trial would remain blind to the interim results until 28 days after the last patient had been randomised to a particular intervention arm. Further details about the role and membership of the independent Data Monitoring Committee are provided in the protocol.

The Data Monitoring Committee determined that to consider recommending stopping a treatment early for benefit would require at least a 3 to 3·5 standard error reduction in mortality. The Committee concluded that examinations of the data at every 10% (or even 5%) of the total data would lead to only a marginal increase in the overall type I error rate.

**Ascertainment and classification of study outcomes**

Information on baseline characteristics and study outcomes was collected through a combination of electronic case report forms (see below) completed by members of the local research team at each participating hospital and linkage to National Health Service, clinical audit, and other relevant health records. Full details are provided in the RECOVERY Definition and Derivation of Baseline Characteristics and Outcomes Document (see Appendix 3).

Thrombotic events were defined as either (i) acute pulmonary embolism; (ii) deep vein thrombosis; (iii) ischaemic stroke; (iv) myocardial infarction; or (v) systemic arterial embolism.

Clinically significant bleeding was defined as intracranial bleeding or bleeding requiring transfusion, endoscopy, surgery, or vasoactive drugs.

*Randomisation form*

The Randomisation form (shown below) was completed by trained study staff. It collected baseline information about the participant (including demographics, COVID-19 history, comorbidities and suitability for the study treatments) and availability of the study treatments. Once completed and electronically signed, the treatment allocation was displayed.

The following modifications were made to the Randomisation form during the trial:

| Randomisation form version | Date of release | Major modifications from previous version |
|---|---|---|
| 1.0 | 19-Mar-20 | Initial version (protocol V1.0) |
| 2.0 | 25-Mar-20 | For protocol V2.0<br>• Hydroxycholoroquine added as treatment<br>• Known long QT syndrome added to comorbidities<br>• Severe depression removed from comorbidities |
| 3.0 | 09-Apr-20 | For protocol V3.0<br>• Azithromycin added as treatment<br>• Suspected SARS-CoV-2 infection included in eligibility criteria |
| [Second randomisation form introduced] | 23-Apr-20 | For protocol 4.0<br>• Eligibility criteria for second randomisation<br>• Tocilizumab vs control as treatment allocations |
| 4.0 | 09-May-20 | For protocol V5.0<br>• Age ≥18 years removed from eligibility criteria<br>• Additional questions on child's age and weight added |
| 5.0 | 21-May-20 | For protocol V6.0<br>• Convalescent plasma added as treatment |
| 6.0 | 28-May-20 | Baseline use of remdesivir |
| 7.0 | 01-Jul-20 | For protocol V7.0<br>• Participants eligible if convalescent plasma is only available and suitable treatment<br>• Hydroxychloroquine, dexamethasone (adults) and lopinavir-ritonavir removed |

| **Randomisation form version** | **Date of release** | **Major modifications from previous version** |
|---|---|---|
| 8.0 | 13-Aug-20 | For protocol V8.0<br>• Addition of low-dose and high-dose corticosteroids and intravenous immunoglobulin for children (and removal of dexamethasone for children) |
| 9.0 | 24-Sep-20 | For protocol V9.0<br>• REGN-COV2 added as treatment<br>• Additional baseline information |
| 10.0 | 06-Nov-20 | For protocol V10.1<br>• Aspirin added as treatment |
| 11.0 | 27-Nov-20 | For protocol V11.1<br>• Colchicine added as treatment<br>• Azithromycin removed |
| 12.0 | 22-Dec-20 | For protocol V12.1<br>• Allow children to enter trial without entering main randomisation |

RECOVERY
Randomised Evaluation of COVID-19 Therapy

#### Sample Form (v12.00 - 17/12/20)

#### Randomisation Program

Call Freefone **0800 138 5451** to contact the RECOVERY team for **URGENT** problems using the Randomisation Program or for medical advice. All **NON-URGENT queries** should be emailed to ****

Logged in as: **RECOVERY Site**

##### Section A: Baseline and Eligibility

Date and time of randomisation: 17 Dec 2020 14:00

**Treating clinician**

**A1.** Name of treating clinician

**Patient details**

**A2.** Patient surname

Patient forename

**A3.** NHS number ☐ Tick if not available

**A4.** What is the patient's date of birth? 01 / January / 2000

**A5.** What is the patient's sex?

**Inclusion criteria**

**A6.** Has consent been taken in line with the protocol?
If answer is No patient cannot be enrolled in the study

**A7.** Does the patient have proven or suspected SARS-CoV-2 infection?
If answer is No patient cannot be enrolled in the study

**A8.** Does the patient have any medical history that might, in the opinion of the attending clinician, put the patient at significant risk if they were to participate in the trial?

**A8B.** Is the patient willing to receive convalescent plasma?

**A9.** COVID-19 symptom onset date: / /

**A10.** Date of hospitalisation: / /

**A11.** Does the patient require oxygen?

**A12.** Please select one of the following to describe the current level of ventilation support

**A12.1** Enter latest oxygen saturation measurement (%)

**A12.2** Enter latest CRP measurement since admission to hospital (mg/L) ☐ Tick if not measured
Enter 0 if below the limit of measurement ☐ Tick if greater than limit of measurement

**A12.3** Enter latest creatinine measurement since admission to hospital (μmol/L) ☐ Tick if not measured

**A12.4** Enter latest D-dimer measurement since admission to hospital (ng/mL) ☐ Tick if not measured
Enter 0 if below the limit of measurement ☐ Tick if greater than limit of measurement

**A12.5** Has the patient received a COVID-19 vaccine?

**Does the patient have any CURRENT comorbidities or other medical problems or treatments?**

**A13.1** Diabetes

**A13.2** Heart disease

**A13.3** Chronic lung disease

**A13.4** Tuberculosis

**A13.5** HIV

**A13.6** Severe liver disease

**A13.7** Severe kidney impairment (eGFR<30 or on dialysis)

**A13.8** Known long QT syndrome

**A13.9** Current treatment with macrolide antibiotics which are to continue
Macrolide antibiotics include clarithromycin, azithromycin and erythromycin

**A13.10** Antiplatelet therapy
Includes aspirin, clopidogrel, ticagrelor, prasugrel, dipyridamole

**A13.11** Previous adverse reaction to blood or blood product transfusion

**Are the following treatments UNSUITABLE for the patient?**
**If you answer Yes it means you think this patient should NOT receive this drug.**

**A14.3** Colchicine

**A14B.1** Convalescent plasma

**A14B.2** Synthetic monoclonal antibodies (REGN10933+REGN10987)

**A14C.1** Aspirin

**Are the following treatments available?**

**A15.3** Colchicine

**A15B.1** Convalescent plasma

**A15B.2** Synthetic monoclonal antibodies (REGN10933+REGN10987)

**A15C.1** Aspirin

**Current medication**

**A16.1** Is the patient currently prescribed remdesivir?

**A16.2** Is the patient currently prescribed systemic corticosteroids (dexamethasone, prednisolone, hydrocortisone, methylprednisolone)?
Please do not include topical or inhaled treatments

**A16.4** Is the patient currently on warfarin or a direct oral anticoagulant?
Includes apixaban, rivaroxaban

**A16.5** What venous thromboembolism prophylaxis is the patient receiving?
Standard = usual for hospitalised patients (not increased due to COVID-19); Higher dose = treatment dose or increased prophylaxis due to COVID-19

**Please sign off this form once complete**

Surname:

Forename:

Professional email:

Continue

Cancel

Home

*72 hour Follow-up Form*

Using an online form (shown on the next page), information on events occurring within the first 72 hours after randomisation for participants in the antibody-based therapy comparisons was collected by trained study staff.

The following modifications were made to the 72 hour follow-up form during the trial:

| Form version | Date of release | Major modifications from previous version |
|---|---|---|
| 1.0 | 28-May-20 | Initial version (protocol V5.0) |
| 2.0 | 03-Jul-20 | For protocol V8.0<br>• Additional detail collected on certain events |
| 3.0 | 24-Sep-20 | For protocol V9.0<br>• Addition of REGN-COV2 |

### Early Safety Data

**This form should NOT be completed less than 72 hours after randomisation (unless the patient has been discharged or died)**

**Patient's date of birth** *

yyyy-mm-dd

#### Convalescent Plasma

**1. How many convalescent plasma infusions did the patient receive?** *

*This is plasma given as part of a trial, not any standard fresh frozen plasma or other blood products that the patient may have been given*

◯ 0 ◯ 1 ◯ 2

**A patient is not expected to receive both a convalescent plasma and a synthetic monoclonal antibodies infusion. Please check your answers.**

**1.1. Were any infusions stopped early for any reason ie, the patient did not receive the full amount?**

◯ Yes ◯ No

**1.2. How many were stopped early?**

◯ 1 ◯ 2

#### Synthetic monoclonal antibodies (REGN10933+REGN10987)

**1.B. Did the patient receive an infusion of REGN10933+REGN10987?** *

◯ Yes ◯ No

**A patient is not expected to receive both a convalescent plasma and a synthetic monoclonal antibodies infusion. Please check your answers.**

| **Date infusion started** | **Time infusion started** |
| --- | --- |
| yyyy-mm-dd | *(hh:mm [24 hr])* |

**The infusion should not start before randomisation. Please check your answers.**

**The infusion is expected to start within 48 hours of randomisation. Please check your answers.**

**1.B.1. Was the infusion stopped early for any reason ie, the patient did not receive the full amount?**

- ◯ Yes
- ◯ No

**1.B.2. Did the patient have a reaction during the infusion?**

- ◯ Yes
- ◯ No

**1.B.3. How was the reaction managed?**

*Please tick all that apply*

- ☐ No intervention required
- ☐ Infusion rate reduced, but infusion completed
- ☐ Antihistamine given
- ☐ Steroid given
- ☐ Adrenaline given
- ☐ Infusion stopped early

#### First 72 hours after the first randomisation

**2. During the first 72 hours after the first randomisation has the patient had any of the following?**

**2.1. Sudden worsening in respiratory status** *

- ◯ Yes
- ◯ No
- ◯ Unknown

**Please indicate the respiratory support delivered**

*Please tick all that apply*

- ☐ No additional support
- ☐ New use or increased concentration of oxygen
- ☐ New use of non-invasive respiratory support (CPAP, BiPAP, HFNO)
- ☐ New use of invasive mechanical ventilation
- ☐ Other

**Please provide details**

**Persistent change**

- ◯ Please indicate if persistent change (ie, increased support still required at 72 hours)

**2.2. Severe allergic reaction** *

- ◯ Yes
- ◯ No
- ◯ Unknown

**Please indicate if adrenaline was required?**

Yes No

#### 2.3. Temperature >39˚C or ≥2˚C rise above baseline *

Yes No Unknown

#### 2.4. Sudden hypotension *

*Defined as either (i) sudden drop in systolic blood pressure of ≥30 mmHg with systolic blood pressure ≤80 mmHg; or (ii) requiring urgent medical attention*

Yes No Unknown

#### Please indicate support given

*Please tick all that apply*

- [ ] No support required
- [ ] New or additional intravenous fluid
- [ ] New or additional inotropic/vasopressor support

#### Persistent change

Please indicate if persistent change (ie, increased support still required at 72 hours)

#### 2.5. Clinical haemolysis *

*Defined as fall in haemoglobin plus one or more of the following: rise in lactate dehydrogenase (LDH), rise in bilirubin, positive direct antiglobulin test (DAT), or positive crossmatch*

Yes No Unknown

#### Haemoglobin

Please indicate if the lowest haemoglobin <100 g/L

#### Bilirubin

Please indicate if the highest bilirubin >50 μmol/L

#### 2.6. Thrombotic event *

*Defined as either (i) acute pulmonary embolism; or (ii) deep-vein thrombosis; or (iii) ischaemic stroke; or (iv) myocardial infarction; or (v) systemic arterial embolism*

Yes No Unknown

#### Please indicate the type of thrombotic event

*Please tick all that apply*

- [ ] Acute pulmonary embolism
- [ ] Deep-vein thrombosis
- [ ] Ischaemic stroke
- [ ] Myocardial infarction
- [ ] Systemic arterial embolism
- [ ] Other

#### Please provide details

**2.7. Was a SHOT report submitted?**

○ Yes ○ No ○ Unknown

**3. Please enter any additional information**

*Main follow-up form*

The Main Follow-up form (shown on the next page) collected information on study treatment adherence (including both the randomised allocation and use of other study treatments), vital status (including date and provisional cause of death if available), hospitalisation status (including date of discharge), respiratory support received during the hospitalisation, occurrence of any major cardiac arrhythmias and renal replacement therapy received. Questions on thrombotic and bleeding events were added with V10.1 of the protocol so these data were collected for only a small proportion of people in the convalescent plasma comparison.

The following modifications were made to the Follow-up form during the trial:

| **Follow-up form version** | **Date of release** | **Modifications from previous version** |
|---|---|---|
| 1.0 | 30-Mar-20 | Initial version |
| 2.0 | 09-Apr-20 | Information on other treatments used during admission:<br>• Azithromycin, IL-6 receptor antagonist<br>Fact and result of SARS-CoV-2 PCR test |
| 3.0 | 09-Apr-20 | Update to functionality; no changes to questions |
| 4.0 | 23-Apr-20 | Duration of treatments added |
| 5.0 | 12-May-20 | Capture of major cardiac arrhythmias added |
| 6.0 | 28-May-20 | Updates to wording of questions.<br>Information on other treatments used during admission:<br>• Remdesivir, convalescent plasma |
| 7.0 | 18-Jun-20 | Clarification of question wording |
| 8.0 | 10-Jul-20 | Information on new treatments for children adherence |
| 9.0 | 24-Sep-20 | Information on REGN-COV2 adherence |
| 10.0 | 06-Nov-20 | Information on aspirin adherence<br>Capture of thrombotic and bleeding events added<br>Information of enrolment into other studies added |
| 11.0 | 16-Nov-20 | Minor changes to in-form validation |
| 12.0 | 27-Nov-20 | Information on colchicine adherence |

### Follow-up

#### Date of randomisation

**Please only report events that occurred from first randomisation until 28 days later on this form (except for Q2).**

**Patient's date of birth** *

yyyy-mm-dd

**1. Which of following treatment(s) did the patient definitely receive as part of their hospital admission after randomisation?** *

*(NB Include RECOVERY study-allocated drug, only if given, PLUS any of the other treatments if given as standard hospital care)*

- ☐ No additional treatment
- ☐ Lopinavir-ritonavir
- ☐ Corticosteroid (dexamethasone, prednisolone, hydrocortisone or methylprednisolone)
- ☐ Hydroxychloroquine
- ☐ Azithromycin or other macrolide (eg, clarithromycin, erythromycin)
- ☐ Tocilizumab or sarilumab
- ☐ Remdesivir
- ☐ Intravenous immunoglobulin
- ☐ Synthetic monoclonal antibodies (REGN10933+REGN10987)
- ☐ Aspirin
- ☐ Colchicine

**Please select number of days the patient received lopinavir-ritonavir**

○ 1 ○ 2 ○ 3 ○ 4 ○ 5 ○ 6 ○ 7 ○ 8 ○ 9 ○ 10

**Please select number of days the patient received corticosteroid (dexamethasone, prednisolone, hydrocortisone or methylprednisolone)**

○ 1 ○ 2 ○ 3 ○ 4 ○ 5 ○ 6 ○ 7 ○ 8 ○ 9 ○ 10

**Please select number of days the patient received hydroxychloroquine**

○ 1 ○ 2 ○ 3 ○ 4 ○ 5 ○ 6 ○ 7 ○ 8 ○ 9 ○ 10

**Please select number of days the patient received azithromycin**

○ 0 ○ 1 ○ 2 ○ 3 ○ 4 ○ 5 ○ 6 ○ 7 ○ 8 ○ 9 ○ 10

**Please select number of days the patient received other macrolides (eg, clarithromycin, erythromycin)**

○ 0 ○ 1 ○ 2 ○ 3 ○ 4 ○ 5 ○ 6 ○ 7 ○ 8 ○ 9 ○ 10

**Please select number of doses of tocilizumab or sarilumab the patient received**

○ 1 ○ >1

**Please select number of days the patient received remdesivir**

○ 1 ○ 2 ○ 3 ○ 4 ○ 5 ○ 6 ○ 7 ○ 8 ○ 9 ○ 10

**Please select the proportion of days the patient received aspirin or other antiplatelet (eg, clopidogrel, prasugrel, ticagrelor, dipyridamole) during the first 28 days after randomisation (or from randomisation to date of discharge if this is sooner)**

○ Most days (≥90%) ○ Some days (≥50% <90%) ○ Few days (<50% of days, but not zero) ○ None

**Please select number of days the patient received colchicine**

○ 1 ○ 2 ○ 3 ○ 4 ○ 5 ○ 6 ○ 7 ○ 8 ○ 9 ○ 10

#### » Convalescent Plasma

**How many convalescent plasma infusions did the patient receive?**

*This is convalescent plasma (i.e. collected from people recovered from COVID-19), not any standard fresh frozen plasma or other blood products that the patient may have been given*

○ 0 ○ 1 ○ 2

**Were any infusions stopped early for any reason ie, the patient did not receive the full amount?**

○ Yes ○ No

**How many were stopped early?**

○ 1 ○ 2

#### » Health Status

**2. Was a COVID-19 test done for this patient at any point during the admission?** *

*(If multiple tests were done, and the results were positive and negative, please tick Yes – positive result and Yes – negative result)*

☐ Yes – positive result

☐ Yes – negative result

☐ Not done

**3. What is the patient's vital status?** *

- Alive
- Dead

**3.1 What is the patient's current hospitalisation status?** *

- Inpatient
- Discharged

The patient has been enrolled in the trial for **NaN** days

**3.1.1 Date follow-up form completed**

yyyy-mm-dd

**3.1.1 What was the date of discharge?** *

yyyy-mm-dd

**3.1 What was the date of death?** *

yyyy-mm-dd

**3.2 What was the underlying cause of death?** *

*This can be obtained from the last entry in part 1 of the death certificate*

- COVID-19
- Other infection
- Cardiovascular
- Other

**Please give details**

**4. Did the patient require any form of assisted ventilation (ie, more than just supplementary oxygen) from day of randomisation until 28 days later?** *

- Yes
- No

**Please answer the following questions:**

**4.1 For how many days did the patient require assisted ventilation?** *

**4.2 What type of ventilation did the patient receive?**

| | Yes | No | Unknown |
|---|---|---|---|
| **CPAP alone** | ○ | ○ | ○ |
| **Non-invasive ventilation (eg, BiPAP)** | ○ | ○ | ○ |
| **High-flow nasal oxygen (eg, AIRVO)** | ○ | ○ | ○ |
| **Mechanical ventilation (intubation/tracheostomy)** | ○ | ○ | ○ |
| **ECMO** | ○ | ○ | ○ |

**Total number of days the patient received invasive mechanical ventilation (intubation/tracheostomy) from randomisation until discharge/death/28 days after randomisation**

**5. Has the patient been documented to have a NEW cardiac arrhythmia at any point since the main randomisation until 28 days later?** *

- ○ Yes
- ○ No
- ○ Unknown

**5.1 Please select all of the following which apply**

- ☐ Atrial flutter or atrial fibrillation
- ☐ Supraventricular tachycardia
- ☐ Ventricular tachycardia (including torsades de pointes)
- ☐ Ventricular fibrillation
- ☐ Atrioventricular block requiring intervention (eg, cardiac pacing)

**6. Did the patient require use of renal dialysis or haemofiltration from main randomisation until 28 days later?** *

- ○ Yes
- ○ No

**7. During the first 28 days after randomisation (or until discharge if sooner), did the participant have a thrombotic event?** *

- Yes
- No
- Unknown

**7.1 Please indicate the type of thrombotic event**

*Select all that apply*

- Pulmonary embolism
- Deep-vein thrombosis
- Ischaemic stroke
- Myocardial infarction
- Systemic arterial embolism
- Other

**8. During the first 28 days after randomisation (or until discharge if sooner), did the participant experience clinically-significant bleeding ie, intra-cranial bleeding or bleeding that required intervention (eg, surgery, endoscopy or vasoactive drugs) or a blood transfusion?** *

- Yes
- No
- Unknown

**8.1 Please indicate the site(s) of bleeding** *

*Select all that apply*

- Intra-cranial
- Gastrointestinal
- Other

**8.2 Please indicate which interventions were required to manage the bleed** *

*Select all that apply*

- Blood transfusion
- Surgery
- Endoscopy
- Vasoactive drugs (e.g. inotropes on ICU)
- None of the the above

**9. Please indicate if the participant participated in any other COVID-19 trials**

*Select all that apply*

- PRINCIPLE

- [ ] PRINCIPLE
- [ ] REMAP-CAP
- [ ] Other treatment trial(s)
- [ ] COVID-19 vaccine trial(s)

**Please give name of other treatment trial(s)**

**Please give name of COVID-19 vaccine trial(s)**

**10. Please enter UKOSS case ID if known** *

*Enter the full UKOSS case ID ie, COR_123*

*(select if you do not know the UKOSS case ID)*

- [ ] Not known

### Supplementary Tables

**Webtable 1: Baseline characteristics of patients considered unsuitable for randomisation to convalescent plasma compared with those randomised to convalescent plasma versus usual care.**

| | Randomised (n=11558) | Considered Unsuitable (n=2764) |
|---|---|---|
| Age, years | 63.5 (14.7) | 61.6 (17.9) |
| <70 | 7453 (64) | 1783 (65) |
| 70 to 79 | 2590 (22) | 566 (20) |
| ≥80 | 1515 (13) | 415 (15) |
| Sex | | |
| Men | 7430 (64) | 1646 (60) |
| Women | 4128 (36) | 1118 (40) |
| Ethnicity | | |
| White | 8655 (75) | 1818 (66) |
| Black, Asian, and minority ethnic | 1742 (15) | 556 (20) |
| Unknown | 1161 (10) | 390 (14) |
| Number of days since symptom onset | 9 (6-12) | 9 (6-12) |
| Number of days since admission to hospital | 2 (1-3) | 2 (1-4) |
| Respiratory support received | | |
| No oxygen received | 897 (8) | 371 (13) |
| Oxygen only* | 10044 (87) | 2194 (79) |
| Invasive mechanical ventilation | 617 (5) | 199 (7) |
| Previous diseases | | |
| Diabetes | 3104 (27) | 699 (25) |
| Heart disease | 2576 (22) | 649 (23) |
| Chronic lung disease | 2713 (23) | 605 (22) |
| Tuberculosis | 43 (<1) | 7 (<1) |
| HIV | 36 (<1) | 11 (<1) |
| Severe liver disease† | 142 (1) | 30 (1) |
| Severe kidney impairment‡ | 616 (5) | 157 (6) |
| Any of the above | 6425 (56) | 1458 (53) |
| SARS-CoV-2 PCR test result | | |
| Positive | 11140 (96) | 2570 (93) |
| Negative | 238 (2) | 133 (5) |
| Unknown | 180 (2) | 61 (2) |
| Patient SARS-CoV-2 antibody test result | | |
| Positive | 5774 (50) | 376 (14) |
| Negative | 3611 (31) | 230 (8) |
| Missing | 2173 (19) | 2158 (78) |
| Corticosteroids received | | |
| Yes | 10681 (92) | 2410 (87) |
| No | 804 (7) | 329 (12) |
| Not recorded | 73 (1) | 25 (1) |
| Other randomised treatments | | |
| Lopinavir-ritonavir | 19 (<1) | 7 (<1) |
| Dexamethasone | 6 (<1) | 2 (<1) |
| Hydroxychloroquine | 1 (<1) | 0 |
| Azithromycin | 1172 (10) | 314 (11) |
| Colchicine | 1583 (14) | 472 (17) |
| Aspirin | 2473 (21) | 651 (24) |

Data are mean (SD), n (%), or median (IQR). * Includes non-invasive ventilation. † Defined as requiring ongoing specialist care. ‡ Defined as estimated glomerular filtration rate <30 mL/min per 1·73 $m^2$

**Webtable 2: Baseline characteristics by patient baseline antibody status and randomised allocation.**

| | Seronegative | | Seropositive | | Antibody status unknown | |
|---|---|---|---|---|---|---|
| | Convalescent Plasma (n=1982) | Usual Care (n=1629) | Convalescent Plasma (n=3022) | Usual Care (n=2752) | Convalescent Plasma (n=791) | Usual Care (n=1382) |
| Age, years | 65.4 (15.4) | 66.0 (15.1) | 62.1 (13.8) | 61.5 (13.6) | 64.6 (15.6) | 63.9 (15.5) |
| <70 | 1133 (57) | 924 (57) | 2102 (70) | 1969 (72) | 470 (59) | 855 (62) |
| 70 to 79 | 487 (25) | 406 (25) | 641 (21) | 566 (21) | 182 (23) | 308 (22) |
| ≥80 | 362 (18) | 299 (18) | 279 (9) | 217 (8) | 139 (18) | 219 (16) |
| Sex | | | | | | |
| Men | 1183 (60) | 1011 (62) | 1973 (65) | 1874 (68) | 487 (62) | 902 (65) |
| Women | 799 (40) | 618 (38) | 1049 (35) | 878 (32) | 304 (38) | 480 (35) |
| Ethnicity | | | | | | |
| White | 1577 (80) | 1307 (80) | 2190 (72) | 1947 (71) | 595 (75) | 1039 (75) |
| Black, Asian, and minority ethnic | 229 (12) | 159 (10) | 507 (17) | 481 (17) | 117 (15) | 249 (18) |
| Unknown | 176 (9) | 163 (10) | 325 (11) | 324 (12) | 79 (10) | 94 (7) |
| Number of days since symptom onset | 7 (5-10) | 7 (4-9) | 10 (7-13) | 10 (7-13) | 8 (5-11) | 8 (5-12) |
| Number of days since admission to hospital | 1 (1-3) | 1 (1-3) | 2 (1-4) | 2 (1-4) | 2 (1-4) | 2 (1-4) |
| Respiratory support received | | | | | | |
| No oxygen received | 222 (11) | 171 (10) | 129 (4) | 129 (5) | 91 (12) | 155 (11) |
| Oxygen only* | 1713 (86) | 1415 (87) | 2675 (89) | 2422 (88) | 663 (84) | 1156 (84) |
| Invasive mechanical ventilation | 47 (2) | 43 (3) | 218 (7) | 201 (7) | 37 (5) | 71 (5) |
| Previous diseases | | | | | | |
| Diabetes | 561 (28) | 454 (28) | 775 (26) | 727 (26) | 199 (25) | 388 (28) |
| Heart disease | 539 (27) | 464 (28) | 534 (18) | 501 (18) | 194 (25) | 344 (25) |
| Chronic lung disease | 575 (29) | 500 (31) | 617 (20) | 525 (19) | 193 (24) | 303 (22) |
| Tuberculosis | 10 (1) | 11 (1) | 8 (<1) | 10 (<1) | 2 (<1) | 2 (<1) |
| HIV | 4 (<1) | 6 (<1) | 10 (<1) | 8 (<1) | 3 (<1) | 5 (<1) |
| Severe liver disease† | 35 (2) | 19 (1) | 21 (1) | 35 (1) | 14 (2) | 18 (1) |
| Severe kidney impairment‡ | 156 (8) | 98 (6) | 115 (4) | 101 (4) | 52 (7) | 94 (7) |
| Any of the above | 1234 (62) | 1027 (63) | 1525 (50) | 1405 (51) | 444 (56) | 790 (57) |
| SARS-CoV-2 PCR test result | | | | | | |
| Positive | 1909 (96) | 1563 (96) | 2950 (98) | 2688 (98) | 722 (91) | 1308 (95) |
| Negative | 50 (3) | 34 (2) | 27 (1) | 27 (1) | 48 (6) | 52 (4) |
| Unknown | 23 (1) | 32 (2) | 45 (1) | 37 (1) | 21 (3) | 22 (2) |
| Corticosteroids received | | | | | | |
| Yes | 1812 (91) | 1497 (92) | 2886 (95) | 2616 (95) | 672 (85) | 1198 (87) |
| No | 167 (8) | 131 (8) | 133 (4) | 131 (5) | 91 (12) | 151 (11) |
| Not recorded | 3 (<1) | 1 (<1) | 3 (<1) | 5 (<1) | 28 (4) | 33 (2) |
| Other randomised treatments | | | | | | |
| Lopinavir-ritonavir | 0 | 0 | 0 | 0 | 5 (1) | 14 (1) |
| Dexamethasone | 0 | 0 | 0 | 0 | 3 (<1) | 3 (<1) |
| Hydroxychloroquine | 0 | 0 | 0 | 0 | 1 (<1) | 0 |
| Azithromycin | 210 (11) | 171 (10) | 279 (9) | 275 (10) | 98 (12) | 139 (10) |
| Colchicine | 263 (13) | 214 (13) | 436 (14) | 394 (14) | 93 (12) | 183 (13) |
| Aspirin | 406 (20) | 312 (19) | 723 (24) | 605 (22) | 137 (17) | 290 (21) |

Data are mean (SD), n (%), or median (IQR). * Includes non-invasive ventilation. † Defined as requiring ongoing specialist care. ‡ Defined as estimated glomerular filtration rate <30 mL/min per 1·73 $m^2$

#### Webtable 3: Treatments given, by randomised allocation

| | Convalescent Plasma (n=5795) | Usual Care (n=5763) |
|---|---|---|
| Infusions of convalescent plasma received | | |
| None | 449 (8%) | 5718 (99%) |
| One | 671 (12%) | 6 (0%) |
| Two | 4675 (81%) | 39 (1%) |
| Follow-up form received | 5458 | 5443 |
| Other treatments received | | |
| Corticosteroid | 4545 (83%) | 4508 (83%) |
| Lopinavir-Ritonavir | 5 (<1%) | 17 (<1%) |
| Hydroxychloroquine | 11 (<1%) | 13 (<1%) |
| Azithromycin or other macrolides | 1933 (35%) | 1962 (36%) |
| Tocilizumab or sarilumab | 421 (8%) | 555 (10%) |
| Remdesivir | 1741 (32%) | 1721 (32%) |

Percentages are of those with complete follow-up data available.

#### Webtable 4: Cause-specific 28-day mortality

| Cause of death | Convalescent Plasma (n=5795) | Usual Care (n=5763) | Absolute percent difference (SE) |
|---|---|---|---|
| COVID | 1282 (22%) | 1280 (22%) | -0.1 (0.77) |
| Other infection | 7 (<1%) | 9 (<1%) | 0.0 (0.07) |
| Cardiac | 6 (<1%) | 11 (<1%) | -0.1 (0.07) |
| Stroke | 6 (<1%) | 4 (<1%) | 0.0 (0.05) |
| Other vascular | 2 (<1%) | 1 (<1%) | 0.0 (0.03) |
| Cancer | 11 (<1%) | 12 (<1%) | 0.0 (0.08) |
| Other medical | 36 (1%) | 42 (1%) | -0.1 (0.15) |
| External | 1 (<1%) | 0 (0%) | 0.0 (0.02) |
| Unknown cause | 47 (1%) | 49 (1%) | 0.0 (0.17) |
| Total: 28-day mortality | 1398 (24%) | 1408 (24%) | -0.3 (0.80) |

#### Webtable 5: Adverse events at 72 hours after randomisation

| | Convalescent Plasma (n=5795) | Usual Care (n=5763) |
|---|---|---|
| Number with form completed | 5267 | 5128 |
| Sudden worsening in respiratory status | | |
| No additional support required | 66 (1%) | 68 (1%) |
| New or increased use of O2 | 681 (13%) | 660 (13%) |
| New non-invasive respiratory support | 447 (8%) | 435 (8%) |
| New invasive mechanical ventilation | 163 (3%) | 160 (3%) |
| Other | 32 (1%) | 22 (<1%) |
| Total: Any sudden worsening in respiratory status | 1130 (21%) | 1132 (22%) |
| Persistent worsening | 729 (14%) | 725 (14%) |
| Severe allergic reaction | | |
| Adrenaline required | 0 (0%) | 1 (<1%) |
| Any severe allergic reaction | 16 (<1%) | 2 (<1%) |
| Temperature >39C or >=2C rise above baseline | 195 (4%) | 168 (3%) |
| Sudden hypotension | | |
| No support required | 68 (1%) | 58 (1%) |
| New or additional intravenous fluid | 29 (1%) | 40 (1%) |
| New or additional inotropic/vasopressor support | 33 (1%) | 53 (1%) |
| Total: Any sudden hypotension | 127 (2%) | 140 (3%) |
| Persistent change | 33 (1%) | 50 (1%) |
| Clinical haemolysis | | |
| Haemoglobin <100 g/L | 45 (1%) | 26 (1%) |
| Bilirubin >50 umol/L | 8 (<1%) | 4 (<1%) |
| Total: Clinical haemolysis | 87 (2%) | 66 (1%) |
| Thrombotic event | | |
| Acute pulmonary embolism | 53 (1%) | 59 (1%) |
| Deep-vein thrombosis | 8 (<1%) | 8 (<1%) |
| Ischaemic stroke | 3 (<1%) | 5 (<1%) |
| Myocardial infarction | 11 (<1%) | 3 (<1%) |
| Systemic arterial embolism | 1 (<1%) | 1 (<1%) |
| Other | 6 (<1%) | 10 (<1%) |
| Total: Any thrombotic event | 73 (1%) | 87 (2%) |

* Percentages are of those with a completed 72-hr safety form.

### Webtable 6: Effect of allocation to convalescent plasma on major cardiac arrythmia

| | Convalescent Plasma (n=5795) | Usual Care (n=5763) |
|---|---|---|
| Number with follow-up form | 5458 | 5443 |
| Atrial flutter or atrial fibrillation | 166 (3%) | 181 (3%) |
| Other supraventricular tachycardia | 33 (1%) | 48 (1%) |
| Subtotal: Supraventricular tachycardia | 191 (3%) | 218 (4%) |
| Ventricular tachycardia | 16 (<1%) | 20 (<1%) |
| Ventricular fibrillation | 5 (<1%) | 2 (<1%) |
| Subtotal: Ventricular tachycardia or fibrillation | 21 (<1%) | 22 (<1%) |
| Atrioventricular block requiring intervention | 5 (<1%) | 6 (<1%) |
| Total: Any major cardiac arrhythmia | 215 (4%) | 247 (5%) |

* Percentages are of those with such a form completed.

#### Webtable 7: Primary, secondary and subsidiary outcomes, after adjustment for the sex imbalance at randomisation

| | Convalescent plasma (n=5795) | Usual Care (n=5763) | RR (95% CI) | p value |
|---|---|---|---|---|
| **Primary outcome** | | | | |
| Mortality at 28 days | 1398 (24%) | 1408 (24%) | 1.00 (0.93-1.08) | 0.93 |
| **Secondary outcomes** | | | | |
| Discharged from hospital within 28 days | 3850 (66%) | 3846 (67%) | 0.98 (0.94-1.03) | 0.42 |
| Invasive mechanical ventilation or death* | 1561/5493 (28%) | 1561/5448 (29%) | 1.00 (0.94-1.06) | 0.96 |
| Invasive mechanical ventilation | 670/5493 (12%) | 681/5448 (13%) | 0.99 (0.89-1.09) | 0.78 |
| Death | 1240/5493 (23%) | 1263/5448 (23%) | 0.98 (0.91-1.05) | 0.56 |
| **Subsidiary outcomes** | | | | |
| Use of ventilation † | 860/3564 (24%) | 863/3441 (25%) | 0.97 (0.90-1.05) | 0.49 |
| Non-invasive ventilation | 822/3564 (23%) | 821/3441 (24%) | 0.98 (0.90-1.06) | 0.57 |
| Invasive mechanical ventilation | 226/3564 (6%) | 237/3441 (7%) | 0.93 (0.78-1.11) | 0.42 |
| Successful cessation of invasive mechanical ventilation ‡ | 87/302 (29%) | 112/315 (36%) | 0.77 (0.58-1.02) | 0.06 |
| Renal replacement therapy § | 258/5729 (5%) | 249/5713 (4%) | 1.05 (0.89-1.24) | 0.58 |

Data are n (%) or n/N (%). RR=sex-adjusted rate ratio for the outcomes of 28-day mortality, hospital discharge, and successful cessation of invasive mechanical ventilation, and sex-adjusted risk ratio for other outcomes.
* Analyses exclude those on invasive mechanical ventilation at randomisation.
† Analyses exclude those on invasive or non-invasive ventilation at randomisation.
‡ Analyses exclude those not receiving invasive mechanical ventilation at randomisation.
§ Analyses exclude those on renal replacement therapy at randomisation.

### Supplementary Figures

#### Webfigure 1: Mortality at 28 Days, According to by Retrospectively−Defined Categories of Days Since Onset, Respiratory Support Received and Date of Randomisation

| | Convalescent plasma | Usual care | RR (95% CI) |
|---|---|---|---|
| **Days since symptom onset ($\chi_1^2$=2.8; p=0.09)** | | | |
| ≤4 | 268/933 (29%) | 305/975 (31%) | 0.91 (0.77−1.08) |
| 5−7 | 338/1293 (26%) | 354/1265 (28%) | 0.93 (0.80−1.09) |
| 8−11 | 409/2001 (20%) | 388/1974 (20%) | 1.05 (0.92−1.21) |
| ≥12 | 380/1563 (24%) | 361/1548 (23%) | 1.07 (0.92−1.23) |
| **Respiratory support received ($\chi_1^2$= 3.5; p=0.06)** | | | |
| No oxygen received | 56/442 (13%) | 69/455 (15%) | 0.83 (0.58−1.18) |
| Oxygen without NIV | 521/3122 (17%) | 523/2986 (18%) | 0.96 (0.85−1.08) |
| Oxygen (NIV unknown) | 69/262 (26%) | 56/259 (22%) | 1.26 (0.89−1.80) |
| Oxygen with NIV | 594/1667 (36%) | 615/1748 (35%) | 1.04 (0.93−1.16) |
| Invasive mechanical ventilation | 158/302 (52%) | 145/315 (46%) | 1.19 (0.95−1.50) |
| **Date of Randomisation ($\chi_1^2$=0.3; p=0.56)** | | | |
| Before 1st Dec 2020 | 623/2716 (23%) | 639/2703 (24%) | 0.97 (0.87−1.09) |
| On or after 1st Dec 2020 | 775/3079 (25%) | 769/3060 (25%) | 1.02 (0.92−1.12) |
| **All participants** | **1398/5795 (24%)** | **1408/5763 (24%)** | **1.00 (0.93−1.07) p=0.93** |

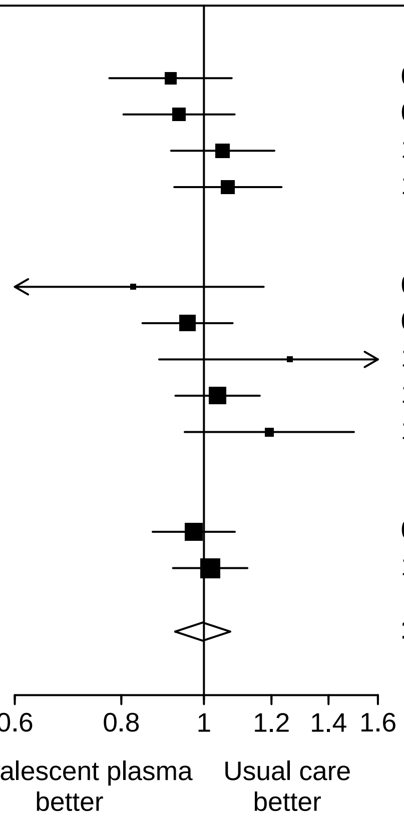

### Webfigure 2: Mortality at 28 days, According to Patient Antibody Status

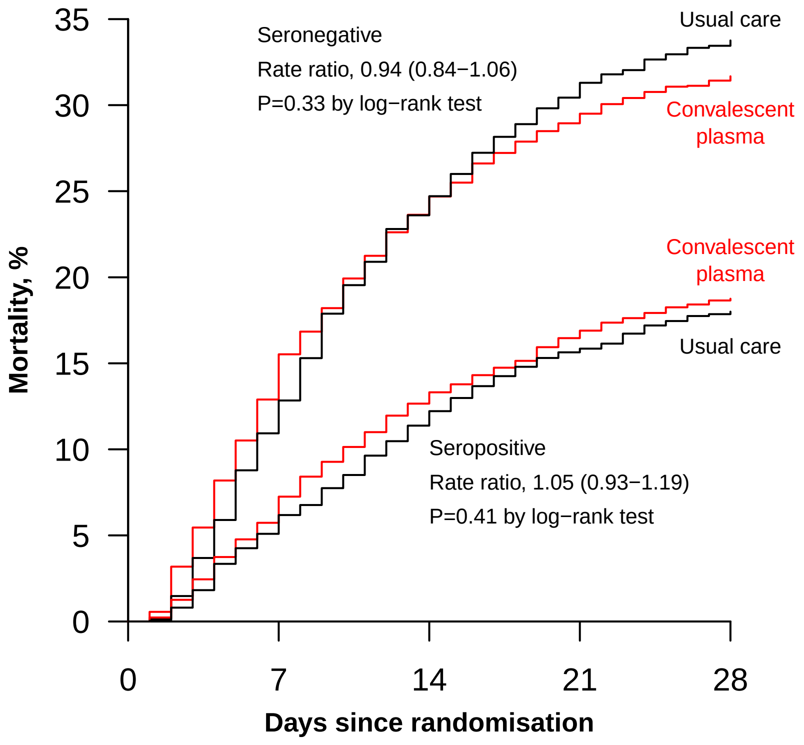

| No. at risk, Seronegative | | | | | |
|---|---|---|---|---|---|
| Convalescent plasma | 1982 | 1669 | 1487 | 1392 | 1349 |
| Usual Care | 1629 | 1418 | 1223 | 1116 | 1076 |

| No. at risk, Seropositive | | | | | |
|---|---|---|---|---|---|
| Convalescent plasma | 3022 | 2799 | 2616 | 2508 | 2452 |
| Usual Care | 2752 | 2580 | 2414 | 2314 | 2255 |

#### Webfigure 3: Discharged Alive within 28 Days, According to Subgroup

| | Convalescent plasma | Usual care | RR (95% CI) |
|---|---|---|---|
| **Age, years ($\chi^2_1$=1.3; p=0.25)** | | | |
| <70 | 2802/3705 (76%) | 2851/3748 (76%) | 0.97 (0.92−1.03) |
| 70 to 79 | 705/1310 (54%) | 683/1280 (53%) | 1.03 (0.92−1.14) |
| ≥80 | 343/780 (44%) | 312/735 (42%) | 1.05 (0.90−1.22) |
| **Sex ($\chi^2_1$= 0.3; p=0.61)** | | | |
| Men | 2342/3643 (64%) | 2467/3787 (65%) | 0.97 (0.92−1.03) |
| Women | 1508/2152 (70%) | 1379/1976 (70%) | 1.00 (0.92−1.07) |
| **Ethnicity ($\chi^2_1$=1.1; p=0.30)** | | | |
| White | 2897/4362 (66%) | 2858/4293 (67%) | 0.98 (0.93−1.04) |
| Black, Asian or minority ethnic | 561/853 (66%) | 611/889 (69%) | 0.92 (0.82−1.03) |
| **Days since symptom onset ($\chi^2_1$= 2.8; p=0.10)** | | | |
| ≤7 | 1403/2226 (63%) | 1379/2240 (62%) | 1.03 (0.96−1.12) |
| >7 | 2446/3564 (69%) | 2466/3522 (70%) | 0.95 (0.90−1.01) |
| **Respiratory support received ($\chi^2_1$=3.9; p=0.05)** | | | |
| No oxygen received | 361/442 (82%) | 353/455 (78%) | 1.11 (0.95−1.30) |
| Oxygen only | 3436/5051 (68%) | 3425/4993 (69%) | 0.97 (0.93−1.02) |
| Invasive mechanical ventilation | 53/302 (18%) | 68/315 (22%) | 0.79 (0.55−1.13) |
| **Use of corticosteroids ($\chi^2_1$=2.2; p=0.14)** | | | |
| Yes | 3568/5370 (66%) | 3553/5311 (67%) | 0.98 (0.93−1.02) |
| No | 267/391 (68%) | 268/413 (65%) | 1.12 (0.94−1.33) |
| **Patient SARS−CoV−2 antibody test result ($\chi^2_1$=8.9; p=0.003)** | | | |
| Positive | 2192/3022 (73%) | 2051/2752 (75%) | 0.93 (0.87−0.99) |
| Negative | 1159/1982 (58%) | 903/1629 (55%) | 1.09 (1.00−1.19) |
| Not done | 499/791 (63%) | 892/1382 (65%) | 0.99 (0.88−1.11) |
| **All participants** | **3850/5795 (66%)** | **3846/5763 (67%)** | **0.98 (0.94−1.03) p=0.50** |

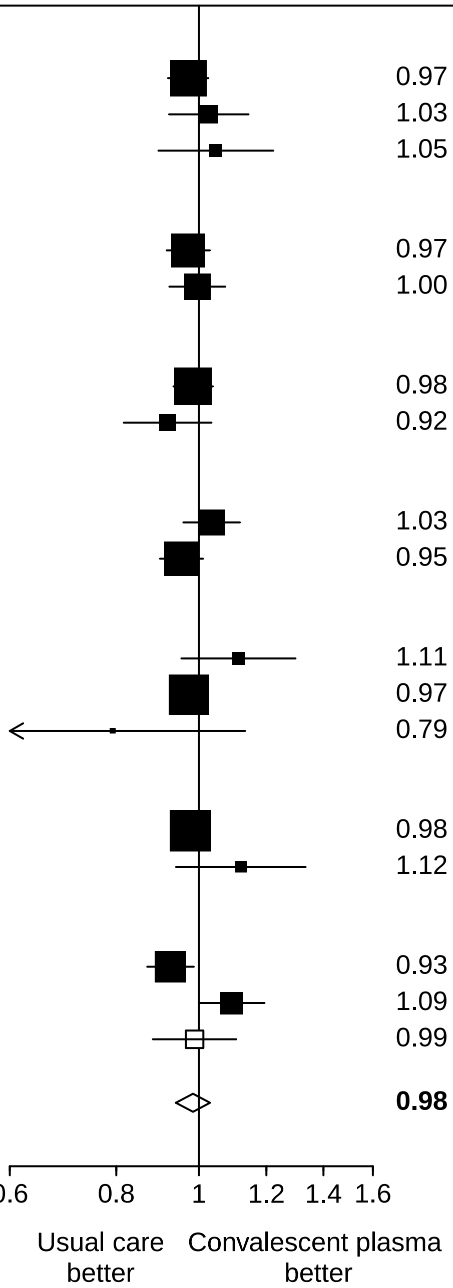

### Webfigure 4: Invasive Mechanical Ventilation or Death, According to Subgroup

| | Convalescent plasma | Usual care | RR (95% CI) |
|---|---|---|---|
| **Age, years ($\chi^2_1$=0.9; p=0.34)** | | | |
| <70 | 697/3471 (20%) | 694/3501 (20%) | 1.01 (0.92−1.11) |
| 70 to 79 | 495/1247 (40%) | 499/1215 (41%) | 0.97 (0.88−1.06) |
| ≥80 | 369/775 (48%) | 368/732 (50%) | 0.95 (0.85−1.05) |
| **Sex ($\chi^2_1$= 0.5; p=0.47)** | | | |
| Men | 1064/3441 (31%) | 1088/3563 (31%) | 1.01 (0.94−1.09) |
| Women | 497/2052 (24%) | 473/1885 (25%) | 0.97 (0.87−1.08) |
| **Ethnicity ($\chi^2_1$=0.1; p=0.80)** | | | |
| White | 1237/4203 (29%) | 1214/4122 (29%) | 1.00 (0.94−1.07) |
| Black, Asian or minority ethnic | 217/762 (28%) | 224/804 (28%) | 1.02 (0.87−1.20) |
| **Days since symptom onset ($\chi^2_1$= 3.3; p=0.07)** | | | |
| ≤7 | 690/2149 (32%) | 741/2156 (34%) | 0.93 (0.86−1.02) |
| >7 | 867/3339 (26%) | 820/3291 (25%) | 1.04 (0.96−1.13) |
| **Respiratory support received ($\chi^2_1$=1.4; p=0.24)** | | | |
| No oxygen received | 60/442 (14%) | 75/455 (16%) | 0.82 (0.60−1.13) |
| Oxygen only | 1501/5051 (30%) | 1486/4993 (30%) | 1.00 (0.94−1.06) |
| **Use of corticosteroids ($\chi^2_1$=3.4; p=0.07)** | | | |
| Yes | 1484/5100 (29%) | 1462/5035 (29%) | 1.00 (0.94−1.06) |
| No | 66/360 (18%) | 90/375 (24%) | 0.76 (0.58−1.01) |
| **Patient SARS−CoV−2 antibody test result ($\chi^2_1$=6.6; p=0.01)** | | | |
| Positive | 616/2804 (22%) | 526/2551 (21%) | 1.07 (0.96−1.18) |
| Negative | 709/1935 (37%) | 649/1586 (41%) | 0.90 (0.82−0.97) |
| Not done | 236/754 (31%) | 386/1311 (29%) | 1.06 (0.93−1.22) |
| **All participants** | **1561/5493 (28%)** | **1561/5448 (29%)** | **0.99 (0.93−1.05)** **p=0.79** |

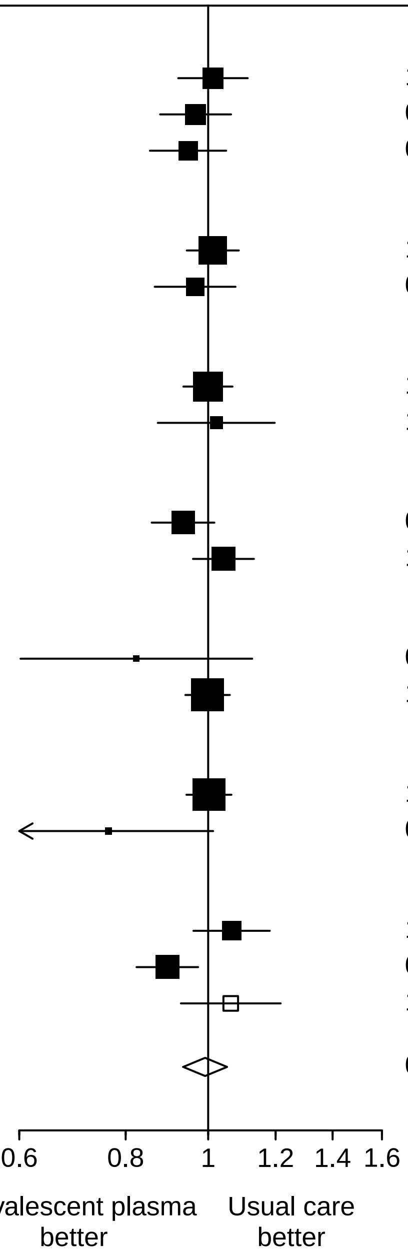

### Appendices

#### Appendix 1: RECOVERY Trial Protocol V12.1

### RANDOMISED EVALUATION OF COVID-19 THERAPY (RECOVERY)

**Background:** In early 2020, as this protocol was being developed, there were no approved treatments for COVID-19, a disease induced by the novel coronavirus SARS-CoV-2 that emerged in China in late 2019. The UK New and Emerging Respiratory Virus Threats Advisory Group (NERVTAG) advised that several possible treatments should be evaluated, including Lopinavir-Ritonavir, low-dose corticosteroids, and Hydroxychloroquine (which has now been done). A World Health Organization (WHO) expert group issued broadly similar advice. These groups also advised that other treatments will soon emerge that require evaluation.

**Eligibility and randomisation:** This protocol describes a randomised trial among patients hospitalised for COVID-19. All eligible patients are randomly allocated between several treatment arms, each to be given in addition to the usual standard of care in the participating hospital: No additional treatment *vs* colchicine *vs* corticosteroids (children only) *vs* intravenous immunoglobulin (children only). In a factorial design (in the UK alone), eligible patients are allocated simultaneously to no additional treatment *vs* convalescent plasma *vs* synthetic neutralising antibodies (REGN-COV2). Separately, all participants aged 18 years or older will be allocated to either aspirin *vs* control. The study allows a subsequent randomisation for patients with progressive COVID-19 (evidence of hyper-inflammatory state): No additional treatment *vs* tocilizumab. For patients for whom not all the trial arms are appropriate or at locations where not all are available, randomisation will be between fewer arms.

**Adaptive design:** The interim trial results will be monitored by an independent Data Monitoring Committee (DMC). The most important task for the DMC will be to assess whether the randomised comparisons in the study have provided evidence on mortality that is strong enough (with a range of uncertainty around the results that is narrow enough) to affect national and global treatment strategies. In such a circumstance, the DMC will inform the Trial Steering Committee who will make the results available to the public and amend the trial arms accordingly. Regardless, follow-up will continue for all randomised participants, including those previously assigned to trial arms that are modified or ceased. New trial arms can be added as evidence emerges that other candidate therapeutics should be evaluated.

**Outcomes:** The main outcomes will be death, discharge, need for ventilation and need for renal replacement therapy. For the main analyses, follow-up will be censored at 28 days after randomisation. Additional information on longer term outcomes may be collected through review of medical records or linkage to medical databases where available (such as those managed by NHS Digital and equivalent organisations in the devolved nations).

**Simplicity of procedures:** To facilitate collaboration, even in hospitals that suddenly become overloaded, patient enrolment (via the internet) and all other trial procedures are greatly streamlined. Informed consent is simple and data entry is minimal. Randomisation via the internet is simple and quick, at the end of which the allocated treatment is displayed on the screen and can be printed or downloaded. Follow-up information is recorded at a single timepoint and may be ascertained by contacting participants in person, by phone or electronically, or by review of medical records and databases.

**Data to be recorded:** At randomisation, information will be collected on the identity of the randomising clinician and of the patient, age, sex, major co-morbidity, pregnancy, COVID-19 onset date and severity, and any contraindications to the study treatments. The main outcomes will be death (with date and probable cause), discharge (with date), need for ventilation (with number of days recorded) and need for renal replacement therapy. Reminders will be sent if outcome data have not been recorded by 28 days after randomisation. Suspected Unexpected Serious Adverse Reactions (SUSARs) to one of the study medications (e.g., Stevens-Johnson syndrome, anaphylaxis, aplastic anaemia) will be collected and reported in an expedited fashion. Other adverse events will not be recorded but may be available through linkage to medical databases.

**Numbers to be randomised:** The larger the number randomised the more accurate the results will be, but the numbers that can be randomised will depend critically on how large the epidemic becomes. If substantial numbers are hospitalised in the participating centres then it may be possible to randomise several thousand with mild disease and a few thousand with severe disease, but realistic, appropriate sample sizes could not be estimated at the start of the trial.

**Heterogeneity between populations:** If sufficient numbers are studied, it may be possible to generate reliable evidence in certain patient groups (e.g. those with major co-morbidity or who are older). To this end, data from this study may be combined with data from other trials of treatments for COVID-19, such as those being planned by the WHO.

**Add-on studies:** Particular countries or groups of hospitals, may well want to collaborate in adding further measurements or observations, such as serial virology, serial blood gases or chemistry, serial lung imaging, or serial documentation of other aspects of disease status. While well-organised additional research studies of the natural history of the disease or of the effects of the trial treatments could well be valuable (although the lack of placebo control may bias the assessment of subjective side-effects, such as gastro-intestinal problems), they are not core requirements.

**To enquire about the trial, contact the RECOVERY Central Coordinating Office**

Nuffield Department of Population Health, Richard Doll Building, Old Road Campus, Roosevelt Drive, Oxford OX3 7LF, United Kingdom

Website: www.recoverytrial.net

**To enquire about the trial outside of the UK, contact the relevant Clinical Trial Units** (see section 10)

**To RANDOMISE a patient, visit:**

Website: www.recoverytrial.net

### 1 BACKGROUND AND RATIONALE

#### 1.1 Setting

In 2019 a novel coronavirus-disease (COVID-19) emerged in Wuhan, China. A month later the Chinese Center for Disease Control and Prevention identified a new beta-coronavirus (SARS coronavirus 2, or SARS-CoV-2) as the aetiological agent.[1] The clinical manifestations of COVID-19 range from asymptomatic infection or mild, transient symptoms to severe viral pneumonia with respiratory failure. As many patients do not progress to severe disease the overall case fatality rate per infected individual is low, but hospitals in areas with significant community transmission have experienced a major increase in the number of hospitalised pneumonia patients, and the frequency of severe disease in hospitalised patients can be as high as 30%.[2-4] The progression from prodrome (usually fever, fatigue and cough) to severe pneumonia requiring oxygen support or mechanical ventilation often takes one to two weeks after the onset of symptoms.[2] The kinetics of viral replication in the respiratory tract are not well characterized, but this relatively slow progression provides a potential time window in which antiviral therapies could influence the course of disease. In May 2020 a new COVID-associated inflammatory syndrome in children was identified, Paediatric Inflammatory Multisystem Syndrome - Temporally associated with SARS-CoV-2 (PIMS-TS).[5] A rapid NHS England-led consensus process identified the need to evaluate corticosteroids and intravenous immunoglobulin (IVIg) as initial therapies in PIMS-TS, and confirmed tocilizumab as one of the biological anti-inflammatory agents to be evaluated as a second line therapy.

#### 1.2 Treatment Options

##### 1.2.1 Main randomisation

This protocol allows reliable assessment of the effects of multiple different treatments (including re-purposed and novel drugs) on major outcomes in COVID-19. All patients will receive usual care for the participating hospital.

**Randomisation part A:** Eligible patients may be randomly allocated between the following treatment arms:

- **No additional treatment**
- **Colchicine (men ≥18 years old and women ≥55 years old only)**
- **Corticosteroids (children ≤44 weeks gestational age, or >44 weeks gestational age with PIMS-TS only)**
- **Intravenous immunoglobulin (children >44 weeks gestational age with PIMS-TS only)**

**Randomisation part B [UK only]:** Simultaneously, eligible patients will be randomly allocated between the following treatment arms:

- **No additional treatment**
- **Convalescent plasma**
- **Synthetic neutralising antibodies (REGN-COV2) (adults and children ≥12 years old only)**

**Randomisation part C (adults ≥18 years old only):** Simultaneously, eligible patients will be randomly allocated between the following treatment arms:

- **No additional treatment**
- **Aspirin**

##### 1.2.2 Second randomisation for patients with progressive COVID-19

Severe COVID-19 is associated with release of pro-inflammatory cytokines, such as IL-1, IL-6 and TNFα, and other markers of systemic inflammation including ferritin and C-reactive protein.[6-8] There is a possibility that this response may cause or exacerbate lung injury, leading to life-threatening disease.

Participants with progressive COVID-19 (as evidenced by hypoxia and an inflammatory state) may undergo an optional second randomisation between the following treatment arms:

- **No additional treatment**
- **Tocilizumab**

##### 1.2.3 Modifications to the number of treatment arms

Other arms can be added to the first or second randomisation if evidence emerges that there are suitable candidate therapeutics. Conversely, in some patient populations, not all trial arms are appropriate (e.g. due to contraindications based on co-morbid conditions or concomitant medication); in some hospitals or countries, not all treatment arms will be available (e.g. due to manufacturing and supply shortages); and at some times, not all treatment arms will be active (e.g. due to lack of relevant approvals and contractual agreements). The Trial Steering Committee may elect to pause one or more of the arms in order to increase trial efficiency during a fluctuating epidemic. In any of these situations, randomisation will be between fewer arms. Depending on the availability and suitability of treatments, it may be allowed for participants to be randomised in only one or two parts (A, B, or C) of the main randomisation.

##### 1.3 Design Considerations

The RECOVERY Protocol describes an overarching trial design to provide reliable evidence on the efficacy of candidate therapies for suspected or confirmed COVID-19 infection in hospitalised patients receiving usual standard of care.

In early 2020, when the trial first started, there were no known treatments for COVID-19. The anticipated scale of the epidemic is such that hospitals, and particularly intensive care facilities, may be massively overstretched at some points in time, with around 10% requiring hospitalisation. In this situation, even treatments with only a moderate impact on survival or on hospital resources could be worthwhile. Therefore, the focus of RECOVERY is the impact of candidate treatments on mortality and on the need for hospitalisation or ventilation.

Critically, the trial is designed to minimise the burden on front-line hospital staff working within an overstretched care system during a major epidemic. Eligibility criteria are therefore simple and trial processes (including paperwork) are minimised.

The protocol is deliberately flexible so that it is suitable for a wide range of settings, allowing:

- a broad range of patients to be enrolled in large numbers;
- randomisation between only those treatment arms that are *both* available at the hospital *and* not believed by the enrolling doctor to be contraindicated (e.g. by particular co-morbid conditions or concomitant medications);
- treatment arms to be added or removed according to the emerging evidence; and
- additional substudies may be added to provide more detailed information on side effects or sub-categorisation of patient types but these are not the primary objective and are not required for participation.

In a cohort of 191 hospitalised COVID-19 patients with a completed outcome, the median time from illness onset to discharge was 22·0 days (IQR 18·0–25·0) and the median time to death was 18·5 days (15·0–22·0). Thirty-two patients (17%) required invasive mechanical ventilation and the median time from onset to mechanical ventilation was 14.5 days. Therefore, early endpoint assessment, such as 28 days after randomisation, is likely to provide largely complete outcome data and will permit early assessment of treatment efficacy and safety.[9]

##### 1.4 Potential for effective treatments to become available

In early 2020, when the trial first started, there were no known treatments for COVID-19. However, over time, effective treatments may become available, typically as the result of reliable information from randomised trials (including from this study). For example, in June 2020, results from the RECOVERY trial showed that dexamethasone reduces the mortality in COVID-19 patients requiring mechanical ventilation or oxygen. In response, many clinical guidelines now recommend the use of dexamethasone as standard of care for these types of patients.

The RECOVERY trial randomises eligible participant to usual standard of care for the local hospital alone vs usual standard of care plus one or more additional study treatments. Over time, it is expected that usual standard of care alone will evolve. Thus randomisation will always be relevant to the current clinical situation and the incremental effects of the study treatments will be appropriately assessed.

### 2 DESIGN AND PROCEDURES

#### 2.1 Eligibility

Patients are eligible for the study if all of the following are true:

**(i) Hospitalised**

**(ii) SARS-CoV-2 infection associated disease (clinically suspected or laboratory confirmed)**

In general, SARS-CoV-2 infection should be suspected when a patient presents with:

a) typical symptoms (e.g. influenza-like illness with fever and muscle pain, or respiratory illness with cough and shortness of breath); and
b) compatible chest X-ray findings (consolidation or ground-glass shadowing); and
c) alternative causes have been considered unlikely or excluded (e.g. heart failure, influenza).

However, the diagnosis remains a clinical one based on the opinion of the managing doctor.

A small number of children (aged <18 years) present with atypical features, including a hyperinflammatory state and evidence of single or multi-organ dysfunction (called Paediatric Multisystem Inflammatory Syndrome temporally associated with COVID-19 [PIMS-TS]). Some do not have significant lung involvement.[a]

**(iii) No medical history that might, in the opinion of the attending clinician, put the patient at significant risk if he/she were to participate in the trial**

In addition, if the attending clinician believes that there is a specific contra-indication to one of the active drug treatment arms (see Appendix 2; section 8.2 and Appendix 3; section 8.3 for children) or that the patient should definitely be receiving one of the active drug treatment arms then that arm will not be available for randomisation for that patient. For patients who lack capacity, an advanced directive or behaviour that clearly indicates that they would not wish to participate in the trial would be considered sufficient reason to exclude them from the trial.

In some locations, children (aged <18 years) will not be recruited, to comply with local and national regulatory approvals (see Section 8.3).

---

[a] https://www.rcpch.ac.uk/sites/default/files/2020-05/COVID-19-Paediatric-multisystem-%20inflammatory%20syndrome-20200501.pdf

#### 2.2 Consent

Informed consent should be obtained from each patient 16 years and over before enrolment into the study. However, if the patient lacks capacity to give consent due to the severity of their medical condition (e.g. acute respiratory failure or need for immediate ventilation) or prior disease, then consent may be obtained from a relative acting as the patient's legally designated representative or – if a suitable relative is not available after reasonable efforts to locate one – an independent doctor. Further consent will then be sought with the patient if they recover sufficiently. For children aged <16 years old consent will be sought from their parents or legal guardian. Where possible, children aged between 10-15 years old will also be asked for assent. Children aged ≥16 years old will asked for consent as for adults. Witnessed consent may be obtained over the telephone or web video link if hospital visiting rules or parental infection mean a parent/guardian cannot be physically present.

Due to the poor outcomes in COVID-19 patients who require ventilation (>90% mortality in one cohort[9]), patients who lack capacity to consent due to severe disease (e.g. needs ventilation), and for whom a relative to act as the legally designated representative is not available, randomisation and consequent treatment will proceed with consent provided by a treating clinician (independent of the clinician seeking to enrol the patient) who will act as the legally designated representative (if allowed by local regulations). Consent will then be obtained from the patient's personal legally designated representative (or directly from the patient if they recover promptly) at the earliest opportunity.

#### 2.3 Baseline information

The following information will be recorded on the web-based form by the attending clinician or delegate:

- Patient details (e.g. name or initials [depending on privacy requirements], NHS/CHI number [UK only] or medical records number, date of birth, sex)
- Clinician details (e.g. name)
- COVID-19 symptom onset date
- COVID-19 severity as assessed by need for supplemental oxygen, non-invasive ventilation or invasive mechanical ventilation/extracorporeal membrane oxygenation (ECMO)
- Oxygen saturations on air (if available)
- Latest routine measurement of creatinine, C-reactive protein, and D-dimer (if available)
- SARS-CoV-2 PCR test result (if available)
- Major co-morbidity (e.g. heart disease, diabetes, chronic lung disease) and pregnancy
- Use of relevant medications (corticosteroids, remdesivir, antiplatelet and anticoagulant therapy)
- Date of hospitalisation
- Contraindication to the study treatment regimens (in the opinion of the attending clinician)
- Willingness to receive a blood product [UK only]
- Name of person completing the form

The person completing the form will then be asked to confirm that they wish to randomise the patient and will then be required to enter their name and e-mail address.

##### 2.4 Main randomisation

In addition to receiving usual care, eligible patients will be allocated using a central web-based randomisation service (without stratification or minimisation). From version 6.0 of the protocol, a factorial design will be used such that eligible patients may be randomised to one of the treatment arms in Randomisation A and, simultaneously, to one of the treatment arms in Randomisation B. From version 10.0 of the protocol, a further factorial randomisation was added (Main Randomisation part C). From version 11.3 of the protocol, children may be recruited into the trial even if there are no main randomisation treatments which are both available and suitable provided they meet the criteria for inclusion in the second randomisation, per section 2.5. They will not be allocated to a main randomisation group, but will be potentially eligible for the second randomisation between tocilizumab and control.

###### 2.4.1 Main randomisation part A:

Eligible patients may be randomised to one of the arms listed below. The doses in this section are for adults. Please see Appendix 3 for paediatric dosing. Study treatments do not need to be continued after discharge from hospital.

- **No additional treatment**

- **Colchine 1 mg after randomisation followed by 500mcg 12 hours later and then 500 mcg twice daily** by mouth or nasogastric tube for 10 days in total.[b] (Men ≥18 years old and women ≥55 years old only.)

- **Corticosteroid (in children ≤44 weeks gestational age, or >44 weeks gestational age with PIMS-TS only)**: see Appendix 3.

- **Intravenous immunoglobulin (in children >44 weeks gestational age with PIMS-TS only)**: see Appendices 2 and 3 for dose, contraindications and monitoring information.

For randomisation part A, the randomisation program will allocate patients in a ratio of 1:1 between the no additional treatment arm and each of the other arms available. If one or more of the active drug treatments is not available at the hospital or is believed, by the attending clinician, to be contraindicated (or definitely indicated) for the specific patient, then this fact will be recorded via the web-based form prior to randomisation; random allocation will then be between the remaining arms. If no treatments are both available and suitable, then it may be possible to only be randomised in part B (UK only) and/or part C.

###### 2.4.2 Main randomisation part B [UK only]:

Eligible patients may be randomised to one of the arms listed below. The doses in this section are for adults. Please see Appendix 3 for paediatric dosing. **Participants in this randomisation should have a serum sample sent to their transfusion laboratory prior to randomisation in which presence of antibodies against SARS-CoV-2 may be tested.**

- **No additional treatment**

---

[b] Treatment should be discontinued at 10 days or on discharge from hospital if sooner

- **Convalescent plasma** Single unit of ABO compatible convalescent plasma (275mls +/- 75 mls) intravenous per day on study days 1 (as soon as possible after randomisation) and 2 (with a minimum of 12 hour interval between 1st and 2nd units). ABO identical plasma is preferred if available. The second transfusion should not be given if patient has a suspected serious adverse reaction during or after the first transfusion.

- **Synthetic neutralising antibodies (REGN-COV2; adults and children aged ≥12 years[c] only).** A single dose of REGN10933 + REGN10987 8 g (4 g of each monoclonal antibody) in 250ml 0.9% saline infused intravenously over 60 minutes +/- 15 minutes as soon as possible after randomisation

For randomisation part B, the randomisation program will allocate patients in a ratio of 1:1:1 between each of the arms. If the active treatment is not available at the hospital, the patient does not consent to receive convalescent plasma, or is believed, by the attending clinician, to be contraindicated for the specific patient, then this fact will be recorded via the web-based form and the patient will be excluded from the relevant arm in Randomisation part B.

##### 2.4.3 Main randomisation part C [adults aged ≥18 years only]:

Eligible patients may be randomised to one of the arms listed below.

- **No additional treatment**

- **Aspirin** 150 mg by mouth (or nasogastric tube) or per rectum once daily until discharge.[d]

Note: The allocation in this randomisation should not influence the use of standard thromboprophylaxis care.

The randomisation program will allocate patients in a ratio of 1:1 between the arms being evaluated in part C of the main randomisation.

#### 2.5 Second randomisation for patients with progressive COVID-19

Patients enrolled in the RECOVERY trial and with clinical evidence of a hyper-inflammatory state may be considered for a second randomisation if they meet the following criteria:

(i) Recruited into the RECOVERY trial no more than 21 days ago[e]
(ii) Clinical evidence of progressive COVID-19:
   a. oxygen saturation <92% on room air or requiring oxygen (or in children (age <18 years), significant systemic disease with persistent pyrexia, with or without evidence of respiratory involvement)[f]; and

[c] Older children who weigh <40kg will also not be eligible for this treatment.
[d] In countries where 150mg aspirin is not available, up to 162mg may be used instead
[e] Children recruited into RECOVERY for whom no main randomisation treatment are both available and suitable (see section 2.4) should undergo this second randomisation as soon as possible after recruitment.
[f] A small number of children (age <18 years) present with atypical features, including a hyperinflammatory state and evidence of single or multi-organ dysfunction. Some do not have significant lung involvement.

b. C-reactive protein ≥75 mg/L

(iii) No medical history that might, in the opinion of the attending clinician, put the patient at significant risk if he/she were to participate in this aspect of the RECOVERY trial. (Note: Pregnancy and breastfeeding are not specific exclusion criteria.)

Note: Participants may undergo this second randomisation at any point after being first randomised, provided they meet the above criteria, and thus may receive up to four study treatments (one each from Main randomisation parts A, B and C, plus one from the second randomisation). For some participants the second randomisation may be immediately after the first but for others it may occur a few hours or days later, if and when they deteriorate.

The following information will be recorded (on the web-based form) by the attending clinician or delegate:

- Patient details (e.g. name or initials, NHS/CHI number [UK only] or medical records number, date of birth, sex)
- Clinician details (e.g. name)
- COVID-19 severity as assessed by need for supplemental oxygen or ventilation/ECMO
- Markers of progressive COVID-19 (including oxygen saturation, C-reactive protein)
- Contraindication to the study drug treatments (in the opinion of the attending clinician)
- Name of person completing the form

The person completing the form will then be asked to confirm that they wish to randomise the patient and will then be required to enter their own name and e-mail address.

Eligible participants may be randomised between the following treatment arms:

- **No additional treatment**

- **Tocilizumab** by intravenous infusion with the dose determined by body weight:

| Weight* | Dose |
|---|---|
| >40 and ≤65 kg | 400 mg |
| >65 and ≤90 kg | 600 mg |
| >90 kg | 800 mg |

* for lower weights, dosing should be 8 mg/kg (see Appendix 3 for paediatric dosing)
(Note: body weight may be estimated if it is impractical to weigh the patient)

Tocilizumab should be given as a single intravenous infusion over 60 minutes in 100ml sodium chloride 0.9%. A second dose may be given ≥12 and <24 hours later if, in the opinion of the attending clinician, the patient's condition has not improved.

The randomisation program will allocate patients in a ratio of 1:1 between the arms being evaluated in the second randomisation. Participants should receive standard management

---

(see: https://www.rcpch.ac.uk/sites/default/files/2020-05/COVID-19-Paediatric-multisystem-%20inflammatory%20syndrome-20200501.pdf) (including blood tests such as liver function tests and full blood count) according to their clinical need.

#### 2.6 Administration of allocated treatment

The details of the allocated study treatments will be displayed on the screen and can be printed or downloaded. The hospital clinicians are responsible for prescription and administration of the allocated treatments. The patient's own doctors are free to modify or stop study treatments if they feel it is in the best interests of the patient without the need for the patient to withdraw from the study (see section 2.9). This study is being conducted within hospitals. Therefore use of medication will be subject to standard medication reviews (typically within 48 hours of enrolment) which will guide modifications to both the study treatment and use of concomitant medication (e.g. in the case of potential drug interactions).

Note: [UK only] NHS guidelines require patients to have **two** separate blood samples taken for Group and Screen prior to administration of blood products. Each sample is approximately 5 mL and both need to be taken at any time between admission to hospital and receipt of the first plasma transfusion (as the laboratory will not issue plasma without both samples), although if a valid historical sample exists this can be used for one of the samples. The participant's blood group is identified to ensure that blood group-compatible plasma is given and this information would be available to the participant if they wish. Such tests may be required as part of the routine care of the participant if the managing team wish to consider using blood products and samples will be stored, retained and destroyed as per trust standard procedures and protocols. The extra serum sample collected for measurement of coronavirus and antibodies against it will be prepared in the local transfusion laboratory (including removing any identifiers and labelling with the participant's study ID) and sent to a central laboratory for analysis. Once testing is complete these samples will be destroyed.

#### 2.7 Collecting follow-up information

The following information will be ascertained at the time of death or discharge or at 28 days after first randomisation (whichever is sooner):

- Vital status (alive / dead, with date and presumed cause of death, if appropriate)
- Hospitalisation status (inpatient / discharged, with date of discharge, if appropriate)
- SARS-CoV-2 test result
- Use of ventilation (with days of use and type, if appropriate)
- Use of renal dialysis or haemofiltration
- Documented new major cardiac arrhythmia (including atrial and ventricular arrhythmias)
- Major bleeding (defined as intracranial bleeding or bleeding requiring transfusion, endoscopy, surgery, or vasoactive drugs)
- Thrombotic event, defined as either (i) acute pulmonary embolism; (ii) deep vein thrombosis; (iii) ischaemic stroke; (iv) myocardial infarction; or (v) systemic arterial embolism.
- Use of any medications included in the RECOVERY trial protocol (including drugs in the same class) or other purported COVID-19 treatments (e.g. remdesivir, favipiravir)
- Participation in other randomised trials of interventions (vaccines or treatments) for COVID-19.

- Additional information including results of routine tests (including full blood count, coagulation and inflammatory markers, cardiac biomarkers, electro- and echo-cardiograms) and other treatments given will be collected for children in the UK. This information will be obtained and entered into the web-based IT system by a member of the hospital clinical or research staff. At some locations, electrocardiograms done as part of routine care of adult participants will also be collected.

Follow-up information is to be collected on all study participants, irrespective of whether or not they complete the scheduled course of allocated study treatment. Study staff will seek follow-up information through various means including medical staff, reviewing information from medical notes, routine healthcare systems, and registries.

For all randomised participants, vital status (alive / dead, with date and presumed cause of death, if appropriate) is to be ascertained at 28 days after first randomisation. This may be achieved through linkage to routine death registration data (e.g. in the UK) or through direct contact with the participant, their relatives, or medical staff and completion of an additional follow-up form.

##### 2.7.1 Additional assessment of safety of antibody-based therapy [UK only]

For at least the first 200 participants in each comparison in Main Randomisation part B (no additional treatment vs. convalescent plasma and no additional treatment vs. synthetic neutralising antibody), the following information will be collected on the following events occurring within the first 72 hours after randomisation:

- Sudden worsening in respiratory status
- Severe allergic reaction or other infusion reaction
- Temperature >39ºC or ≥2ºC rise above baseline
- Sudden hypotension, defined as either (i) sudden drop in systolic blood pressure of ≥30 mmHg with systolic blood pressure ≤80 mmHg; or (ii) requiring urgent medical attention
- Clinical haemolysis, defined as fall in haemoglobin plus one or more of the following: rise in lactate dehydrogenase (LDH), rise in bilirubin, positive direct antiglobulin test (DAT), or positive crossmatch
- Thrombotic event, defined as either (i) acute pulmonary embolism; or (ii) deep-vein thrombosis; or (iii) ischaemic stroke; or (iv) myocardial infarction; or (v) systemic arterial embolism.

The Data Monitoring Committee will review unblinded information on these outcomes and advise if, in their view, the collection of such information should be extended to more participants.

In addition, Serious Hazards Of Transfusion (SHOT) reporting will be conducted for all patients receiving convalescent plasma for the full duration of the study (see section 4.1).

#### 2.8 Duration of follow-up

All randomised participants are to be followed up until death, discharge from hospital or 28 days after randomisation (whichever is sooner). It is recognised that in the setting of this trial, there may be some variability in exactly how many days after randomisation, information on disease status is collected. This is acceptable and will be taken account of in the analyses and interpretation of results, the principle being that some information about post-randomisation disease status is better than none.

In the UK, longer term (up to 10 years) follow-up will be sought through linkage to electronic healthcare records and medical databases including those held by NHS Digital, Public Health England and equivalent bodies, and to relevant research databases (e.g. UK Biobank, Genomics England).

##### 2.9 Withdrawal of consent

A decision by a participant (or their parent/guardian) that they no longer wish to continue receiving study treatment should **not** be considered to be a withdrawal of consent for follow-up. However, participants (or their parent/guardian) are free to withdraw consent for some or all aspects of the study at any time if they wish to do so. In accordance with regulatory guidance, de-identified data that have already been collected and incorporated in the study database will continue to be used (and any identifiable data will be destroyed).

For participants who lack capacity, if their legal representative withdraws consent for treatment or methods of follow-up then these activities would cease.

#### 3 STATISTICAL ANALYSIS

All analyses for reports, presentations and publications will be prepared by the coordinating centre at the Nuffield Department of Population Health, University of Oxford. A more detailed statistical analysis plan will be developed by the investigators and published on the study website whilst still blind to any analyses of aggregated data on study outcomes by treatment allocation.

##### 3.1 Outcomes

For each pairwise comparison with the 'no additional treatment' arm, the **primary objective** is to provide reliable estimates of the effect of study treatments on all-cause mortality at 28 days after randomisation (with subsidiary analyses of cause of death and of death at various timepoints following discharge).

The **secondary objectives** are to assess the effects of study treatments on duration of hospital stay; and, among patients not on invasive mechanical ventilation at baseline, the composite endpoint of death or need for invasive mechanical ventilation or ECMO.

Other objectives include the assessment of the effects of study treatments on the need for any ventilation (and duration of invasive mechanical ventilation), renal replacement therapy and thrombotic events. Safety outcomes include bleeding, new major cardiac arrhythmias and (assessed at 72 hours after randomization among participants in main randomization part B only) sudden worsening in respiratory status, severe allergic reaction, significant fever, sudden hypotension and clinical haemolysis.

Study outcomes will be assessed based on data recorded up to 28 days and up to 6 months after randomisation.

Where available, data from routine healthcare records (including linkage to medical databases held by organisations such as NHS Digital in the UK) and from relevant research studies (such as UK Biobank, Genomics England, ISARIC-4C and PHOSP-COVID) will allow subsidiary analyses of the effect of the study treatments on particular non-fatal events (e.g. ascertained through linkage to Hospital Episode Statistics), the influence of pre-existing major co-morbidity (e.g. diabetes, heart disease, lung disease, hepatic insufficiency, severe depression, severe kidney impairment, immunosuppression), and longer-term outcomes as well as in particular sub-categories of patient (e.g. by genotype, pregnancy).

#### 3.2 Methods of analysis

For all outcomes, comparisons will be made between all participants randomised to the different treatment arms, irrespective of whether they received their allocated treatment ("intention-to-treat" analyses).

For time-to-event analyses, each treatment group will be compared with the no additional treatment group using the log-rank test. Kaplan-Meier estimates for the time to event will also be plotted (with associated log-rank p-values). The log-rank 'observed minus expected' statistic (and its variance) will also be used to estimate the average event rate ratio (and its confidence interval) for those allocated to each treatment group versus the no additional treatment group. For binary outcomes where the timing is unknown, the risk ratio and absolute risk difference will be calculated with confidence intervals and p-value reported. For the primary outcome (death within 28 days of randomisation), discharge alive before 28 days will assume safety from the event (unless there is additional data confirming otherwise).

Pairwise comparisons within each randomisation will be made between each treatment arm and the no additional treatment arm (reference group) in that particular randomisation (main randomisation part A, B or C, and second randomisation). However, since not all treatments may be available or suitable for all patients, those in the no additional treatment arm will only be included in a given comparison if, at the point of their randomisation, they *could* alternatively have been randomised to the active treatment of interest. Allowance for multiple treatment comparisons due to the multi-arm design will be made. All p-values will be 2-sided.

Pre-specified subgroup analysis (e.g., level of respiratory support, time since onset of symptoms; sex; age group; ethnicity; use of corticosteroids) will be conducted for the primary outcome using the statistical test for interaction (or test for trend where appropriate). Sensitivity analyses will be conducted among those patients with laboratory confirmed SARS-CoV-2.

Further details will be fully described in the Statistical Analysis Plan.

### 4 DATA AND SAFETY MONITORING

#### 4.1 Recording Suspected Serious Adverse Reactions

The focus is on those events that, based on a single case, are highly likely to be related to the study medication. Examples include anaphylaxis, Stevens Johnson Syndrome, or bone marrow failure, where there is no other plausible explanation.

Any Serious Adverse Event[g] that is believed with a reasonable probability to be due to one of the study treatments will be considered a Suspected Serious Adverse Reaction (SSAR). In making this assessment, there should be consideration of the probability of an alternative cause (for example, COVID-19 itself or some other condition preceding randomisation), the timing of the event with respect to study treatment, the response to withdrawal of the study treatment, and (where appropriate) the response to subsequent re-challenge.

All SSARs should be reported by telephone to the Central Coordinating Office and recorded on the study IT system immediately.

[UK only] Suspected serious transfusion reactions in patients who receive convalescent plasma should additionally be reported to Serious Hazards of Transfusions (SHOT) and through the MHRA Serious Adverse Blood Reactions and Events (SABRE) system.[h]

#### 4.2 Central assessment and onward reporting of SUSARs

Clinicians at the Central Coordinating Office are responsible for expedited review of reports of SSARs received. Additional information (including the reason for considering it both serious and related, and relevant medical and medication history) will be sought.

The focus of Suspected Unexpected Serious Adverse Reaction (SUSAR) reporting will be on those events that, based on a single case, are highly likely to be related to the study medication. To this end, anticipated events that are either efficacy endpoints, consequences of the underlying disease, or common in the study population will be exempted from expedited reporting. Thus the following events will be exempted from expedited reporting:

(i) Events which are the consequence of COVID-19; and
(ii) Common events which are the consequence of conditions preceding randomisation.

Any SSARs that are not exempt will be reviewed by a Central Coordinating Office clinician and an assessment made of whether the event is “expected” or not (assessed against the relevant Summary of Product Characteristics or Investigator Brochure). Any SSARs that are not expected would be considered a Suspected Unexpected Serious Adverse Reaction (SUSAR).

---

[g] Serious Adverse Events are defined as those adverse events that result in death; are life-threatening; require in-patient hospitalisation or prolongation of existing hospitalisation; result in persistent or significant disability or incapacity; result in congenital anomaly or birth defect; or are important medical events in the opinion of the responsible investigator (that is, not life-threatening or resulting in hospitalisation, but may jeopardise the participant or require intervention to prevent one or other of the outcomes listed above).
[h] https://www.shotuk.org/reporting/

All confirmed SUSARs will be reported to the Chair of the DMC and to relevant regulatory authorities, ethics committees, and investigators in an expedited manner in accordance with regulatory requirements.

#### 4.3 Recording other Adverse Events

In addition to recording Suspected Serious Adverse Reactions (see section 4.1), information will be collected on all deaths and efforts will be made to ascertain the underlying cause. Other serious or non-serious adverse events will not be recorded unless specified in section 2.7. It is anticipated that for some substudies, more detailed information on adverse events (e.g. through linkage to medical databases) or on other effects of the treatment (e.g. laboratory or radiological features) will be recorded and analysed but this is not a requirement of the core protocol.

#### 4.4 Role of the Data Monitoring Committee (DMC)

During the study, interim analyses of all study data will be supplied in strict confidence to the independent DMC. The DMC will request such analyses at a frequency relevant to the emerging data from this and other studies.

The DMC will independently evaluate these analyses and any other information considered relevant. The DMC will determine if, in their view, the randomised comparisons in the study have provided evidence on mortality that is strong enough (with a range of uncertainty around the results that is narrow enough) to affect national and global treatment strategies. In such a circumstance, the DMC will inform the Trial Steering Committee who will make the results available to the public and amend the trial arms accordingly. Unless this happens, the Trial Steering Committee, Chief Investigator, study staff, investigators, study participants, funders and other partners will remain blind to the interim results until 28 days after the last patient has been randomised for a particular intervention arm (at which point analyses may be conducted comparing that arm with the no additional treatment arm).

The DMC will review the safety and efficacy analyses among children (age <18 years) both separately and combined with the adult data. As described in section 2.7.1, the DMC will advise if collection of information relating to the safety of convalescent plasma should be extended beyond the first 200 patients enrolled to each comparison in Main Randomisation part B.

#### 4.5 Blinding

This is an open-label study. However, while the study is in progress, access to tabular results of study outcomes by allocated treatment allocation will not be available to the research team, patients, or members of the Trial Steering Committee (unless the DMC advises otherwise).

### 5 QUALITY MANAGEMENT

#### 5.1 Quality By Design Principles

In accordance with the principles of Good Clinical Practice and the recommendations and guidelines issued by regulatory agencies, the design, conduct and analysis of this trial is focussed on issues that might have a material impact on the wellbeing and safety of study participants (hospitalised patients with suspected or confirmed SARS-CoV-2 infection) and the reliability of the results that would inform the care for future patients.

The critical factors that influence the ability to deliver these quality objectives are:

- to minimise the burden on busy clinicians working in an overstretched hospital during a major epidemic
- to ensure that suitable patients have access to the trial medication without impacting or delaying other aspects of their emergency care
- to provide information on the study to patients and clinicians in a timely and readily digestible fashion but without impacting adversely on other aspects of the trial or the patient's care
- to allow individual clinicians to use their judgement about whether any of the treatment arms are not suitable for the patient
- to collect comprehensive information on the mortality and disease status

In assessing any risks to patient safety and well-being, a key principle is that of proportionality. Risks associated with participation in the trial must be considered in the context of usual care. At present, there are no proven treatments for COVID-19, basic hospital care (staffing, beds, ventilatory support) may well be overstretched, and mortality for hospitalised patients may be around 10% (or more in those who are older or have significant co-morbidity).

#### 5.2 Training and monitoring

The focus will be on those factors that are critical to quality (i.e. the safety of the participants and the reliability of the trial results). Remedial actions would focus on issues with the potential to have a substantial impact on the safety of the study participants or the reliability of the results.

The study will be conducted in accordance with the principles of International Conference on Harmonisation Guidelines for Good Clinical Research Practice (ICH-GCP) and relevant local, national and international regulations. Any serious breach of GCP in the conduct of the clinical trial will be handled in accordance with regulatory requirements. Prior to initiation of the study at each Local Clinical Centre (LCC), the Central Coordinating Office (CCO) or relevant Regional Coordinating Centre (RCC) will confirm that the LCC has adequate facilities and resources to carry out the study. LCC lead investigators and study staff will be provided with training materials.

In the context of this epidemic, visits to hospital sites is generally not appropriate as they could increase the risks of spreading infection, and in the context of this trial they generally would not influence the reliability of the trial results or the well-being of the participants. In exceptional circumstances, the CCO or RCC may arrange monitoring visits to LCCs as considered appropriate based on perceived training needs and the results of central statistical monitoring of study data.[10,11] The purpose of such visits will be to ensure that the study is being conducted in accordance with the protocol, to help LCC staff to resolve any local problems, and to provide extra training focussed on specific needs. No routine source data verification will take place.

In the UK, training of laboratory and transfusion staff and initiation of convalescent plasma delivery will be performed by NHS Blood and Transplant Clinical Trials Unit.

##### 5.3 Data management

LCC clinic staff will use the bespoke study web-based applications for study management and to record participant data (including case report forms) in accordance with the protocol. Data will be held in central databases located at the CCO or on secure cloud servers. In some circumstances (e.g. where there is difficulty accessing the internet or necessary IT equipment), paper case report forms may be required with subsequent data entry by either LCC or CCO staff. Although data entry should be mindful of the desire to maintain integrity and audit trails, in the circumstances of this epidemic, the priority is on the timely entry of data that is sufficient to support reliable analysis and interpretation about treatment effects. CCO staff will be responsible for provision of the relevant web-based applications and for generation of data extracts for analyses.

All data access will be controlled by unique usernames and passwords, and any changes to data will require the user to enter their username and password as an electronic signature in accordance with regulatory requirements.[12] Staff will have access restricted to the functionality and data that are appropriate for their role in the study.

##### 5.4 Source documents and archiving

Source documents for the study constitute the records held in the study main database. These will be retained for at least 25 years from the completion of the study. Identifiable data will be retained only for so long as it is required to maintain linkage with routine data sources (see section 2.8), with the exception of children for whom such data must be stored until they reach 21 years old (due to the statute of limitations). The sponsor and regulatory agencies will have the right to conduct confidential audits of such records in the CCO and LCCs (but should mindful of the workload facing participating hospitals and the infection control requirements during this epidemic).

#### 6 OPERATIONAL AND ADMINISTRATIVE DETAILS

##### 6.1 Sponsor and coordination

The University of Oxford will act as the trial Sponsor. The trial will be coordinated by a Central Coordinating Office (CCO) within the Nuffield Department of Population Health staffed by members of the two registered clinical trials units – the Clinical Trial Service Unit and the National Perinatal Epidemiology Unit Clinical Trials Unit. The CCO will oversee Regional Coordinating Centres which will assist with selection of Local Clinical Centres (LCCs) within their region and for the administrative support and monitoring of those LCCs. The data will be collected, analysed and published independently of the source of funding.

#### 6.2 Funding

This study is supported by a grant to the University of Oxford from UK Research and Innovation/National Institute for Health Research (NIHR) and by core funding provided by NIHR Oxford Biomedical Research Centre, the Wellcome Trust, the Bill and Melinda Gates Foundation, Department for International Development, Health Data Research UK, NIHR Health Protection Unit in Emerging and Zoonotic Infections and the Medical Research Council Population Health Research Unit, and NIHR Clinical Trials Unit Support Funding.

#### 6.3 Indemnity

The University has a specialist insurance policy in place which would operate in the event of any participant suffering harm as a result of their involvement in the research (Newline Underwriting Management Ltd, at Lloyd's of London). In the UK, NHS indemnity operates in respect of the clinical treatment that is provided.

#### 6.4 Local Clinical Centres

The study will be conducted at multiple hospitals (LCCs) within each region. At each LCC, a lead investigator will be responsible for trial activities but much of the work will be carried out by medical staff attending patients with COVID-19 within the hospital and by hospital research nurses, medical students and other staff with appropriate education, training, and experience. Where LCCs plan to recruit children the principal investigator will co-opt support from a local paediatrician and/or neonatologists to oversee the management of children and infants in the trial.

#### 6.5 Supply of study treatments

For licensed treatments (e.g. lopinavir-ritonavir, corticosteroids, tocilizumab) all aspects of treatment supply, storage, and management will be in accordance with standard local policy and practice for prescription medications. Treatment issue to randomised participants will be by prescription. Such study treatments will not be labelled other than as required for routine clinical use. They will be stored alongside other routine medications with no additional monitoring. No accountability records will be kept beyond those used for routine prescriptions.

For unlicensed treatments, manufacture, packaging, labelling and delivery will be the responsibility of the pharmaceutical company and, in the UK, the Department of Health and Social Care. Each LCC will maintain an accountability log and will be responsible for the storage and issue of study treatment. If treatments require storage at a specific temperature, LCCs can use existing temperature-controlled facilities and associated monitoring. Treatment issue to randomised participants will be in accordance with local practice (and may be in line with the processes required for routine prescriptions or compassionate use).

For convalescent plasma in the UK, manufacture, packaging, and delivery will be the responsibility of the relevant UK Blood Service (NHS Blood and Transplant for England, Welsh Blood Service for Wales, Scottish National Blood Transfusion Service for Scotland, and the Northern Ireland Blood Transfusion Service for Northern Ireland). Convalescent plasma will be labelled in accordance with regulatory requirements and the unit will be issued to the ward for a named patient in a bag marked for clinical trial use only.

Treatment will be issued to randomised participants by prescription.

#### 6.6 End of trial

The end of the scheduled treatment phase is defined as the date of the last follow-up visit of the last participant. In the UK, it is intended to extend follow-up for a year or more beyond the final study visit through linkage to routine medical records and central medical databases. The end of the study is the date of the final data extraction from NHS Digital (anticipated to be 10 years after the last patient is enrolled).

#### 6.7 Publications and reports

The Trial Steering Committee will be responsible for drafting the main reports from the study and for review of any other reports. In general, papers initiated by the Trial Steering Committee (including the primary manuscript) will be written in the name of the RECOVERY Collaborative Group, with individual investigators named personally at the end of the report (or, to comply with journal requirements, in web-based material posted with the report).

The Trial Steering Committee will also establish a process by which proposals for additional publications (including from independent external researchers) are considered by the Trial Steering Committee. The Trial Steering Committee will facilitate the use of the study data and approval will not be unreasonably withheld. However, the Trial Steering Committee will need to be satisfied that any proposed publication is of high quality, honours the commitments made to the study participants in the consent documentation and ethical approvals, and is compliant with relevant legal and regulatory requirements (e.g. relating to data protection and privacy). The Trial Steering Committee will have the right to review and comment on any draft manuscripts prior to publication.

#### 6.8 Substudies

Proposals for substudies must be approved by the Trial Steering Committee and by the relevant ethics committee and competent authorities (where required) as a substantial amendment or separate study before they begin. In considering such proposals, the Trial Steering Committee will need to be satisfied that the proposed substudy is worthwhile and will not compromise the main study in any way (e.g. by impairing recruitment or the ability of the participating hospitals to provide care to all patients under their care).

### 7 VERSION HISTORY

| Version number | Date | Brief Description of Changes |
|---|---|---|
| 1.0 | 13-Mar-2020 | Initial version |
| 2.0 | 21-Mar-2020 | Addition of hydroxychloroquine. Administrative changes and other clarifications. |
| 3.0 | 07-Apr-2020 | Extension of eligibility to those with suspected COVID-19<br>Addition of azithromycin arm.<br>Addition of inclusion of adults who lack permanently lack capacity.<br>Change to primary outcome from in-hospital death to death within 28 days of randomisation. |
| 4.0 | 14-Apr-2020 | Addition of second randomisation to tocilizumab vs. standard of care among patients with progressive COVID-19. |
| 5.0 | 24-Apr-2020 | Addition of children to study population. |
| 6.0 | 14-May-2020 | Addition of convalescent plasma |
| 7.0 | 18-Jun-2020 | Allowance of randomisation in part B of main randomisation without part A.<br>Removal of hydroxychloroquine and dexamethasone treatment arms. |
| 8.0 | 03-Jul-2020 | Removal of lopinavir-ritonavir<br>Addition of intravenous immunoglobulin arm for children<br>Changes to corticosteroid dosing for children.<br>Addition of baseline serum sample in convalescent plasma randomisation |
| 9.0 | 10-Sep-2020 | Addition of synthetic neutralizing antibodies<br>Additional baseline data collection<br>Addition of countries outside UK |
| 9.1 | 18-Sep-2020 | Addition of information about vaccination of children of pregnant mothers receiving REGN10933+REGN10987 |
| 9.2 [not submitted in UK] | 15-Oct-2020 | Additional information for countries outside UK |
| 10.0 | 26-Oct-2020 | Addition of main randomisation part C<br>General updates to avoid duplication and improve clarity |
| 10.1 | 01-Nov-2020 | Additional information for pregnant women |
| 11.0 | 19-Nov-2020 | Addition of colchicine to main randomisation part A<br>Removal of azithromycin from main randomization part A<br>Change in randomisation ratio in main randomisation part A from 2:1 to 1:1 |
| 11.1 | 21-Nov-2020 | Clarification of colchicine age thresholds |
| 11.2 [not submitted in UK] | 01-Dec-2020 | Addition of modified aspirin dose if 150mg not available |
| 12.0 | 10-Dec-2020 | Allow second randomisation of children without first randomisation |
| 12.1 | 16-Dec-2020 | Clarification of change in V12.0 |

### 8 APPENDICES

#### 8.1 Appendix 1: Information about the treatment arms

All patients will receive usual care in the participating hospital.

**[UK only] Corticosteroids:** Favourable modulation of the immune response is considered one of the possible mechanisms by which corticosteroids might be beneficial in the treatment of severe acute respiratory coronavirus infections, including COVID-19, SARS and MERS. Common to severe cases of these infections is the presence of hypercytokinemia (a cytokine 'storm') and development of acute lung injury or acute respiratory distress syndrome (ARDS).[13-16] Pathologically, diffuse alveolar damage is found in patients who die from these infections.[17] A growing volume of clinical trial data from patients with severe community acquired pneumonia, ARDS and septic shock suggest benefit from low-to-moderate dose corticosteroids in relation to mortality and length of stay.[18-20]

In trials of low-to-moderate doses of corticosteroids, the main adverse effect has been hyperglycaemia.[19,21] A systematic review of (mainly low-dose) corticosteroid trials in severe sepsis and septic shock did not identify any increased risk of gastroduodenal bleeding, superinfection or neuromuscular weakness; an association with an increased risk of hyperglycaemia (RR 1.16, 95% CI 1.07 to 1.25) and hypernatraemia (RR 1.61, 95% CI 1.26 to 2.06) was noted.[22]

Methylprednisolone is a corticosteroid with mainly glucocorticoid activity. It is used in the treatment of conditions in which rapid and intense corticosteroid effect is required. Its licensed indications for paediatrics include a wide range of conditions including inflammatory disorders, allergic disorders, draft rejection reactions, severe erythema multiforme, juvenile idiopathic arthritis, and many others. In the paediatric population, a dosage of 10 mg/kg/day to a maximum of 1 g/day for up to 3 days is recommended in the treatment of graft rejection reactions following transplantation. A higher dosage of 30 mg/kg/day to a maximum of 1 g/day for up to 3 days is recommended for the treatment of haematological, rheumatic, renal and dermatological conditions (Source: British National Formulary for Children). Storage should be as per conditions in the Summary of Product Characteristics.

PIMS-TS is associated with a hyper-inflammatory state with elevated ESR, C-reactive protein, D-dimers, lactate dehydrogenase, ferritin, and increased levels of pro-inflammatory cytokines including as IL-1 and IL-6. While there is a pharmacological basis for using high dose methylprednisolone, the Delphi consensus process conducted by NHS England identified equipoise for its use in the treatment of PIMS-TS.

**Colchicine:** Colchicine inhibits cellular transport and mitosis by binding to tubulin and preventing its polymerisation as part of the cytoskeleton transport system. As a consequence, colchicine has a wide range of anti-inflammatory effects, including inhibition of certain inflammasomes (cytosolic pattern recognition receptor systems that are activated in response to detection of pathogens in the cytosol).[23,24] There is evidence that inflammasomes are activated in COVID-19, and the degree of activation is correlated with disease severity.[25] Colchicine has been widely used for treatment of gout and pericarditis, and there is evidence of cardiovascular benefit in patients with coronary artery disease. The

UK COVID-19 Therapeutics Advisory Panel has recommended that RECOVERY assess colchicine.

**[UK only] Intravenous immunoglobulin (IVIg):** IVIg is human normal immunoglobulin, available in a number of different preparations in routine NHS practice. The NHS England consensus process has established intravenous immunoglobulin as the interim first line treatment in non-shocked COVID-associated PIMS-TS and also that there is need for evaluation of intravenous immunoglobulin and corticosteroid in the initial management of PIMS-TS. In the similar but different disease process known as Kawasaki Diseases, randomised controlled trials and meta-analyses have demonstrated that early recognition and treatment of KD with IVIg (and aspirin) reduces the occurrence of coronary artery aneurysms. Current published guidelines recommend a dose of 2 g/kg IVIg given as a single infusion, as this has been shown to reduce the coronary artery aneurysm rate compared to a lower divided dose regimen.[26]

IVIg is licensed for immunomodulation in adults, children and adolescents (0-18 years) in a number of clinical conditions including but not limited to primary immune thrombocytopenia, Guillain Barré syndrome, Kawasaki disease (in association with aspirin), chronic inflammatory demyelinating polyradiculoneuropathy and multifocal motor neuropathy.

**Tocilizumab** is a monoclonal antibody that binds to the receptor for IL-6, blocking IL-6 signalling and reduces inflammation. Tocilizumab is licensed for use in patients with rheumatoid arthritis and for use in people aged at least 2 years with chimeric antigen receptor (CAR) T cell-induced severe or life-threatening cytokine release syndrome.

Severe COVID-19 is associated with a hyper-inflammatory state with elevated ESR, C-reactive protein, D-dimers, lactate dehydrogenase, ferritin, and increased levels of pro-inflammatory cytokines including as IL-1 and IL-6.[4,9,27] There have been published and unpublished (pre-print) case series reports of the successful treatment of COVID-19 patients with IL-6 inhibitors.[27,28] IL-6 inhibitors have not been evaluated for the treatment of COVID-19 in randomised controlled trials.

**[UK only] Convalescent plasma:** Convalescent plasma treatment, containing high titres of polyclonal antibody, has been used to treat severe viral pneumonias. Many studies have been small or poorly controlled but have reported beneficial effects in avian influenza[29-31], influenza A (H1N1) infections in 1915-1917[32] and 2009/2010[33,34], and seasonal influenza B[35]. More relevant to SARS-CoV-2, a systematic review of convalescent plasma treatment in SARS-CoV infections in 2003 identified eight observational studies that all reported improved mortality associated with the use of convalescent plasma – infected patients received various amounts of convalescent plasma.[36] Recent studies in seasonal influenza A and in MERS-CoV highlight the importance of high avidity and high titre antibodies respectively.[37,38]

Convalescent plasma therapy had been given to at least 245 COVID-19 patients by the end of February 2020, and, according to a Chinese health official, 91 cases had shown improvement in clinical indicators and symptoms. Five small case series (26 patients in total) have been published that report the use of convalescent plasma in people with COVID-19 infection.[39-43] These studies have reported clinical and radiological improvements after treatment with convalescent plasma. However, these small uncontrolled studies have significant flaws and the reported effects are unreliable. Convalescent plasma is currently being tested in the REMAP-CAP trial among patients on intensive care units.

**[UK only] Synthetic neutralising antibodies (REGN-COV2):** Synthetic monoclonal antibodies (mAbs) have been demonstrated to be safe and effective in viral disease when used as prophylaxis (respiratory syncytial virus and rabies) and treatment (Ebola virus disease).[44,45] Anti-SARS-CoV-2 mAbs are designed to bind to and neutralise the virus. In addition, mAbs may have additional effector functions (antibody dependent phagocytosis and cytotoxicity) through binding to SARS-CoV-2 spike protein expressed on the surface of cells. Anti-SARS-CoV-2 spike protein neutralizing mAbs have demonstrated in vivo efficacy in both therapeutic and prophylactic settings in mouse, and non-human primates models, with decreases in viral load and lung pathology.[2,46,47]

Regeneron has developed 2 non-competing, high-affinity human IgG1 anti-SARS-CoV-2 mAbs, REGN10933 and REGN10987 that bind specifically to the receptor binding domain of the spike glycoprotein of SARS-CoV-2, blocking viral entry into host cells.[48,49] REGN10933 and REGN10987 are both potent neutralizing antibodies that block the interaction between the spike protein and its canonical receptor angiotensin-converting enzyme 2. REGN10933 and REGN10987 are intended to be utilized as a combination treatment, known as REGN-COV2, and should not be used individually as monotherapy. A combination of antibodies that bind to non-overlapping epitopes may minimize the likelihood of loss of antiviral activity due to naturally circulating viral variants or development of escape mutants under drug pressure. In animal studies (rhesus macaques and hamsters) the antibody cocktail (REGN10933+REGN10987) reduced virus load in lower and upper airway and decreased virus induced pathological sequelae when administered prophylactically or therapeutically.[50]

**Aspirin:** Patients with COVID-19 appear to be at high risk of thromboembolism.[51] Classical risk factors for thromboembolism are common in the COVID-19 hospitalised population, but the relatively low incidence of deep vein thrombosis compared to the incidence of pulmonary embolism (and the often peripheral location of the pulmonary emboli observed) suggests that inflammation and associated endothelial injury and platelet activation may be an important cause of thromboembolism in this patient population.[51,52] Therefore antiplatelet therapy is a potential thromboprophylactic therapy in COVID-19. It is also being tested in the REMAP-CAP trial.

#### 8.2 Appendix 2: Drug specific contraindications and cautions

**Corticosteroid (children only)**

- Known contra-indication to short-term corticosteroid.

**Colchicine (men ≥18 years old and women ≥55 years old only)**
Contraindications:

- Female participants <55 years old (as contraindicated in women of child-bearing potential)
- Severe hepatic impairment (defined as requiring ongoing specialist care)
- Significant cytopaenia (e.g. neutrophil count <1.0 x$10^9$/L; platelet count <50 x$10^9$/L; reticulocyte count <20 x$10^9$/L [if available])
- Concomitant use of strong CYP3A4 inhibitor (e.g. clarithromycin, erythromycin, systemic azole antifungal, HIV protease inhibitor) or P-gp inhibitor (e.g. ciclosporin, verapamil, quinidine).
- Hypersensitivity to lactose

Cautions: dose frequency should be halved (i.e. 1 mg at randomisation, 500 mcg 12 hours later then 500 mcg <u>once</u> daily) in the following circumstances.

- Concomitant use of moderate CYP3A4 inhibitor (e.g. diltiazem)
- Renal impairment: eGFR <30 mL/min/1.73$m^2$ (either chronic or acute)
- Estimated body weight <70 kg

(If >1 of these is present, investigator should consider not including colchicine in randomisation.)

Participants allocated colchicine should have full blood counts monitored at a frequency determined by their clinician.

**Intravenous Immunoglobulin (children only)**

- Hypersensitivity to the active substance (human immunoglobulins) or to any of the excipients
- Patients with selective IgA deficiency who developed antibodies to IgA, as administering an IgA-containing product can result in anaphylaxis
- Hyperprolinaemia type I or II.

Potential complications can often be avoided by ensuring that participants:

i. are carefully monitored for any symptoms throughout the infusion period;
ii. have urine output and serum creatinine levels monitored; and
iii. avoid concomitant use of loop diuretics.

Such monitoring should occur regularly during the admission, at a frequency appropriate to the illness of the child.

**Aspirin**

- Age <18 years old
- Known hypersensitivity to aspirin
- Recent major bleeding that precludes use of aspirin in opinion of managing physician

- Current use of aspirin, clopidogrel or other antiplatelet therapy

**Tocilizumab**

- Known hypersensitivity to tocilizumab.
- Evidence of active TB infection[i]
- Clear evidence of active bacterial, fungal, viral, or other infection (besides COVID-19)

(Note: Pregnancy and breastfeeding are not exclusion criteria.)

**Convalescent plasma**

- Known moderate or severe allergy to blood components*
- Not willing to receive a blood product*

**Synthetic neutralising antibodies (REGN-COV2)**

- Intravenous immunoglobulin treatment during current admission*
- Age <12 years old or child with weight <40kg*

(Note: Pregnancy and breastfeeding are not exclusion criteria.)

The infusion of synthetic neutralising antibodies should be interrupted if any of the following are observed (or worsen during the infusion): sustained/severe cough, rigors/chills, rash, pruritus, urticaria, diaphoresis, hypotension, dyspnoea, vomiting, or flushing. The reactions should be treated symptomatically, and the infusion may be restarted at 50% of the original rate once all symptoms have ceased (or returned to baseline) and at the discretion of the managing physician. If the managing physician feels there is medical need for treatment or discontinuation of the infusion other than described above, they should use clinical judgement to provide appropriate response according to typical clinical practice.

Pregnant women that are administered REGN10933 and REGN10987 must be advised that live vaccines should be avoided in children with *in utero* exposure to biologics for at least the first 6 months of life.

* If these conditions are recorded on the baseline case report form, patients will be ineligible for randomisation to that arm of the study.

Note: This study is being conducted within hospitals. Therefore use of medication will be subject to standard medication reviews (typically within 48 hours of enrolment) and clinical assessments (including appropriate blood tests) which will guide modifications to both the study treatment and use of concomitant medication (e.g. in the case of potential drug interactions). The doctor may decide whether it is appropriate to stop such medications temporarily to allow the patient to complete the course of their assigned intervention.

Although all available data on use in pregnancy are reassuring, since the effect of some of the treatments on unborn babies is uncertain, female participants who are not already pregnant will be advised that they should not get pregnant within 3 months of the completion of trial treatment(s).

[i] Note: The risk of reactivation of latent tuberculosis with tocilizumab is considered to be extremely small.

#### 8.3 Appendix 3: Paediatric dosing information

Children (aged <18 years old) will be recruited in the UK only.

##### Main Randomisation Part A

| Arm | Route | Weight # | Dose (Duration for all arms = 10 days or until discharge from hospital) |
|---|---|---|---|
| **No additional treatment** | - | - | - |
| **Corticosteroid**<br>- Solution for injection*<br>- Powder for solution for injection*<br>-<br>*various strengths available | Intravenous | All Including pre-term neonates | Neonates/infants with a corrected gestational age of ≤44 weeks<br><br>**Hydrocortisone** (IV):<br>0.5 mg/kg every 12 hours for 7 days and then 0.5mg/kg once daily for 3 days |
| | Intravenous | All | For all other children (with PIMS-TS):<br><br>**Methylprednisolone sodium succinate**<br>10 mg/kg (as base) once daily for 3 days (max 1 gram)<br><br>No additional oral corticosteroid should be prescribed to follow the 3 day treatment course. |
| **Human normal immunoglobulin (IVIg)**<br><br>- solution for infusion<br><br>*various strengths available | Intravenous | All | For children with corrected gestational age >44 weeks and <18 years with PIMS-TS phenotype:<br><br>2 g/kg as a single dose. (Dose should be based on ideal body weight in line with NHS England guidance.) |

### Weight to be rounded to the nearest kg unless dosage expressed as mg/kg or mL/kg.

#### Main Randomisation Part B

| | | | |
|---|---|---|---|
| **Convalescent Plasma** | Intravenous | | 5 mL/kg of ABO compatible convalescent plasma intravenous up to standard adult dose of 275 mLs per day on study days 1 and 2.<br><br>Minimum of 12 hour interval between 1st and 2nd units.<br><br>Convalescent plasma for neonates and infants up to one year of age needs to be ordered on a named patient basis from the relevant National Blood Service to ensure the unit meets neonatal requirements. Data transfer storage and retention will be in line with NHSBT standard procedures and protocols. |
| **Synthetic neutralising antibodies** (REGN10933 + REGN10987) | Intravenous | ≥12 years And ≥40 kg | 8 g (4 g of each monoclonal antibody) |

#### Second stage randomisation (Patients < 1 year of age will NOT be eligible)

| Arm | Route | Weight | Dose |
|---|---|---|---|
| **No additional treatment** | - | - | - |
| **Tocilizumab** | Intravenous | Infants < 1 year excluded | |
| | | < 30 kg | 12 mg/kg<br>A second dose may be given ≥12 and ≤24 hours later if, in the opinion of the attending clinicians, the patient's condition has not improved. |
| | | ≥ 30 kg | 8 mg/kg (max 800 mg)<br>A second dose may be given ≥12 and ≤24 hours later if, in the opinion of the attending clinicians, the patient's condition has not improved. |

#### 8.4 Appendix 4: Use of IMPs in pregnant and breastfeeding women

All trial drugs (except REGN-COV2) have been used in pregnant women with pre-existing medical disorders where benefits outweigh the risks to fetus or woman, including in the first trimester. The existing data related to each drug is summarized below.

**Colchicine**
Colchicine is contraindicated in pregnant or breastfeeding women.

**Convalescent plasma**
Convalescent plasma is plasma from people who had confirmed COVID-19 (SARS-Cov-2) infection, and have now recovered and been free of the infection for 28 days. The plasma contains antibodies that their immune systems have produced in fighting the virus. It is hoped that giving this plasma will help speed up recovery of a patient with active infection and improve their chances of survival. Plasma is already used as a treatment in pregnant patients who are bleeding,[53] or have particular blood conditions.[54,55] The plasma being used in this trial is from a selected donor and hopefully contains anti-SARS-Cov-2 antibodies, but is otherwise no different. Plasma infusions can occasionally cause side effects. Mostly this is a rise in temperature, itching or a rash, and in very extreme cases, anaphylaxis. Other potential complications include breathlessness and changes in blood pressure. Monitoring of pulse and blood pressure takes place before and after the infusion. There is no risk of miscarriage or fetal loss, preterm birth, preterm rupture of membranes, perinatal mortality or low birthweight, from plasma transfusions and there are no concerns with breast feeding.

**REGN-COV2 Monoclonal antibodies**
Monoclonal antibodies have been used as therapeutic agents in pregnancy over recent years, for a variety of conditions. Human monoclonal antibodies in use in pregnancy include anti-TNF agents, such as adalimumab, indicated for a variety of chronic inflammatory diseases such as rheumatoid arthritis and inflammatory bowel disease. Data have recently accumulated from a variety of cohort and registry studies indicating that such exposure in pregnancy was not associated with an increased risk for adverse pregnancy outcomes, when compared to unexposed pregnancies with the same underlying medical diseases.[56] This is supported by a consensus report on immunosuppressives and biologics during pregnancy and lactation, confirming no evidence of elevated adverse pregnancy outcomes or malformation risks.[57] Some monoclonal antibodies are transported across the placenta (and may also enter breast milk) but as REGN10933 and REGN10987 do not have any human targets such exposure should not be associated with risk of harm. Pregnant women, just like other patients with COVID-19, are at significant risk from the infection itself (particularly those in the third trimester.[58,59] All pregnant women in RECOVERY are entered into the UK Obstetric Surveillance System which follows all pregnancies to their conclusion.[59] Given the early safety experience with REGN10933+REGN10987 it would appear appropriate not to exclude pregnant women from this aspect of the trial (as such exclusion would inhibit the development of treatments for this population).[60]

**Aspirin**
Aspirin is widely used for the prevention of pre-eclampsia in pregnant women at increased risk of the disease. A recent Cochrane meta-analysis on this topic included seventy-seven trials (40,249 women) taking aspirin at doses between 60 and 150mg daily.[61] In most trials, aspirin was started from 12 weeks' gestation, although a more recent meta-analysis has reported eight trials (1426 women) in which aspirin was initiated in the first trimester.[62] In

light of the clear evidence of effectiveness, 75-150mg aspirin is recommended for pre-eclampsia prophylaxis in NICE guidelines for management of hypertension in pregnancy (NG133), and in the NHS England document ‘Saving Babies’ Lives for women at increased risk of placental dysfunction disorders.[63,64] There is some ongoing uncertainty as to the optimal dose (75mg vs. 150mg) for pre-eclampsia prophylaxis, but both doses are in widespread clinical use in pregnancy in the UK for these indications and in other conditions (e.g. in pregnant women with antiphospholipid syndrome).

**Tocilizumab**

Two pharmaceutical global safety registry database studies have reported on tocilizumab use in pregnancy, including outcomes from 288 pregnancies [65] and 61 pregnancies,[66] typically for rheumatoid or other arthritides, and with the majority having received the drug in the first trimester. These data suggest that the rates of congenital abnormality, spontaneous pregnancy loss and other adverse outcomes were not higher than in the general population.[66] Small studies have shown that tocilizumab is transferred to the fetus with serum concentrations approximately 7-fold lower than those observed in maternal serum at the time of birth.[67] Very low concentrations of tocilizumab are identified in breast milk and no drug is transferred into the serum of breast fed infants.[67,68] Women should be advised that if treated after 20 weeks’ gestation, their infant should not be immunised with live vaccines (rotavirus and BCG) for the first 6 months of life. All non-live vaccinations are safe and should be undertaken.[69]

#### 8.5 Appendix 5: Organisational Structure and Responsibilities

##### Chief Investigator

The Chief Investigator has overall responsibility for:

(i) Design and conduct of the Study in collaboration with the Trial Steering Committee;
(ii) Preparation of the Protocol and subsequent revisions;

##### Trial Steering Committee

The Trial Steering Committee (see Section 8.6 for list of members) is responsible for:

(i) Agreement of the final Protocol and the Data Analysis Plans;
(ii) Reviewing progress of the study and, if necessary, deciding on Protocol changes;
(iii) Review and approval of study publications and substudy proposals;
(iv) Reviewing new studies that may be of relevance.

##### Regional (South East Asia) Steering Committee

The regional SEA Steering Committee (see Section 8.6 for list of members) is responsible for:

(i) Reviewing progress of the study in South East Asia;
(ii) Review of study publications and substudy proposals;
(iii) Considering potential new therapies to be included in South East Asia;
(iv) Assisting RCC in selection of LCCs
(v) Reviewing new studies that may be of relevance.

##### Data Monitoring Committee

The independent Data Monitoring Committee is responsible for:

(i) Reviewing unblinded interim analyses according to the Protocol;
(ii) Advising the Steering Committee if, in their view, the randomised data provide evidence that may warrant a change in the protocol (e.g. modification or cessation of one or more of the treatment arms).

##### Central Coordinating Office (CCO)

The CCO is responsible for the overall coordination of the Study, including:

(i) Study planning and organisation of Steering Committee meetings;
(ii) Ensuring necessary regulatory and ethics committee approvals;
(iii) Development of Standard Operating Procedures and computer systems
(iv) Monitoring overall progress of the study;
(v) Provision of study materials to RCCs/LCCs;
(vi) Monitoring and reporting safety information in line with the protocol and regulatory requirements;
(vii) Dealing with technical, medical and administrative queries from LCCs.

**Regional Coordinating Centre (RCC)**
The RCCs are responsible for:

(i) Ensuring necessary regulatory and ethics committee approvals;
(ii) Provision of study materials to LCCs;
(iii) Dealing with technical, medical and administrative queries from LCCs.

**Local Clinical Centres (LCC)**
The LCC lead investigator and LCC clinic staff are responsible for:

(i) Obtaining all relevant local permissions (assisted by the CCO)
(ii) All trial activities at the LCC, including appropriate training and supervision for clinical staff
(iii) Conducting trial procedures at the LCC in line with all relevant local policies and procedures;
(iv) Dealing with enquiries from participants and others.

#### 8.6 Appendix 5: Organisational Details

##### STEERING COMMITTEE

(Major organisational and policy decisions, and scientific advice; blinded to treatment allocation)

| | |
|---|---|
| Chief Investigator | Peter Horby |
| Deputy Chief Investigator | Martin Landray |
| Clinical Trial Unit Lead | Richard Haynes |
| Co-investigators | Kenneth Baillie (Scotland Lead), Maya Buch, Lucy Chappell, Saul Faust, Thomas Jaki, Katie Jeffery, Edmund Juszczak, Wei Shen Lim, Marion Mafham, Alan Montgomery, Andrew Mumford, Kathy Rowan, Guy Thwaites, Jeremy Day (South East Asia Leads) |

##### South East Asia Steering Committee (Members TBD)

| | |
|---|---|
| Regional Lead Investigators | Guy Thwaites, Jeremy Day |
| Country Lead Investigators: | TBD (Nepal), TBD (VietNam), TBD (Indonesia) |
| MOH or local country representatives: | TBD (Nepal), TBD (VietNam), TBD (Indonesia) |
| Independent members: | TBD (3 members) |

##### DATA MONITORING COMMITTEE

(Interim analyses and response to specific concerns)

| | |
|---|---|
| Chair | Peter Sandercock |
| Members | Janet Darbyshire, David DeMets, Robert Fowler, David Lalloo, Ian Roberts, Janet Wittes |
| Statisticians (non-voting) | Jonathan Emberson, Natalie Staplin |

### 10 CONTACT DETAILS

Website: www.recoverytrial.net
(copies of this protocol and related forms and information can be downloaded)

**RECOVERY Central Coordinating Office:**
Richard Doll Building, Old Road Campus, Roosevelt Drive, Oxford OX3 7LF
United Kingdom

**RECOVERY Vietnam:**
Oxford University Clinical Research Unit, Centre for Tropical Medicine, 764 Vo Van Kiet, District 5, Ho Chi Minh City, Vietnam


**RECOVERY Indonesia:**
Eijkman Oxford Clinical Research Unit (EOCRU), Eijkman Institute for Molecular Biology
Jl. P. Diponegoro No. 69, Jakarta-Indonesia 10430

**RECOVERY Nepal:**
Clinical Trial Unit, Oxford University Clinical Research Unit-Nepal, Patan Academy of Health Sciences, Kathmandu, Nepal

**To RANDOMISE a patient, visit:**

---

RECOVERY
Randomised Evaluation of COVID-19 Therapy

Website: www.recoverytrial.net

---

#### Appendix 2: RECOVERY Trial Statistical Analysis Plan V2.1

### RECOVERY

Randomised Evaluation of COVID-19 Therapy

### Statistical Analysis Plan

**Version 2.1**

**Date: 02 December 2020**

Aligned with protocol version: 11.1, 21 November 2020

IRAS no: 281712
REC ref: EE/20/0101
ISRCTN: 50189673
EudraCT: 2020-001113-21

Nuffield Department of
POPULATION HEALTH

UNIVERSITY OF OXFORD

#### Abbreviations

| | |
|---|---|
| ADaM | Analysis Data Model |
| AE | Adverse event |
| CDISC | The Clinical Data Interchange Standards Consortium |
| CI | Confidence interval |
| COVID | Coronavirus-induced disease |
| CPAP | Continuous Positive Airway Pressure |
| CRP | C-reactive protein |
| DMC | Data Monitoring Committee |
| ECMO | Extra Corporeal Membrane Oxygenation |
| eCRF | Electronic case report form |
| ICD | International Classification of Diseases |
| ICNARC | Intensive Care National Audit and Research Centre |
| ITT | Intention to treat |
| MedDRA | Medical Dictionary for Regulatory Activities |
| OPCS-4 | National Health Service OPCS Classification of Interventions and Procedures version 4 |
| SARS | Severe acute respiratory syndrome |
| SARS-CoV-2 | Severe acute respiratory syndrome coronavirus 2 |
| SSAR | Suspected serious adverse reaction |
| SUSAR | Suspected unexpected serious adverse reaction |
| TSC | Trial Steering Committee |

#### List of authors and reviewers (up to and including SAP version 1.1)

##### *Authors*

Dr Louise Linsell, Lead Trial Statistician, Nuffield Department of Population Health (NDPH), University of Oxford

Jennifer Bell, Trial Statistician, NDPH, University of Oxford

##### *Reviewers*

Professor Jonathan Emberson, Data Monitoring Committee (DMC) Statistician, NDPH, University of Oxford (prior to unblinded interim analysis of trial outcomes)

Professor Richard Haynes, Clinical Coordinator, NDPH, University of Oxford

Professor Peter Horby, Chief Investigator (CI), Nuffield Department of Medicine, University of Oxford

Professor Thomas Jaki, TSC Member, Department of Mathematics and Statistics, Lancaster University

Associate Professor Edmund Juszczak, TSC Member, NDPH, University of Oxford (until 6 July 2020)

Professor Martin Landray, Deputy CI, NDPH, University of Oxford

Professor Alan Montgomery, TSC Member, Nottingham Clinical Trials Unit, University of Nottingham

Dr Natalie Staplin, DMC Statistician, NDPH, University of Oxford (prior to unblinded interim analysis of trial outcomes)

#### List of authors and reviewers (version 2.0 onwards)

##### *Authors*

Professor Edmund Juszczak, TSC Member (University of Nottingham from 6 July 2020)

Professor Alan Montgomery (University of Nottingham), TSC Member

Professor Thomas Jaki (University of Cambridge) co-investigator and TSC Member

##### *Reviewers*

Enti Spata, Jennifer Bell, Dr Louise Linsell, Trial Statisticians, NDPH, University of Oxford

Professor Richard Haynes, Clinical Coordinator, NDPH, University of Oxford

Professor Martin Landray, Deputy CI, NDPH, University of Oxford

Professor Peter Horby, CI, Nuffield Department of Medicine, University of Oxford

#### Roles and responsibilities

##### *Trial Statisticians*

*Until 30th September 2020:* Dr Louise Linsell and Jennifer Bell (NDPH, University of Oxford)

Role: To develop the statistical analysis plan (blinded to trial allocation) and conduct the final comparative analyses for Lopinavir-Ritonavir¸ Corticosteroid (dexamethasone) and Hydroxychloroquine (main randomisation part A).

*From 1st October 2020:* Enti Spata (NDPH, University of Oxford)

Role: To develop the statistical analysis plan (blinded to trial allocation) and conduct the final comparative analyses for all other treatment arms.

##### *Data Monitoring Committee (DMC) Statisticians*

Professor Jonathan Emberson and Dr Natalie Staplin (NDPH, University of Oxford)

Role: To conduct regular interim analyses for the DMC. Contribution restricted up until unblinded to trial allocation.

##### *Statisticians on the Trial Steering Committee (TSC)*

Professor Edmund Juszczak (University of Nottingham), Professor Alan Montgomery (University of Nottingham), and Professor Thomas Jaki (University of Cambridge)

Role: Major organisational and policy decisions, and scientific advice; blinded to treatment allocation.

##### *Trial IT systems & Programmers*

Andy King, David Murray, Richard Welsh (NDPH, University of Oxford)

Role: To generate and prepare reports monitoring the randomisation schedule. To supply data snapshots for interim and final analysis. Responsibility for randomisation system, clinical databases and related activities.

Bob Goodenough (NDPH, University of Oxford)

Role: Validation of IT systems

Dr Will Stevens, Karl Wallendszuz (NDPH, University of Oxford)

Role: To produce analysis-ready datasets according to CDISC standards.

### 1 INTRODUCTION

This document details the proposed presentation and analysis for the main paper(s) reporting results from the multicentre randomised controlled trial RECOVERY (ISRCTN50189673) to investigate multiple treatments on major outcomes in inpatients for COVID-19 (clinically suspected or laboratory confirmed).

The results reported in these papers will follow the strategy set out here, which adheres to the guidelines for the content of a statistical analysis plan (SAP).[1] Any subsequent analyses of a more exploratory nature will not be bound by this strategy.

Suggestions for subsequent analyses by oversight committees, journal editors or referees, will be considered carefully in line with the principles of this analysis plan.

Any deviations from the statistical analysis plan will be described and justified in the final report. The analysis will be carried out by identified, appropriately qualified and experienced statisticians, who will ensure the integrity of the data during their processing.

This SAP is based on multiple versions of the protocol. All regulatory documents can be found in the RECOVERY trial directory: https://www.recoverytrial.net/for-site-staff/site-set-up-1/regulatory-documents.

SAP versions 1.0 & 1.1 applied to the first three principal comparisons (hydroxychloroquine, dexamethasone, and lopinavir-ritonavir versus no additional treatment respectively), for which data matured in the first UK wave of the pandemic. However, due to its later introduction, enrolment of patients in the azithromycin arm was much slower. Over time, factorial randomisations and a second randomisation have been added, introducing new treatment arms including convalescent plasma, tocilizumab, synthetic neutralizing antibodies, and aspirin. These changes, combined with the fact that use of corticosteroids (one of the original treatment arms) is now the usual standard of care for many patients, makes this a sensible juncture to re-evaluate the SAP and produce version 2.0.

### 2 BACKGROUND INFORMATION

#### 2.1 Rationale

In early 2020, as the protocol was being developed, there were no approved treatments for COVID-19. The aim of the trial is to provide reliable evidence on the efficacy of candidate therapies (including re-purposed and novel drugs) for suspected or confirmed COVID-19 infection on major outcomes in hospitalised adult patients receiving standard care.

#### 2.2 Objectives of the trial

##### 2.2.1 *Primary objective*

To provide reliable estimates of the effect of study treatments on all-cause mortality within 28 days of the relevant randomisation.

##### 2.2.2 *Secondary objectives*

To investigate the effect of study treatments on the duration of hospital stay and on the combined endpoint of use of invasive mechanical ventilation (including Extra Corporal Membrane Oxygenation [ECMO]) or death.

#### 2.3 Trial design

This is a multi-centre, multi-arm, adaptive, open label, randomised controlled trial with three possible stages of randomisation, as described below. The trial is designed with streamlined processes in order to facilitate rapid large-scale recruitment with minimal data collection.

#### 2.4 Eligibility

##### 2.4.1 *Inclusion criteria*

Patients are eligible for the trial if all of the following are true:

- Hospitalised
- SARS-Cov-2 infection (clinically suspected or laboratory confirmed)
- No medical history that might, in the opinion of the attending clinician, put the patient at significant risk if they were to participate in the trial.

##### 2.4.2 *Exclusion criteria*

If one or more of the active drug treatments is not available at the hospital or is believed, by the attending clinician, to be contraindicated (or definitely indicated) for the specific patient, then this fact will be recorded via the web-based form prior to randomisation; random allocation will then be between the remaining arms.

#### 2.5 Treatments

All patients will receive standard management for the participating hospital. The main randomisation will be between the following treatment arms (although not all arms may be available at any one time). The doses listed are for adults; paediatric dosing is described in the protocol.

##### 2.5.1 *Main randomisation part A:*

- **No additional treatment**
- **Lopinavir 400mg-Ritonavir 100mg** by mouth (or nasogastric tube) every 12 hours for 10 days. [Introduced in protocol version 1.0; **enrolment closed** 29 June 2020]

- **Corticosteroid** in the form of dexamethasone**,** administered as an oral liquid or intravenous preparation 6 mg once daily for 10 days. In pregnancy, prednisolone 40 mg administered by mouth (or intravenous hydrocortisone 80 mg twice daily) should be used instead. [Introduced in protocol version 1.0; **enrolment closed to adults** 8 June 2020]
- **Hydroxychloroquine** by mouth for 10 days (4 doses in first 24 hours and 1 dose every 12 hours for 9 days). [Introduced in protocol version 2.0; **enrolment closed** 5 June 2020]
- **Azithromycin 500mg** by mouth (or nasogastric tube) or intravenously once daily for a total of 10 days. [Introduced in protocol version 3.0; enrolment closed 27 November 2020]
- **Colchicine** by mouth for 10 days (1.5 mg in first 12 hours then 0.5 mg twice daily)

##### 2.5.2 *Main randomisation part B:*

In a factorial design, eligible patients may be randomised to the arms below. The doses listed are for adults; paediatric dosing is described in the protocol.

- **No additional treatment**
- **Convalescent plasma** Single unit of ABO compatible convalescent plasma (275mls ± 75 mls) intravenous per day on study days 1 (as soon as possible after randomisation) and 2 (with a minimum of 12-hour interval between 1st and 2nd units). ABO identical plasma is preferred if available. The second transfusion should not be given if patient has a suspected serious adverse reaction during or after the first transfusion. [Introduced in protocol version 6.0; enrolment ongoing]
- **Synthetic neutralising antibodies** (REGN-COV2; adults and children aged ≥12 years only - children who weigh <40kg will also not be eligible for this treatment). A single dose of REGN10933 + REGN10987 8 g (4 g of each monoclonal antibody) in 250ml 0.9% saline infused intravenously over 60 minutes ± 15 minutes as soon as possible after randomisation. [Introduced in protocol version 9.1; enrolment ongoing]

##### 2.5.3 *Main randomisation part C:*

In a factorial design, eligible patients may be randomised to the arms below. The dose listed is for adults; children are excluded from this comparison.

- **No additional treatment**
- **Aspirin** 150 mg by mouth (or nasogastric tube) or per rectum once daily until discharge. [Introduced in protocol version 10.1; enrolment ongoing]

##### 2.5.4 *Second randomisation for patients with progressive COVID-19*

Patients enrolled in the main RECOVERY trial and with clinical evidence of a hyper-inflammatory state may be considered for a second randomisation if they meet the following criteria:

- Randomised into the main RECOVERY trial no more than 21 days ago
- Clinical evidence of progressive COVID-19:
  - oxygen saturation <92% on room air or requiring oxygen (or in children, significant systemic disease with persistent pyrexia, with or without evidence of respiratory involvement); and
  - C-reactive protein (CRP) ≥75 mg/L
- No medical history that might, in the opinion of the attending clinician, put the patient at significant risk if they were to participate in this aspect of the RECOVERY trial

Eligible participants may be randomised between the following treatment arms:

- **No additional treatment**
- **Tocilizumab** by intravenous infusion with the dose determined by body weight. [Introduced in protocol version 4.0; enrolment ongoing]

#### 2.6 Definitions of primary and secondary outcomes

Outcomes will be assessed at 28 days and then 6 months after the relevant randomisation. Analysis of longer-term outcomes collected beyond this will be described in a separate Statistical Analysis Plan.

##### 2.6.1 *Primary outcome*

Mortality (all-cause)

##### 2.6.2 *Secondary clinical outcomes*

- Time to discharge from hospital
- Use of invasive mechanical ventilation (including Extra Corporal Membrane Oxygenation [ECMO]) or death (among patients not on invasive mechanical ventilation or ECMO at time of randomisation)

##### 2.6.3 *Subsidiary clinical outcomes*

- Use of ventilation (overall and by type) among patients not on ventilation (of any type) at time of randomisation
- Duration of invasive mechanical ventilation among patients on invasive mechanical ventilation at time of randomisation (defined as time to successful cessation of invasive mechanical ventilation: see section 5.1.3.2)
- Use of renal dialysis or haemofiltration (among patients not on renal dialysis or haemofiltration at time of randomisation)
- Thrombotic events (overall and by type; introduced in Protocol version 10.1)

##### 2.6.4 *Safety outcomes*

- Cause-specific mortality (COVID-19, other infection, cardiac, stroke, other vascular, cancer, other medical, external, unknown cause)

- Major cardiac arrhythmia (recorded on follow-up forms completed from 12 May 2020 onwards)
- Major bleeding (overall and by type; introduced in Protocol version 10.1)
- Early safety of antibody-based therapy (sudden worsening in respiratory status; severe allergic reaction; temperature >39°C or ≥2°C rise since randomisation; sudden hypotension; clinical haemolysis; and thrombotic events within the first 72 hours; Main randomization phase B only)

###### 2.6.5 *Detailed derivation of outcomes*

The detailed derivation of outcomes included in statistical analysis will be described separately in a data derivation document and included in the Study Data Reviewer's Guide.

##### 2.7 Hypothesis framework

For each of the primary, secondary and subsidiary outcomes, the null hypothesis will be that there is no true difference in effect between any of the treatment arms.

##### 2.8 Sample size

The larger the number randomised, the more accurate the results will be, but the numbers that can be randomised will depend critically on how large the epidemic becomes. If substantial numbers are hospitalised in the participating centres then it may be possible to randomise several thousand with moderate disease and a few thousand with severe disease. Some indicative sample sizes and projected recruitment will be estimated using emerging data for several different scenarios. Sample size and recruitment will be monitored by the TSC throughout the trial.

##### 2.9 Randomisation

Eligible patients will be randomised using a 24/7 secure central web-based randomisation system, developed and hosted within NDPH, University of Oxford. Users of the system will have no insight into the next allocation, given that simple randomisation is being used. If a patient is randomised inadvertently more than once during the same hospital admission, the first allocation will be used.

The implementation of the randomisation procedure will be monitored by the Senior Trials Programmer, and the TSC notified if an error in the randomisation process is identified.

###### 2.9.1 *Main randomisation part A*

Simple randomisation will be used to allocate participants to one of the following treatment arms (in addition to usual care), which is subject to change:

- No additional treatment
- Lopinavir-Ritonavir [Introduced in protocol version 1.0; **enrolment closed** 29 June 2020]
- Corticosteroid [Introduced in protocol version 1.0; **enrolment closed to adults** 8 June 2020]
- Hydroxychloroquine [Introduced in protocol version 2.0; **enrolment closed** 5 June 2020]

- Azithromycin [Introduced in protocol version 3.0; **enrolment closed 27 November 2020**]
- Colchicine [Introduced in protocol version 11.1]

The randomisation programme will allocate patients in a ratio of 2:1 between the no additional treatment arm and each of the other arms that are not contra-indicated and are available. Hence if all 4 active treatment arms are available, then the randomisation will be in the ratio 2:1:1:1:1. If one or more of the active drug treatments is not available at the hospital or is believed, by the attending clinician, to be contraindicated (or definitely indicated) for the specific patient, then this fact will be recorded via the web-based form prior to randomisation; random allocation will then be between the remaining arms (in a 2:1:1:1, 2:1:1 or 2:1 ratio).

##### 2.9.2 *Main randomisation part B*

In a factorial design, eligible patients will be randomised simultaneously using simple randomisation with allocation ratio 1:1:1 to one of the following arms, which is subject to change:

- No additional treatment
- Convalescent plasma [Introduced in protocol version 6.0; enrolment ongoing]
- Synthetic neutralising antibodies [Introduced in protocol version 9.1; enrolment ongoing]

If the active treatment is not available at the hospital, the patient does not consent to receive convalescent plasma, or is believed, by the attending clinician, to be contraindicated for the specific patient, then this fact will be recorded via the web-based form and the patient will be excluded from the relevant arm in Randomisation part B.

##### 2.9.3 *Main randomisation part C*

In a factorial design, eligible patients will be randomised simultaneously using simple randomisation with allocation ratio 1:1 to one of the following arms, which is subject to change:

- No additional treatment
- Aspirin [Introduced in protocol version 10.1; enrolment ongoing]

Note: From protocol version 7.0 onwards, randomisation is permitted in part B of main randomisation without randomisation in part A. From protocol version 10.1 onwards, randomisation is permitted in any combination of parts A, B, and C.

##### 2.9.4 *Second randomisation for patients with progressive COVID-19*

Eligible participants will be randomised using simple randomisation with an allocation ratio 1:1 between the following arms, which is subject to change:

- No additional treatment
- Tocilizumab [Introduced in protocol version 4.0; enrolment ongoing]

#### 2.10 Blinding

This is an open-label study. However, while the study is in progress, access to tabular results of study outcomes by treatment allocation will not be available to the research team, CIs, trial statisticians, clinical teams, or members of the TSC (unless the DMC advises otherwise). The DMC and DMC statisticians will be unblinded.

#### 2.11 Data collection schedule

Baseline and outcome information will be collected on trial-specific electronic case report forms (eCRFs) and entered into a web-based IT system by a member of the hospital or research staff. Follow-up information will be collected on all study participants, irrespective of whether they complete the scheduled course of allocated study treatment. Study staff will seek follow-up information through various means, including routine healthcare systems and registries.

All randomised participants will be followed up until death or 6 months post-randomisation to the main trial (whichever is sooner). NHS Digital and equivalent organisations in the devolved nations will supply data fields relevant to trial baseline and outcome measures to NDPH, University of Oxford on a regular basis, for participants enrolled into the trial. This will be combined with the trial-specific data collected via the web-based IT system and adjudicated internally.

Longer term (up to 10 years) follow-up will be sought through linkage to electronic healthcare records and medical databases including those held by NHS Digital, Public Health England and equivalent bodies, and to relevant research databases (e.g. UK Biobank, Genomics England).

#### 2.12 Data monitoring

During the study all study data will be supplied in strict confidence to the independent DMC for independent assessment and evaluation. The DMC will request such analyses at a frequency relevant to the emerging data from this and other studies.

The DMC has been requested to determine if, in their view, the randomised comparisons in the study have provided evidence on mortality that is strong enough (with a range of uncertainty around the results that is narrow enough) to affect national and global treatment strategies. Hence, multiple reviews by the Data Monitoring Committee have no material impact on the final analysis. In such a circumstance, the DMC will inform the TSC who will make the results available to the public and amend the trial arms accordingly.

#### 2.13 Trial reporting

The trial will be reported according to the principles of the CONSORT statements.[2, 3, 4] The exact composition of the trial publication(s) depends on the size of the epidemic, the availability of drugs, and the findings from the various pairwise comparative analyses (with the no additional treatment arm) in the main trial.

### 3 ANALYSIS POPULATIONS

#### 3.1 Population definitions

The intention to treat (ITT) population will be all participants randomised, irrespective of treatment received. This ITT population will be used for analysis of efficacy and safety data. For interim analyses, baseline data will be reported for all participants with data available and outcome data will be reported for all participants who have died, been discharged from hospital, or reached day 28 after the first randomisation.

### 4 DESCRIPTIVE ANALYSES

#### 4.1 Participant throughput

The flow of participants through the trial will be summarised for each separate pairwise comparison using a CONSORT diagram. The flow diagram will show the contribution of participants from each of the paths (from each of the parts of the main randomisation and from the second randomisation), where applicable. The flow diagrams will describe the numbers of participants randomly allocated, who received allocation, withdrew consent, and included in the ITT analysis population. The flow diagrams for arms in the main randomisation will also report the number of participants who underwent the second randomisation.

#### 4.2 Baseline comparability of randomised groups

The following characteristics will be described separately for patients randomised to each main comparison (for each separate pairwise comparison of active treatment with the no additional treatment arm), and separately for the first and second randomisation.

##### 4.2.1 Main randomisation (parts A, B and C)

- Age at randomisation
- Sex
- Ethnicity
- Region (UK, South East Asia)
- Time since COVID-19 symptoms onset
- Time since hospitalisation
- Current respiratory support
- Comorbidities (diabetes, heart disease, chronic lung disease, tuberculosis, human immunodeficiency virus, severe liver disease, severe kidney impairment)
- SARS-Cov-2 test result
- If female, known to be pregnant
- Use of systemic corticosteroid (including those allocated to corticosteroid in part A)
- Use of other relevant treatments (e.g. remdesivir, antiplatelet treatment, anticoagulant treatment)
- For part B only, anti-SARS-CoV-2 antibody concentration
- For treatment comparisons introduced in protocol v9.1 onwards:
    - C-reactive protein
    - Estimated glomerular filtration rate (calculated using the CKD-EPI formula)
    - D-dimer

###### 4.2.2 Second randomisation

In addition to the above:

- Current respiratory support
- Latest oxygen saturation measurement
- Latest C-reactive protein
- Latest ferritin
- Latest estimated glomerular filtration rate (calculated using the CKD-EPI formula)
- Allocation in main randomisation parts A, B, and C
- Interval between first and second randomisation

The number and percentage will be presented for binary and categorical variables. The mean and standard deviation or the median and the interquartile range will be presented for continuous variables.

##### 4.3 Completeness of follow-up

All reasonable efforts will be taken to minimise loss to follow-up, which is expected to be minimal as data collection for primary and secondary outcomes using trial-specific eCRFs is combined with linkage to routine clinical data on study outcomes from NHS Digital, ICNARC, and similar organisations in the devolved nations.

The number and percentage of participants with follow-up information at day 28 and at 6 months after the relevant randomisation will be reported. Data will be shown for each of the following: all-cause mortality, hospital discharge status, ventilation status, and will be shown for each randomised group for the main and second randomisation separately.

##### 4.4 Adherence to treatment

The number and proportion of patients who did not receive the treatment they were allocated to will be reported. If any other trial treatment options were known to be received, instead of or in addition to, the allocated treatment during the 28-day follow-up period after the first randomisation, these will be collected and reported. Details on the number of days (or doses) of treatment received will be reported for all trial treatments received where available.

#### 5 COMPARATIVE ANALYSES

For all outcomes, the primary analysis will be performed on the intention to treat (ITT) population at 28 days after randomisation. An ITT analysis of all outcomes at 6 months post-randomisation will also be conducted.

Pairwise comparisons will be made between each treatment arm and the no additional treatment arm (reference group) in that particular randomisation (main randomisation part A, main randomisation part B, main randomisation part C, and second randomisation). Since not all treatments may be available or suitable for all patients, those in the no additional treatment arm will only be included in a given comparison if, at the point of their randomisation, they *could* alternatively have been randomised to the active treatment of interest (i.e. the active treatment was available at the time and it was not contra-indicated). The same applies to treatment arms added at a later stage; they will only be compared to those patients recruited concurrently.

#### 5.1 Main randomisation part A

##### 5.1.1 *Primary outcome*

Mortality (all-cause) will be summarised with counts and percentages by randomised comparison group. A time-to-event analysis will be conducted using the log-rank test, with the p-value reported. Kaplan-Meier estimates for the time to event will also be plotted (with associated log-rank p-values). The log-rank 'observed minus expected' statistic (and its variance) will be used to calculate the one-step estimate of the event rate ratio and confidence interval for each treatment group versus the no additional treatment group.[5] For the primary outcome, discharge alive before the relevant time period (28 days after randomisation) will be assumed as absence of the event (unless there is additional data confirming otherwise).

##### 5.1.2 *Secondary outcomes*

###### *5.1.2.1 Time to discharge alive from hospital*

A time-to-event analysis will be used to compare each treatment group with the no additional treatment group using the log-rank test. As described for the primary outcome, the rate ratio and its confidence interval will be estimated from the log-rank observed minus expected statistic and its variance, and Kaplan-Meier curves will be drawn. Patients who die in hospital will be censored after 28 days after randomisation. This gives an unbiased estimate of the recovery rate and comparable estimates to the competing risks approach in the absence of other censoring (which is expected to be very minimal).[6]

###### *5.1.2.2 Use of invasive mechanical ventilation (including ECMO) or death*

Counts and percentages will be presented by randomised group and the risk ratio will be calculated for each pairwise comparison with the no additional treatment arm, with confidence intervals and p-values reported. The absolute risk difference will also be presented with confidence intervals. Each component of this composite outcome will also be summarised. Patients who were already on invasive mechanical ventilation or ECMO at randomisation will be excluded from these analyses.

##### 5.1.3 *Subsidiary clinical outcomes*

###### *5.1.3.1 Use of ventilation (overall and by type)*

Counts and percentages will be presented by randomised group for patients who received any assisted ventilation, together with risk ratios and confidence intervals for each pairwise comparison with the no additional treatment arm. The number of patients receiving the two main types of ventilation will also be reported: non-invasive ventilation (including CPAP, other non-invasive ventilation or high-flow nasal oxygen), and invasive mechanical ventilation (including ECMO). Patients who were already receiving ventilation[a] at randomisation will be excluded from these analyses.

###### *5.1.3.2 Duration of invasive mechanical ventilation (time to successful cessation of invasive mechanical ventilation)*

Successful cessation of invasive mechanical ventilation will be defined as removal of invasive mechanical ventilation within (and survival to) 28 days after randomisation. A time-to-event analysis will be used to compare each treatment group with the no additional treatment group using the log-rank test, as described above. The rate ratio and its confidence interval will be estimated from the log-rank observed minus expected statistic and its variance, and Kaplan-Meier curves will be drawn. Patients who die within 28 days of randomisation will be censored *after* 28 days after randomisation. Patients who were not already on invasive mechanical ventilation or ECMO at randomisation will be excluded from these analyses.

###### *5.1.3.3 Use of renal dialysis or haemofiltration*

Counts and percentages will be presented by randomised group and the risk ratio will be calculated for each pairwise comparison with the no additional treatment arm, with confidence intervals and p-values reported. The absolute risk difference will also be presented with confidence intervals. Patients who were already on renal dialysis or haemofiltration at randomisation will be excluded from these analyses.

###### *5.1.3.4 Thrombotic event*

Counts and percentages will be presented by randomised group. The absolute risk differences will also be presented with confidence intervals. Type of thrombotic event will also be described: (i) acute pulmonary embolism; (ii) deep vein thrombosis; (iii) ischaemic stroke, (iv) myocardial infarction; (v) systemic arterial embolism; and (vi) all sites combined.

#### 5.2 Main randomisation part B

In the factorial design, the main effects of treatments evaluated in part B will be presented and tested across all arms in main randomisation parts A and C combined, as described in 5.1. (Assessments of whether the effects of treatments in part B vary depending on other randomised treatments are described in section 5.7).

#### 5.3 Main randomisation part C

In the factorial design, the main effects of treatments evaluated in part C will be presented and tested across all arms in main randomisation parts A and B combined, as described in 5.1. (Assessments of whether the effects of treatments in part C vary depending on other randomised treatments are described in section 5.7).

#### 5.4 Second randomisation

[a] For comparisons introduced to the main randomisation prior to protocol version 9.1, patients who were already receiving oxygen at randomisation will also be excluded from these analyses (since it is not possible to distinguish those who were already receiving non-invasive ventilation).

Evaluation of treatment effects in the main randomisation and the second randomisation will be conducted independently, as described in 5.1.

#### 5.5 Pre-specified subgroup analyses

Pre-specified subgroup analyses will be conducted for the main randomisation (parts A, B and C) and the second randomisation, for the following outcomes:

- Mortality (all-cause)
- Time to discharge from hospital
- Use of invasive mechanical ventilation (including ECMO) or death

Tests for heterogeneity (or tests for trend for 3 or more ordered groups) will be conducted to assess whether there is any good evidence that the effects in particular subgroups differ materially from the overall effect seen in all patients combined. Results will be presented on forest plots as event rate ratios, or risk ratios, with confidence intervals. The following subgroups will be examined based on information at randomisation:

- Age (<70; 70-79; 80+ years)
- Sex (Male; Female)
- Ethnicity (White; Black, Asian or Minority Ethnic)
- Region (UK, South East Asia)
- Time since illness onset (≤7 days; >7 days)
- Requirement for respiratory support
  - For main randomisation: None; Oxygen only (with or without non-invasive ventilation); Invasive mechanical ventilation (including ECMO)
  - For second randomisation: No ventilator support (including no or low-flow oxygen); Non-invasive ventilation (including CPAP, other non-invasive ventilation, or high-flow nasal oxygen), Invasive mechanical ventilation (including ECMO)
- Use of systemic corticosteroid (including dexamethasone)
- For part B only: Recipient anti-SARS-CoV-2 antibody concentration at randomisation (<8 x$10^6$ units; ≥8 x$10^6$ units[b])

#### 5.6 Sensitivity analyses

Sensitivity analyses of the primary and secondary outcomes will be conducted among those patients with a positive test for SARS-COV-2 (i.e. confirmed cases).

#### 5.7 Other exploratory analyses

In addition, exploratory analyses will be conducted to test for interactions between treatments allocated in each of the different randomisations, provided that doing so does not lead to premature unblinding of results for ongoing comparators.

[b] Measured by Oxford immunoassay. 8 x$10^6$ represents the threshold shown to distinguish evidence of prior infection from no evidence of prior infection with sensitivity and specificity >98%.[7] Exploratory analyses using other assays may determine appropriate cut-offs based on blinded inspection of associations between assay results and baseline characteristics and mortality.

Non-randomised exploratory analyses will be used to explore the likely influence of different levels of convalescent plasma antibody concentration on the efficacy of convalescent plasma.

Additional analyses will set the results for children (<18 years) and pregnant women in the context of the overall results.

#### 5.8 Adjustment for baseline characteristics

The main analyses described above will be unadjusted for baseline characteristics. However, if there are any important imbalances between the randomised groups in key baseline pre-specified subgroups (see section 5.4) or allocation in the orthogonal components of the main randomisation, where applicable, emphasis will be placed on analyses that are adjusted for the relevant baseline characteristic(s). This will be done using Cox regression for the estimation of adjusted hazard ratios and a log-binomial regression model for the estimation of adjusted risk ratios.

#### 5.9 Significance levels and adjustment of p-values for multiplicity

Evaluation of the primary trial (main randomisation) and secondary randomisation will be conducted independently, and no adjustment be made for these. Formal adjustment will not be made for multiple treatment comparisons, the testing of secondary and subsidiary outcomes, or subgroup analyses. However, due allowance for multiple testing will be made in the interpretation of the results: the larger the number of events on which a comparison is based and the more extreme the P-value after any allowance has been made for the nature of the particular comparison (i.e. primary or secondary; pre-specified or exploratory), the more reliable the comparison and, hence, the more definite any finding will be considered. 95% confidence intervals will be presented for the main comparisons.

#### 5.10 Statistical software employed

The statistical software SAS version 9.4 and R Studio 3.6.2 (or later) for Windows will be used for the interim and final analyses.

#### 5.11 Data standards and coding terminology

Datasets for analysis will be prepared using CDISC standards for SDTM and ADaM. Wherever possible, clinical outcomes (which may be obtained in a variety of standards, including ICD10 and OPCS-4) will be coded using MedDRA version 20.1.

### 6 SAFETY DATA

Suspected serious adverse reactions (SSARs) and suspected unexpected serious adverse reactions (SUSARs) will be listed by trial allocation.

For each of the following, counts and percentages will be presented by randomised group. Where possible, the absolute risk differences will also be presented with confidence intervals:

###### *6.1.1.1 Cause-specific mortality*

Cause-specific mortality (COVID-19, other infection, cardiac, stroke, other vascular, cancer, other medical, external, unknown cause) will be analysed in a similar manner to the primary outcome.

###### *6.1.1.2 Major cardiac arrhythmia*

Type of arrhythmia will also be described: (i) atrial flutter or fibrillation; (ii) supraventricular tachycardia; (iii) ventricular tachycardia; (iv) ventricular fibrillation; (v) atrioventricular block requiring intervention, with subtotals for (i)-(ii) and (iii)-(iv).

###### *6.1.1.3 Major bleeding*

Type of bleeding will also be described: (i) intracranial bleeding; (ii) gastro-intestinal bleeding; (iii) other bleeding site, and (iv) all sites combined.

###### *6.1.1.4 Early safety of antibody-based therapy*

Additional safety data will be collected in a subset of patients randomised to part B: (i) sudden worsening in respiratory status; (ii) severe allergic reaction; (iii) temperature >39°C or ≥2°C rise since randomisation; (iv) sudden hypotension; (v) clinical haemolysis; and (vi) thrombotic event.

### 7 ADDITIONAL POST-HOC EXPLORATORY ANALYSIS

Any post-hoc analysis requested by the oversight committees, a journal editor or referees will be labelled explicitly as such. Any further future analyses not specified in the analysis protocol will be exploratory in nature and will be documented in a separate statistical analysis plan.

### 8 DIFFERENCES FROM PROTOCOL

The testing of multiple treatment arms will not formally be adjusted for, but given the number of comparisons, due allowance will be made in their interpretation. Formal methods of adjustment for multiplicity were not adopted because of treatment arms being added over time (including the factorial convalescent plasma comparison), unequal recruitment into each arm, and the ultimate number of treatments under evaluation not known in advance.

This analysis plan will be updated prior to unblinding of the 6-month follow-up results. Additional analyses may be specified, e.g. to explore the impact of randomised treatment allocation on hospital re-admission for COVID-19.

### 9 REFERENCES

#### 9.1 Trial documents

Study protocol, case report forms, training materials, and statistical analysis plan are published on the trial website.

#### 9.2 Other references

#### 10 APPROVAL

| | | |
|---|---|---|
| Trial Statistician | Name: Mr Enti Spata | |
| | Signature: | Date: |
| Chief Investigator | Name: Professor Peter Horby | |
| | Signature: | Date: |
| Deputy Chief Investigator | Name: Professor Martin Landray | |
| | Signature: | Date: |
| Steering Committee Statistician | Name: Professor Edmund Juszczak | |
| | Signature: | Date: |
| Steering Committee Statistician | Name: Professor Alan Montgomery | |
| | Signature: | Date: |
| Steering Committee Statistician | Name: Professor Thomas Jaki | |
| | Signature: | Date: |

#### 11 DOCUMENT HISTORY

| Version | Date | Edited by | Comments/Justification | Timing in relation to unblinded interim monitoring | Timing in relation to unblinding of Trial Statisticians |
|---|---|---|---|---|---|
| 0.1 | 20/03/20 | LL/JB | First draft. | Prior | Prior |
| 0.2 | 01/04/20 | LL/JB | Comments and amendments from Martin Landray, Jonathan Emberson & Natalie Staplin. Also aligned with updated protocol and CRFs. | Prior | Prior |
| 0.3 | 01/04/20 | EJ/LL | Further edits and comments. | Prior | Prior |
| 0.4 | 07/04/20 | JB/EJ/LL | Following statistics group meeting on 02/04/20. | Prior | Prior |
| 0.5 | 22/04/20 | JB/LL/EJ | Following statistics group meeting on 09/04/20 and further protocol update. | After | Prior |
| 0.6 | 24/04/20 | LL | Following statistics group meeting on 23/04/20. | After | Prior |
| 0.7 | 10/05/20 | LL | Protocol update. | After | Prior |
| 0.8 | 15/05/20 | LL | Following statistics group meeting on 15/05/20. | After | Prior |
| 0.9 | 27/05/20 | LL | Further comments from TSC members prior to interim analysis on 28/05/20. | After | Prior |
| 1.0 | 09/06/20 | LL | Revised following the stopping of the hydroxychloroquine arm, and prior to the trial statisticians receiving unblinded data for this arm. | After | Prior |
| 1.1 | 21/06/20 | LL/JB/RH | Additional clarification of ventilation denominators. Adjustment for any imbalances of subgroup characteristics between treatment arms at randomisation. Clarification of analysis of composite outcome. Removal of 'Unknown' ethnicity subgroup. Addition of section 5.5 Adjustment for baseline characteristics. | After | After unblinding of hydroxychloroquine and dexamethasone arms. |

| Version | Date | Edited by | Comments/Justification | Timing in relation to unblinded interim monitoring | Timing in relation to unblinding of Trial Statisticians |
|---|---|---|---|---|---|
| 2.0 | 04/11/20 | EJ/ES | Revised to reflect changes in protocol, including introduction of factorial randomisations and new arms, including convalescent plasma, tocilizumab, synthetic neutralizing antibodies (REGN-COV2, and aspirin. | Prior to interim analysis of aspirin arm<br><br>After interim analyses of all other arms | After unblinding of 28-day results for hydroxychloroquine, lopinavir-ritonavir, and dexamethasone arms.<br><br>Prior to unblinding of any other arms |
| 2.1 | 02/12/20 | ES | Addition of colchicine. Modification of definition of recipient antibody concentration subgroup. | Prior to interim analyses including antibody results or of colchicine arm. | After unblinding of 28-day results for hydroxychloroquine, lopinavir-ritonavir, and dexamethasone arms.<br><br>Prior to unblinding of any other arms |

### Appendix 3: Definition and Derivation of Baseline Characteristics and Outcomes

### Definition and Derivation of Baseline Characteristics and Outcomes

### 1 Version

| Date | Version | Comments |
|---|---|---|
| 06-Jun-2020 | 0.1 | Initial version |
| 08-Jun-2020 | 0.2 | Minor updates |
| 09-Jun-2020 | 1.0 | First released version |
| 11-Dec-2020 | 2.0 | Update to sections 6.4 (use of assisted ventilation) and 6.6 (use of renal replacement therapy) |
| 06-Jan-2020 | 3.0 | Update to clarify the derivation of outcomes and baseline data for the second randomisation and define complete follow-up |

### 2 Scope

This document describes the definition and derivation of the primary, secondary and other outcomes of the RECOVERY trial for the published trial analyses. It should be read alongside the study protocol which defines the study outcomes briefly, and the Statistical Analysis Plan (SAP) which describes the statistical methods used to analyse these outcomes. The SAP refers to this document (see Section 2.6.4 Detailed derivation of outcomes) which provides detail on how the outcomes are defined, captured and derived.

Most outcomes have more than one potential source which improves completeness of capture but also will inevitably identify discrepancies between different sources. This document describes the principles for how such discrepancies are resolved; the rules for this were developed blind to results. Further details of the methods are described in the RECOVERY trial internal operating procedure for identifying data discrepancies.

### 3 Abbreviations

| | |
|---|---|
| ADDE | Annual District Death Extract |
| CCDS | Critical Care Dataset |
| CHESS | COVID-19 Hospitalisation in England Surveillance System |
| CPAP | Continuous Positive Airway Pressure |
| CRP | C-reactive protein |
| ECMO | Extra-corporeal membrane oxygenation |
| eCRF | Electronic Case Report Form |

| | |
|---|---|
| FCE | Finished Consultant Episode |
| FU | Follow-up |
| HESAPC | Hospital Episode Statistics Admitted Patient Care |
| HFNO | High-flow nasal oxygen |
| ICD-10 | International Classification of Diseases 10th edition |
| ICNARC | Intensive Care National Audit and Research Centre |
| IMV | Invasive mechanical ventilation |
| NHSCR | NHS Central Register (Scotland) |
| NIV | Non-invasive ventilation |
| NRS | National Records of Scotland |
| ONS | Office for National Statistics (ONS) |
| OPCS-4 | Office of Population Censuses Surveys Classification of Surgical Operations and Procedures 4th revision |
| PDS | Patient Demographic Service |
| PEDW | Patient Episode Database for Wales |
| RRT | Renal replacement therapy |
| PHE | Public Health England |
| SAP | Statistical Analysis Plan |
| SICSAG | Scottish Intensive Care Society Audit Group |
| SMR | Scottish Morbidity Record |
| SUSAPC | Secondary Use Service Admitted Patient Care |
| UKRR | UK Renal Registry |
| WDSD | Welsh Demographic Service |
| WRRS | Welsh Results Reporting Service |

### 4 Data sources

#### 4.1 Electronic case report forms

##### 4.1.1 Main randomisation

The Randomisation eCRF is completed by hospital staff after patients (or a legal representative) have given consent to participate in the trial. It collects the following participant information:

- Identifiers
  - First name, family name
  - NHS number
  - Date of birth
  - Sex (male/female/unknown)
- Inclusion criteria
  - COVID-19 symptom onset date
  - Date of hospitalisation
- Details of acute illness
  - Requirement for oxygen[1]

[1] NHS England advice published on 9 April 2020 stated that the usual oxygen target saturation for prescribed oxygen should change from 94-98% to 92-96% in the first instance. Hospitals may further reduce this to 90-94% if clinically appropriate according to prevailing oxygen demands. https://www.england.nhs.uk/coronavirus/wp-content/uploads/sites/52/2020/04/C0256-specialty-guide-oxygen-therapy-and-coronavirus-9-april-2020.pdf. Guidance on admission to hospital was similar in Scotland. https://www.nhsggc.org.uk/media/259232/covid-19_gps_national_supporting_guidance_for_scottish_general_practice.pdf although hospital guidelines in Scotland did not specify a target oxygen saturation.

    - Requirement for ventilatory support (none, continuous positive airway pressure, non-invasive ventilation, high-flow nasal oxygen, invasive mechanical ventilation (IMV) or extra-corporeal membrane oxygenation) (ECMO)
    - Latest oxygen saturation
    - Latest C-reactive protein, creatinine and D-dimer measurement (if available)
- Comorbidities
    - Diabetes
    - Heart disease
    - Chronic lung disease
    - Tuberculosis
    - HIV
    - Severe chronic liver disease
    - Severe kidney impairment (eGFR <30 mL/min/1.73m$^2$ or on dialysis)
    - Long QT syndrome
    - Pregnancy
- Current treatment
    - Macrolide antibiotics
    - Aspirin or other antiplatelet therapy
    - Warfarin or direct oral anticoagulant
    - Venous thromboembolism prophylaxis (standard or increased dose due to COVID-19)
    - Remdesivir
    - Systemic corticosteroids
- Other
    - Weight (children only)

##### 4.1.2 Second randomisation

The Second Randomisation eCRF is completed by hospital staff when they wish to randomise participants between tocilizumab or standard care alone if they fulfil the protocol-defined oxygenation and inflammation criteria. It collects the following participant information:

- Inclusion criteria
    - Requirement for oxygen
    - Current level of ventilation support (none/CPAP/NIV/HFNO/IMV/ECMO)
    - Latest CRP
- Other information
    - Latest ferritin and creatinine

##### 4.1.3 Convalescent plasma safety eCRF

This eCRF is completed by hospital staff as soon as possible after 72 hours post-main randomisation for participants who entered the convalescent plasma comparison. It collects the following information:

- Adherence to convalescent plasma allocation (number of units received, whether any were stopped early)
- Adverse events
    - Sudden worsening of respiratory status

- Severe allergic reaction
- Temperature ≥39C (or rise ≥2C above baseline)
- Sudden hypotension
- Clinical haemolysis
- Thrombotic event

##### 4.1.4 Follow-up

The FU eCRF is completed by hospital staff at the earliest of (i) discharge from acute care (see Section 6.3 below), (ii) death, or (iii) 28 days after the main randomisation. It collects the following information from date of randomisation onwards:

- Adherence to randomised allocation, and receipt of other study treatments or remdesivir (and number of days of treatment)
- COVID diagnostic test result
- Vital status and underlying cause of death (COVID, other infection, cardiovascular, other; if other, a free text description is collected)
- Date of discharge
- Requirement for assisted ventilation (CPAP, NIV, HFNO, IMV, ECMO) and number of days of assisted ventilation and IMV/ECMO separately
- Occurrence of major cardiac arrhythmia (atrial flutter/fibrillation, supraventricular tachycardia, ventricular tachycardia [including torsades de pointes], ventricular fibrillation or bradycardia requiring intervention) (from 12 May 2020)
- Occurrence of thrombotic event (pulmonary embolism; deep-vein thrombosis; ischaemic stroke; myocardial infarction; systemic arterial embolism; other) (from 6 November 2020)
- Occurrence of clinically-significant bleeding i.e. intracranial or requiring intervention (blood transfusion; surgery; endoscopy; vasoactive drug or blood transfusion), by site (intra-cranial; gastrointestinal; other) (from 6 November 2020)
- Requirement for renal replacement therapy

#### 4.2 Registries and NHS datasets

##### 4.2.1 Hospital admissions datasets

###### *4.2.1.1 Secondary Use Service Admitted Patient Care*

The SUSAPC dataset is a repository of data hosted by NHS Digital that relates to in-patient care provided in England, which aims to enable reporting and analyses to support the NHS in the delivery of healthcare services. These data are submitted on a regular basis by NHS hospital trusts and at pre-arranged dates during the year. Submissions are consolidated, validated and cleaned and then incorporated into the HESAPC dataset. Data may be incomplete in places and is not quality assured to the same extent as HES, but is available more rapidly.

In the SUSAPC dataset, each record contains data relating to a continuous period of care under one consultant known as a Finished Consultant Episode (FCE). FCEs can be grouped together to form 'Spells'. Each spell is a continuous periods of inpatient care within one hospital. Each FCE contains data about the patient (e.g. sex, ethnicity), the specialty providing the care (e.g. cardiology), ICD-10 diagnostic and OPCS-4 procedure codes, along with dates for each procedure and details about the admission and discharge and other data.

For the main RECOVERY analyses the following data are used;

- Ethnicity
- Sex

- Date of admission and discharge
- Start and end date of the FCE
- Discharge method and destination (which may indicate death of participant)
- Diagnoses recorded during FCE (ICD-10 coded)
- Procedures performed during FCE (OPCS-4 coded) and corresponding dates

Linked SUSAPC data are imported to the RECOVERY trial database approximately twice a month.

###### *4.2.1.2 Hospital Episode Statistics Admitted Patient Care*

HESAPC contains data relating to admissions to NHS hospitals in England and is produced from the SUSAPC following a number of cleaning and validation steps. For participants in England, HESAPC is available for the 5 year period prior to enrolment in the study. For the main RECOVERY analyses these data are used to identify prior medical conditions on the basis of recorded ICD-10 and OPCS-4 codes (excluding the admission during which the patient was randomised). Linked HESAPC data are imported to the RECOVERY trial database quarterly.

###### *4.2.1.3 NHS Central Register Scottish Morbidity Record One*

The NHSCR SMR01 data set holds episode level data on hospital inpatient and day case discharges from acute specialities from hospitals in Scotland. The data fields used in the RECOVERY trial are equivalent to those used in SUSAPC and HESAPC. Linked NHSCR-SMR01 data are imported approximately twice a month.

###### *4.2.1.4 Patient Episode Data Wales*

PEDW contains data relating to admissions to NHS hospitals in Wales. Linked data for RECOVERY participants recruited via sites in Wales will be available for future analysis.

##### 4.2.2 Mortality datasets

###### *4.2.2.1 Patient Demographic Service*

The PDS is the electronic database of NHS patient details such as name, address, date of birth and NHS Number for patients in England. For RECOVERY it is used to provide information on fact and date of death. It provides both ‘informal’ notifications of death (which occur when a health care provider is informed of their patients death and records the reported date of death in their electronic data systems) and ‘formal’ notifications of death (which are provided by the Office for National Statistics).

###### *4.2.2.2 Office for National Statistics Mortality data*

The ONS mortality data contains information related to a person's death taken from the death certificate for all deaths registered in England and Wales. The following data are provided

- The underlying cause of death
- Contributory causes of death
- Other conditions recorded on the death certificate but not contributing to death
- Whether a post-mortem took place

Clinical data are recorded using ICD-10 codes. Linked ONS mortality data are imported into the RECOVERY trial via a monthly extract from NHS Digital.

###### *4.2.2.3 Welsh Demographic Service*

WDS data are the electronic database of NHS patient details for patients in Wales and are similar to PDS (4.2.2), providing fact and date of death (including formal or informal notifications). Linked data for RECOVERY participants recruited via sites in Wales will be available for future analysis.

###### *4.2.2.4 National Records of Scotland Mortality Data*

The NRS mortality data contain information related to a person's death taken from the death certificate for all deaths registered in Scotland. The data provided includes the date of death and the underlying and contributory causes of death coded in ICD-10. Linked data are imported into the RECOVERY trial database approximately twice a month.

##### 4.2.3 COVID specific datasets

###### *4.2.3.1 Public Health England Second Generation Surveillance data*

The SGSS is an application that captures, stores and manages routine laboratory surveillance data on infectious diseases and antimicrobial resistance from laboratories across England. Once the reports have been loaded into SGSS, each record is subject to a number of validation processes, and local LIMS codes are translated to SGSS codes to standardise the data for analysis. The data is stored in a central database within PHE and details of tests indicating SAR-CoV-2 have been made available to NHS Digital for dissemination for a limited time period. For each test, the following data are available

- Date the sample was collected
- Date the result was reported
- Organism identified (only SARS-CoV-2)

Linked PHE SGSS data are imported into the RECOVERY trial on approximately twice a month.

###### *4.2.3.2 Public Health Scotland COVID-19 laboratory antigen test positive list*

The Electronic Communication of Surveillance in Scotland (ECOSS) collects routine laboratory surveillance data on infectious diseases from laboratories in Scotland. The data provided to RECOVERY is limited to SARS-CoV-2 results along with the date of the sample and result. .

###### *4.2.3.3 Welsh Results Reporting Service Pathology Data*

The WRRS contains all Pathology Test Results for Wales in a single database. Tests indicating a positive SAR-CoV-2 antigen linked to the trial participants are obtained.

###### *4.2.3.4 COVID-19 Hospitalisation in England Surveillance System*

PHE has established the COVID-19 Hospitalisation in England Surveillance System (CHESS), which collects epidemiological data (demographics, risk factors, clinical information on severity, and outcome) on COVID-19 infection in patients requiring hospitalisation and ICU/HDU level care. This dataset has been made available to NHS Digital for dissemination for a limited time period. For RECOVERY the following information is used;

- Date of ICU/HDU admission and discharge
- Use of respiratory support during the admission (including oxygen via cannulae or mask, high flow nasal oxygen, non-invasive ventilation, invasive mechanical ventilation and ECMO)
- Complications during the admission (including viral pneumonia, secondary bacterial pneumonia, ARDS, unknown, and other co-infections)

The CHESS dataset is imported into the RECOVERY trial approximately twice a month.

###### 4.2.3.5 GPES Data for Pandemic Planning and Research (COVID-19) (GDPPR)

GDPPR data is available for RECOVERY participants in England. Data includes patient demographic information and coded medical information (mainly in SNOMED codes).

##### 4.2.4 Intensive Care Datasets

###### 4.2.4.1 Intensive Care National Audit and Research Centre

The ICNARC Case Mix Programme is the national clinical audit covering all NHS adult, general intensive care and combined intensive care/high dependency units in England, Wales and Northern Ireland, plus some additional specialist and non-NHS critical care units. Data are collected about the first 24 hours in ICU/HDU and at discharge from the ICU/HDU with a further data collection point after discharge from hospital. For RECOVERY, the following data recorded at discharge from ICU/HDU are used:

- Date of admission to and discharge from ICU/HDU
- Use of Advanced Respiratory Support (ARS), Basic Respiratory Support (BRS) or Renal Support during the admission
- The number of days of ARS, BRS or Renal Support during the admission
- Date of death (if relevant)

Linked ICNARC data is requested for hospitals recruiting to RECOVRY and are imported approximately twice a month.

###### 4.2.4.2 Scottish Intensive Care Society Audit Group

SICSAG collects data from all general adult Intensive Care Units, Combined Units and the majority of High Dependency Units in Scotland using the WardWatcher system. The following data are used in the RECOVERY trial:

- Date of admission and discharge from ICU/HDU
- Used of mechanical ventilation via endotracheal tube or tracheostomy and use of haemofiltration for each day of during admission

Linked SICSAG data are imported into the RECOVERY trial approximately twice a month.

###### 4.2.4.3 Critical Care dataset

In England and Wales much of the key data collected by ICNARC is also available in the CCDS from NHS Digital or the SAIL datalink Wales. However, both the ICNARC and CCDS data can be subject to different delays during collection, consolidation and dissemination and therefore either source may be incomplete at any one time-point. Both sources are therefore combined to provide information about ICU/HDU care for participants in England and Wales.

##### 4.2.5 Disease specific registries

###### 4.2.5.1 UK Renal Registry

Data from the UK Renal Registry will be available at a later date.

### 5 Baseline characteristics

Baseline characteristics for the trial cohort are obtained from the first randomisation eCRF for the main randomisation comparisons. For the second randomisation comparisons, the baseline data are obtained either from the second randomisation form directly (e.g. baseline use of respiratory support) or from a calculation based on the first randomisation form data and the number of days between the first and second randomisation forms (e.g. days since symptom onset).

Where fields are missing, they may be supplemented by data from the linked health care data. Generally corrections to the randomisation eCRF data are not made. Exceptions to this would include key participant identifiers (Date of birth, NHS or CHI number, sex) or cases where information is missing. For example, if a site later report that the date of birth was entered incorrectly, this would be confirmed with the site (recorded in the trial data query system) and updated (with appropriate audit trail).

###### 5.1.1 Baseline corticosteroid use

Baseline steroid use is determined as follows:

- Baseline steroid use = yes if allocated dexamethasone in main randomisation OR responded 'yes' to baseline steroid question on main randomisation form (OR [for tocilizumab comparison only] responded 'yes' to baseline steroid question on second randomisation form
- Otherwise, Baseline steroid use = no if answered 'no' to steroid question on main OR [for tocilizumab comparison only] second randomisation forms
- Otherwise, Baseline steroid use = not asked if recruited prior to June 18th[2]
- Otherwise, Baseline steroid use = unknown

For the purposes of analysis, baseline steroid use = no and not asked will be combined for subgroup analyses. Participants with baseline steroid use = unknown will be exluded from subgroup analysis, but the number in this subgroup provided in a footnote.

##### 5.2 Additional baseline characteristics

Some baseline characteristics that are not collected on the randomisation eCRF may be extracted from registry data or other sources. These include:

- Ethnicity by Office for National Statistics 2001 census categories (White, BAME [Mixed, Asian or Asian British, Black or Black British, Other Ethnic Groups], Unknown) from linked health care records. Ethnic groups characterised using SNOMED codes within the GDPPR data are mapped to these categories. Where ethnicity records are discrepant between individual episodes in HES/SMR01/PEDW, the most frequently recorded code is used. Where there is discrepancy between this code and the ethnic group recorded in the GDPPR data, the GDPPR code is used.
- Confirmed SARS-CoV-2 diagnostic test from linked health care records. A positive SARS-CoV-2 with a test date within 28 days of the date of first randomisation is considered as confirmed SARS-CoV-2. In the absence of such data for a participant, the data from the randomisation eCRF may be used.
- Comorbidity score: It is possible to calculate comorbidity and frailty scores (e.g. Charlston Comorbidity Score) from prior linked hospital admissions data and this will be done for future exploratory analyses (not specified in the trial SAP).
- Prior End Stage Kidney Disease (see section 6.6)
- Risk: The risk of death by 28 days can be modelled using available baseline characteristics (in the overall trial population) and a risk score derived. Participants will be divided into thirds based on this score (such that each third has approximately the same number of deaths), with the tertiles rounded to clinically-relevant values. For the main trial analyses the groups will defined as risk of death by 28 days of <30%; ≥30 ≤45%; and >45%.

[2] From 18th June onwards a question on baseline systemic corticosteroid use was added to the main randomisation form following the release of the dexamethasone comparison results.

### 6 Outcomes

#### 6.1 All-cause mortality

The primary outcome is all-cause mortality at 28 days after randomisation. All-cause mortality will also be assessed at 6 months and other later time points.

##### 6.1.1 Sources

Information on death may come from the following sources:

- FU eCRF (for deaths within first 28 days after randomisation)
- PDS (for participants in England)
- PDS Wales ((or participants in Wales)
- SUSAPC (for participants in England)
- SMR01 (for participants in Scotland)
- PEDW (for participants in Wales)
- ONS mortality data (for participants in England and Wales)
- NRS mortality data (for participants in Scotland)

In general, the primary source will be considered ONS (which includes formal death notification within PDS) and NRS mortality data as these are the official national death registries.

##### 6.1.2 Discrepancies

###### *6.1.2.1 Fact of death*

The ONS and NRS mortality data will be considered the defining source for fact of death. In order to allow rapid analysis of results, other sources (e.g. informal death notification via PDS, report of death on the FU eCRF, report of death from SUSAPC) are used for DMC and interim analyses. Cases where these reports are not later substantiated by ONS or NRS are individually reviewed and are not considered as deaths, unless a suitable explanation exists.

###### *6.1.2.2 Date of death*

The ONS and NRS data will be considered the defining source for date of death. In order to allow rapid analysis of data, other sources may be used. Where data sources are discrepant the following hierarchy is applied;

- ONS/NRS (most reliable for date of death), then
- Linked hospital admissions data, then
- FU eCRF , then
- PDS informal death notification (least reliable for date of death)

#### 6.2 Cause-specific mortality

The cause of death for the 28 day analysis will be the underlying cause of death as provided by ONS. The causes of death will be categorised as follows:

- Non-vascular death
    - Death from infection
        - Death from COVID-19
        - Death from other infection
    - Death from cancer
    - Death from other medical causes
    - External deaths
- Vascular death

    - Cardiac death
    - Stroke death
    - Other vascular death
- Unknown death

The ICD-10 codes contributing to these categories are shown in Appendix 1.

#### 6.3 Time to discharge

Time to discharge (which is a more accurate term for duration of admission because only the period from randomisation onwards is relevant) is defined as the number of days a participant remained in hospital for acute care after randomisation. Discharge excludes transfer to another acute hospital, but might include transfer to community hospital for rehabilitation or a hospice for end-of-life care.

##### 6.3.1 Sources

Information on date of discharge may come from the following sources:

- FU eCRF
- SUSAPC (for participants in England)
- PEDW (for participants in Wales)
- SMR01 (for participants in Scotland)

The participant is considered to have transferred between hospitals (i.e. not discharged) if there is another admission to a hospital on that, or the next, day where either the method or source of the admission recorded indicates transfer from another hospital. The first date of discharge which does not fulfil these criteria for an inter-hospital transfer after first or second randomisation is used to determine time to discharge.

##### 6.3.2 Discrepancies

Linked hospital admissions data will be used if date of discharge is discrepant with FU eCRF data. If no linked hospital admissions data are available and the FU eCRF indicates discharge without a date, the date of completion for the FU eCRF will be used.

#### 6.4 Use and duration of ventilation

Assisted ventilation can be broadly divided into

i. Invasive mechanical ventilation (IMV) which includes ECMO (a secondary outcome in combination with all-cause mortality)
ii. Non-invasive ventilation which includes CPAP, NIV and HFNO (which are included in the subsidiary outcomes)

Information on non-invasive ventilation was collected because at the time the trial was designed there were concerns that the availability of mechanical ventilators would be insufficient to meet demand, so some patients would be treated with non-invasive ventilation when in other circumstances they would have received invasive mechanical ventilation. In reality this situation did not occur, so the emphasis of the analyses (and efforts to resolve discrepancies) is on invasive mechanical ventilation.

##### 6.4.1 Sources

Information on ventilation may come from the following sources:

- FU eCRF
- SUSAPC/SMR01/PEDW
- ICNARC

- SICSAG
- CHESS
- CCDS

However, the coding of ventilation is different in each source.

##### 6.4.2 Fact of assisted ventilation

A participant is considered to have received IMV/ECMO if use of these treatments was recorded on the FU eCRF; if a relevant procedure code was recorded in SUSAPC/SMR01/PEDW within 28 days of randomisation (Appendix 2); if days of advanced respiratory support (ARS) in the ICNARC/CCDS data were considered to fall between randomisation and 28 days (see section 6.4.3) or if the daily SICSAG record indicated that the participant was receiving respiratory support via an endotracheal tube or tracheostomy.

A participant is considered to have received non-invasive ventilation if the site recorded 'yes' to the question 'did the participant receive assisted ventilation' or 'yes' to any of the individual types of non-invasive ventilation (CPAP, BIPAP, HFNO) on the FU eCRF; if a relevant procedure code was recorded in SUSAPC/SMR01/PEDW within 28 days of randomisation (Appendix 2) or if use of HFNO or NIV was recorded in CHESS when the admission and discharge date were both between randomisation and 28 days.

##### 6.4.3 Duration of invasive mechanical ventilation

The data from the critical care datasets (ICNARC, CCDS and SICSAG) are considered the primary source of the duration of IMV. Within ICNARC/CCDS, ARS is considered to be equivalent to IMV, however only the dates of admission and discharge from ICU/HDU and the number of days of ARS are provided. The days of ARS within each critical care episode are assumed to be continuous. The days of ARS were assumed to include randomisation if the participant was recorded as receiving IMV at baseline on the first or second randomisation eCRF as appropriate. Otherwise, the days of ARS are assumed to start from admission to critical care, occur at the mid-point of the critical care admission or end on discharge from critical care depending on the level of care recorded on admission and discharge and, in some cases, the destination on discharge (Appendix 3). Using these assumptions, the information from both ICNARC and the CCDS were used to identify whether IMV was received on each of the 28 days following randomisation. The SICSAG daily record indicated use of IMV on each day.

If no relevant information on IMV is received from ICNARC/CCDS/SICSAG, then the duration of IMV was obtained from the FU eCRF. Cessation of mechanical ventilation is deemed successful if it occurs within (and the participant survives until) 28 days after randomisation.

#### 6.5 Major cardiac arrhythmia

Major cardiac arrhythmias are defined as either:

i. Atrial flutter or fibrillation
ii. Supraventricular tachycardia
iii. Ventricular tachycardia (including torsades de pointes)
iv. Ventricular fibrillation
v. Significant bradycardia (requiring intervention)

##### 6.5.1 Sources

Information on cardiac arrhythmias is collected on the FU eCRF (but only for those eCRFs completed from 12 May 2020 onwards when these outcomes were added).

#### 6.6 Renal replacement therapy

Renal replacement therapy (RRT) includes haemodialysis, haemofiltration (and their combination) and peritoneal dialysis. (Kidney transplantation is not relevant in this case.) Individuals receiving RRT at baseline are identified as follows;

- Patients already receiving renal replacement for End Stage Kidney Disease at baseline are identified using linked hospitalisation data (appendix 4).
- From the ICNARC/CCDS data, the combination of the number of Renal Support Days and the start and end date of a critical episode may imply that they must have been receiving renal support at randomisation.
- The SICSAG daily record indicates that Renal Support was received on the day of, or on the day before randomisation.
- A procedure code in SUS/SMR01/PEDW indicating dialysis or haemofiltration with a date within the 3 days prior to first or second randomisation as appropriate (appendix 2).
- (When available) A record of prior RRT (without documented recovery) from the UK Renal Registry

##### 6.6.1 Sources

- FU eCRF
- Linked hospitalisation data (SUSAPC, HES, PEDW, SMR01)
- ICNARC
- SICSAG
- UKRR

##### 6.6.2 Discrepancies

Use of RRT is collected on the FU eCRF. Use of RRT is also identified within the linked hospitalisation data from relevant OPCS-4 codes (Appendix 2). Use of RRT in the ICNARC/CCDS is identified from the recording of Renal Support days where the both the date of admission to and discharge from critical care fall between randomisation and 28 days. The SICSAG daily record indicates RRT if Renal Support is recorded on any day between randomisation and 28 days.

Further information on renal outcomes may become available from the UK Renal Registry data.

### 7 Competeness of Follow-up

For the 28 day analysis, follow-up information is considered to be complete if a FU eCRF has been completed, or data has been received from a hospital admissions dataset (SUSAPC, PEDW or SMR01) which includes data from the admission during which the participant was randomised.

### 8 Appendix 1: Cause-specific mortality categories

| Category | Label | ICD-10 codes[1] |
|---|---|---|
| COVID-19 | DTH_COVID | U07.1;U07.2 |
| Other infection | DTH_OTHER_INFECTION | A00*-A99*;B00*-B99*; G00*-G08*; H60*; H62.0-H62.4; H65*-H67*; I33.0; J00*-J22*; J350; J36*-J37*;J39.0; J39.1; J40*-J42*; K61*; K63.0; K67*; L03*-L04*; M00*-M018*; M462*-M465*; M490*-M493*; M600*; M650*- M651*; M710*; M711*; M730*; M731*; M86*; M866*-M869*; M900*; N75.1; O23*; O26.4; O85*; O86.0-I86.3; O86.8; O91*; O98*; P35*-P39*; U04; U04.9 |
| Infection | DTH_INFECTION | DTH_COVID or DTH_OTHER_INFECTION |
| Cancer | DTH_CAN_ANY | C00*-C97* |
| Other medical | DTH_OTHMED | DTH_NONVASC not (DTH_CAN_ANY or DTH_INFECTION or DTH_EXTERNAL) |
| External causes | DTH_EXTERNAL | S00*-Y98* |
| Non-vascular | DTH_NONVASC | DTH_INFECTION or DTH_CAN_ANY or DTH_OTHMED or DTH_EXTERNAL |
| Cardiac | DTH_CARDIAC | I00*-I09*; I11*; I13*; I20*-I25*; I271; I27.8; I27.9; I30.9-I32.0; I32.8; I33.9-I51.5; I51.7-I52* |
| Stroke | DTH_STR_ANY | I60*-I66*; I69* |
| Other vascular | DTH_OTH_VASC | I10*; I15*; I26*; I27.0; I27.2; I28*; I51.6; I67*; I68*; I70*-I83*; I86*-I97*; I98.0, I98.1; I99* |
| Vascular | DTH_VASC | DTH_CARDIAC or DTH_STR_ANY or DTH_VASC |
| Unknown | DTH_UNK | R00*-R99* |

[1] For example, I2* includes all codes beginning with I2.

ICD-10 5th edition (implemented in the NHS in 2016)

### 9 Appendix 2: OPCS-4 and ICD-10 codes used to identify assisted ventilation and other outcomes in the linked hospitalisation data

| Outcome | code | Code type | Description |
|---|---|---|---|
| Use of CPAP | E85.6 | OPCS | Continuous positive airway pressure |
| Use of NIV | E85.2 | OPCS | Non-invasive ventilation NEC |
| Use IMV | E85.1 | OPCS | Invasive ventilation |
| Use of ECMO | X58.1 | OPCS | Extracorporeal membrane oxygenation |
| Use of RRT | X40.1 | OPCS | Renal dialysis |
| | X40.3 | OPCS | Haemodialysis NEC |
| | X40.4 | OPCS | Haemofiltration |

(OPCS and ICD-10 codes used to identify serious arrhythmia and other non-fatal outcomes to be added at a later date.)

### 10 Appendix: 3: Rules for determining start/end of advanced respiratory support days in the critical care datasets

Information is available in ICNARC/CCDS on

- The start and end date of the critical care episode
- The level of care at admission to the unit
- The level of care at discharge from the unit
- The reason for discharge from the unit
- The number of days of Advance Respiratory Support (ARS) received during the episode

The table below defines the rules for deciding whether the days on ARS in an ICNARC/CCDS episode should count from admission onwards (A), before discharge (D) or at the midpoint between admission and discharge (M)

| | | Level of care at admission to the unit | | | | |
|---|---|---|---|---|---|---|
| | | 0 | 1 | 2 | 3 | blank |
| Level of care at discharge from the unit | 0 | M | M | M | A | A |
| | 1 | M | M | M | A | A |
| | 2 | M | M | M | A | A |
| | 3 | D | D | D | A | D |
| | blank | * | * | * | A | A |

* If the reason for discharge from the unit is 'comparable critical care' or 'more-specialist critical care' then D, otherwise M.

The following definitions are taken from the ICNARC data collection manual Version 3.1 (29 June 2009).

Level 3 – indicated by one or more of the following:

- admissions receiving advanced respiratory monitoring and support due to an acute illness
- admissions receiving monitoring and support for two or more organ system dysfunctions (excluding gastrointestinal support) due to an acute illness
- admissions solely receiving basic respiratory monitoring and support and basic cardiovascular monitoring and support due to an acute illness only meet Level 2

Level 2 – indicated by one or more of the following:

- admissions receiving monitoring and support for one organ system dysfunction (excluding gastrointestinal support) due to an acute illness
- admissions solely receiving advanced respiratory monitoring and support due to an acute illness meet Level 3
- admissions solely receiving basic respiratory and basic cardiovascular monitoring and support due to an acute illness meet Level 2
- admissions receiving pre-surgical optimisation including invasive monitoring and treatment to improve organ system function
- admissions receiving extended post-surgical care either because of the procedure and/or the condition of the admission
- admissions stepping down to Level 2 from Level 3 care

Level 1 – indicated by one or more of the following:

- admission recently discharged from a higher level of care
- admissions receiving a greater degree of observation, monitoring, intervention(s), clinical input or advice than Level 0 care
- admissions receiving critical care outreach service support fulfilling the medium-score group, or higher, as defined by NICE Guidelines 50

Level 0 – indicated by the following:

- admissions in hospital and receiving normal ward care

### 11 Appendix 4: Definition of prior RRT for End Stage Renal Disease

A previously validated algorithm was adapted to identify people requiring dialysis for ESRD from the prior HES/SMR01/PEDW.

Individuals who met the criteria for Rules 2-4 during a hospital admission prior to the admission during which they were randomised were considered to have prior ESRD provided they did not meet the criteria for Rule 1 after meeting the other criteria.

**Rule 1: Kidney Transplantation**

Occurrence of any incident kidney transplant code (with no removal within 90 days), or a prevalent kidney transplant code with no removal having occurred prior to the record.

**Rule 2: Peritoneal maintenance dialysis**

Occurrence of any admission with a peritoneal dialysis code (without diagnosis of acute kidney injury).

**Rule 3: Definite maintenance dialysis**

Occurrence of a dialysis code in a patient who has had:

(a) a diagnostic code for ESRD any time prior to, or within 365 days; or

(b) the insertion of an AV fistula or graft any time prior to, or within 365 days.

**Rule 4: Probable maintenance dialysis**

The occurrence of at least two episodes containing a dialysis code, with at least 90 days between the start of the first recorded dialysis, and the start of any subsequent dialysis (without agnosis of acute kidney injury).

#### Relevant ICD-10 and OPCS-4 codes for Rules 1-4 above

| Group | Category | ICD-10 | OPCS-4 | Description |
|---|---|---|---|---|
| Diagnosis | Acute kidney injury | N17 | | Acute renal failure |
| Diagnosis | End-stage renal disease | N18.0 | | End-stage renal disease |
| Diagnosis | End-stage renal disease | N18.5 | | Chronic kidney disease, stage 5 |
| Diagnosis | End-stage renal disease | Q60.1 | | Renal agenesis, bilateral |
| Dialysis | Dialysis | E85.3 | | Secondary systemic amyloidosis (dialysis related) |
| Dialysis | Dialysis | Y60.2 | | Unintentional cut, puncture, perforation or haemorrhage during surgical and medical care; during kidney dialysis….. |
| Dialysis | Dialysis | Y61.2 | | Foreign object accidentally left in body during surgical and medical care; during kidney dialysis or other perfusion |
| Dialysis | Dialysis | Y62.2 | | Failure of sterile precautions during surgical and medical care; during kidney dialysis or other perfusion |
| Dialysis | Dialysis | Y84.1 | | Other medical procedures as the cause of abnormal reaction of the patient, or of later complication; kidney dialysis |
| Dialysis | Dialysis | Z99.2 | | Dependence on enabling machines and devices, not elsewhere classified; dependence on renal dialysis |
| Dialysis | Dialysis | | X40.1 | Renal dialysis |
| Dialysis | Haemodialysis | T82.4 | | Mechanical complication of vascular dialysis catheter |
| Dialysis | Haemodialysis | Z49.1 | | Care involving dialysis; extracorporeal dialysis |
| Dialysis | Haemodialysis | | X40.3 | Haemodialysis NEC |
| Dialysis | Haemodialysis | | X40.4 | Haemofiltration |
| Dialysis | Insertion of AVF or graft | | L74.1 | Insertion of arteriovenous prosthesis |
| Dialysis | Insertion of AVF or graft | | L74.2 | Creation of arteriovenous fistula NEC |
| Dialysis | Insertion of AVF or graft | | L74.6 | Creation of graft fistula for dialysis |
| Dialysis | Insertion of AVF or graft | | L74.8 | Other specified arteriovenous shunt |
| Dialysis | Insertion of AVF or graft | | L74.9 | Unspecified arteriovenous shunt |
| Dialysis | Insertion of PD catheter | | X41.1 | Insertion of ambulatory peritoneal dialysis catheter |
| Dialysis | Peritoneal dialysis | Z49.2 | | Care involving dialysis; other dialysis |
| Dialysis | Peritoneal dialysis | | X40.2 | Peritoneal dialysis NEC |
| Dialysis | Peritoneal dialysis | | X40.5 | Automated peritoneal dialysis |
| Dialysis | Peritoneal dialysis | | X40.6 | Continuous ambulatory peritoneal dialysis |
| Dialysis | Tunnelled line insertion | | L91.5 | Insertion of tunnelled venous catheter |
| Transplantation | Incident kidney transplant | | M01.2 | Allotransplantation of kidney from live donor |
| Transplantation | Incident kidney transplant | | M01.3 | Allotransplantation of kidney from cadaver NEC |
| Transplantation | Incident kidney transplant | | M01.4 | Allotransplantation of kidney from cadaver heart beating |
| Transplantation | Incident kidney transplant | | M01.5 | Allotransplantation of kidney from cadaver heart non-beating |
| Transplantation | Incident kidney transplant | | M01.8 | Other specified transplantation of kidne |
| Transplantation | Incident kidney transplant | | M01.9 | Unspecified transplantation of kidney |
| Transplantation | Prevalent kidney transplant | N16.5 | | Renal tubulo-interstitial disorders in transplant rejection |
| Transplantation | Prevalent kidney transplant | T86.1 | | Kidney transplant failure and rejection |
| Transplantation | Prevalent kidney transplant | Z94.0 | | Kidney transplant status |
| Transplantation | Prevalent kidney transplant | | M08.4 | Exploration of transplanted kidney |
| Transplantation | Prevalent kidney transplant | | M17.4 | Post-transplantation of kidney examination - recipient |
| Transplantation | Prevalent kidney transplant | | M17.8 | Other specified interventions associated with transplantation of kidney |
| Transplantation | Prevalent kidney transplant | | M17.9 | Unspecified interventions associated with transplantation of kidney |
| Transplantation | Removal of kidney transplant | | M02.6 | Excision of rejected transplanted kidney |
